# Leveraging multiple data types to estimate the true size of the Zika epidemic in the Americas

**DOI:** 10.1101/19002865

**Authors:** Sean M. Moore, Rachel J. Oidtman, K. James Soda, Amir S. Siraj, Robert C. Reiner, Christopher M. Barker, T. Alex Perkins

**Affiliations:** Department of Biological Sciences, University of Notre Dame, USA; Eck Institute for Global Health, University of Notre Dame, USA; Environmental Change Initiative, University of Notre Dame, USA; Institute for Health Metrics and Evaluation, USA; School of Veterinary Medicine, University of California, Davis, USA

## Abstract

Since the first Zika virus (ZIKV) infection was confirmed in Brazil in May 2015, several hundred thousand cases have been reported across the Americas. This figure gives an incomplete picture of the epidemic, however, given that asymptomatic infections, imperfect surveillance, and variability in reporting rates imply that the incidence of infection was likely much higher. The infection attack rate (IAR)—defined as the proportion of the population that was infected over the course of the epidemic—has important implications for the longer-term epidemiology of Zika in the region, such as the timing, location, and likelihood of future outbreaks. To estimate the IAR and the total number of people infected, we leveraged multiple types of Zika case data from 15 countries and territories where subnational data were publicly available. Datasets included confirmed and suspected Zika cases in pregnant women and in the total population, Zika-associated Guillan-Barré syndrome cases, and cases of congenital Zika syndrome. We used a hierarchical Bayesian model with empirically-informed priors that leveraged the different case report types to simultaneously estimate national and subnational reporting rates, the fraction of symptomatic infections, and subnational IARs. In these 15 countries and territories, estimates of Zika IAR ranged from 0.084 (95% CrI: 0.067 − 0.096) in Peru to 0.361 (95% CrI: 0.214 − 0.514) in Ecuador, with significant subnational variability in IAR for every country. Totaling these infection estimates across these and 33 other countries and territories in the region, our results suggest that 132.3 million (95% CrI: 111.3-170.2 million) people in the Americas have been infected by ZIKV since 2015. These estimates represent the most extensive attempt to date to determine the size of the Zika epidemic in the Americas, and they offer an important baseline for assessing the risk of future Zika epidemics in this region.

## Introduction

Zika virus is a mosquito-borne pathogen that was first identified in Uganda in 1947 (Dick et al., 1952). Smaller Zika outbreaks have occurred in Africa, Asia, and the Pacific islands since its discovery, but there had been no confirmed cases in the Americas (excluding Easter Island, Chile) prior to the first confirmation of a Zika case in Brazil in May, 2015 (Fauci and Morens, 2016). Subsequent to its discovery in Brazil, the epidemic spread rapidly and cases were reported throughout the Americas over the next two years (Pan American Health Organization (PAHO), 2018). The Zika epidemic generated a large amount of concern in the public health community and the general public, leading to a declaration of a Public Health Emergency of International Concern by the World Health Organization (WHO) in February, 2016 (WHO, 2016), because of the discovery of a link between ZIKV infection in pregnant women and congenital Zika syndrome (CZS) in newborns (Brasil et al., 2016; Mlakar et al., 2016; Rasmussen et al., 2016; Brady et al., 2019). Following the large epidemic from 2015-2017, substantially fewer Zika cases have been reported in 2018 and 2019 (Pan American Health Organization (PAHO), 2018).

Now that the ZIKV invasion has run its course, there are a number of unanswered questions about what will happen next. If the remaining population at risk is large, then additional outbreaks in the coming years are still possible. On the other hand, modeling has suggested that if a large proportion of the population is now immune, herd immunity will likely prevent another large epidemic for more than a decade (Ferguson et al., 2016). This scenario could lead to a slow buildup of new susceptible individuals over time, and of particular concern, an eventual buildup of susceptible women of childbearing age if a ZIKV vaccine is not licensed and broadly deployed (Ferguson et al., 2016; Lessler et al., 2016; Durham et al., 2018). The number of recent ZIKV infections could also have relevance for the epidemiology of dengue virus (DENV) in the region (Ferguson et al., 2016). There is evidence of an interaction between ZIKV and DENV via the human immune response to infection with either virus (Stettler et al., 2016; Dejnirattisai et al., 2016; Bardina et al., 2017; Andrade and Harris, 2018; Rodriguez-Barraquer et al., 2019). If a ZIKV infection provides any temporary cross-protection to DENV, then the reduction in dengue incidence in several Latin American countries over the past few years (Borchering et al., 2019) could be followed by a large dengue epidemic as this temporary cross-protection wanes.

The Pan American Health Organization (PAHO) reported suspected and confirmed Zika cases for every country and territory in the Americas, but these reported cases vastly underestimate the total number of ZIKV infections due to inadequate surveillance, the non-specificity of ZIKV symptoms, and the high proportion of asymptomatic infections (Lessler et al., 2016; O’Reilly et al., 2018). Underreporting is particularly an issue for pathogens such as ZIKV where the majority of infections are asymptomatic or produce only mild symptoms (Duffy et al., 2009; Mitchell et al., 2019). Estimates of ZIKV infections from blood donors in Puerto Rico through 2016 suggest that almost 470,000 people might have been infected in Puerto Rico alone (Chevalier et al., 2017). High ZIKV seroprevalence estimates from several major cities with populations of more than one million—46% in Managua, Nicaragua (Zambrana et al., 2018), 63-68% in Salvador, Brazil (Netto et al., 2017; Moreira-Soto et al., 2018), and 64% in Recife, Brazil (de Araújo et al., 2016)—also suggest that the 806,928 suspected and confirmed cases reported by PAHO represent only a small fraction of the total number of ZIKV infections.

In this study, we take advantage of several features of the recent Zika epidemic that allow us to estimate national and subnational IARs throughout the Americas. First, due to the WHO emergency declaration, surveillance for Zika began in most countries relatively early in the epidemic, and all countries and territories in the Americas reported case data to PAHO. Second, in addition to reporting suspected and confirmed Zika cases in the entire population, many countries also reported Zika cases among pregnant women, CZS or microcephaly cases, and Guillan-Barré syndrome (GBS) cases, providing additional information about the underlying IAR. Third, a number of countries have published subnational Zika surveillance data, which increases the sample sizes for estimation purposes. The subnational data also allows us to capture spatial heterogeneity in ZIKV IAR within these countries. Using a hierarchical Bayesian model fit to multiple data types we estimated the subnational IARs and reporting rates for each data type. Estimated IAR and reporting rates were then used to extrapolate national-level case data from the rest of the American countries and territories to provide an estimate of the total number of ZIKV infections across this region.

## Methods

### Data

The cumulative numbers of suspected and confirmed Zika cases in each country and territory in the Americas, as well as the number of confirmed CZS cases, were reported by PAHO on a weekly basis through the first week of 2018 (Pan American Health Organization (PAHO), 2018). In addition, PAHO also published country reports following epidemiological week 35 in 2017 that contained additional information, including the total number of cases of microcephaly and Guillan-Barré syndrome (GBS) associated with Zika, where available (Pan American Health Organization (PAHO), 2017a). Because ZIKV infection attack rates could vary significantly within a country, we restricted our primary analysis to countries and territories where we were able to obtain Zika data at a subnational level for at least one data type. In total, we were able to estimate subnational and national ZIKV IARs for 15 countries and territories (SI Table 1).

The data types considered were confirmed Zika cases, suspected Zika cases, microcephaly cases associated with a ZIKV infection in the mother, Zika-associated cases of GBS, and the proportion of blood donors with a positive ZIKV infection (Puerto Rico only). In addition, due to the risk of CZS in newborns, many countries also reported the number of pregnant women with either a suspected or confirmed ZIKV infection, which we treated as independent data points from the confirmed and suspected cases in the entire population. Reporting of Zika-associated microcephaly and GBS cases varied by country, with some reporting only cases associated with a lab-confirmed ZIKV infection and others both confirmed and suspected cases.

Where available, we obtained Zika data at the first administrative level (e.g., province or state) within a country or territory. Lower level data were aggregated to the first administrative level in cases where they were available. National and subnational population estimates were generated from WorldPop 2015 population rasters for each country or territory (WorldPop, 2019; Sorichetta et al., 2015). The number of pregnant women in each country potentially at risk of a ZIKV infection was estimated using 2015 pregnancies rasters from WorldPop, and the number of births at risk for microcephaly were estimated using the WorldPop 2015 births rasters (Tatem et al., 2014). All of the data used in our analysis, along with model code, are located in the Github repository https://github.com/mooresea/Zika_IAR.

## Model

We estimated the national and subnational Zika IAR in each country using a hierarchical Bayesian model with the number of total infections and symptomatic infections treated as latent variables (Figure 1). Short descriptions for each of the model variables and parameters are presented in SI Table (2). The IAR in a population of size *N*_*i*_ in administrative unit *i* is the proportion of the population that was infected, 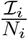, where ℐ_*i*_ is the number of infections. The number of symptomatic infections in a population, *Ƶ*_*i*_, depends on the size of the infected population, ℐ_*i*_, and the probability that an infection is symptomatic, *ρ*_*Ƶ*_, resulting in *Ƶ*_*i*_ ∼ Bin(ℐ_*i*_, *ρ*_*Ƶ*_). The number of confirmed Zika cases in a population, *C*_*T,i*_, depends on the number of symptomatic infections and the local reporting rate, 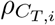, resulting in 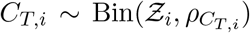. The number of suspected Zika cases was similarly dependent on a reporting rate for suspected cases, 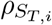, and the number of symptomatic infections in the total population, 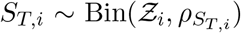. Because misdiagnosis could contribute to the number of suspected cases during an epidemic, we also considered the possibility that there were more suspected cases than symptomatic infections by using a Poisson distribution rather than a binomial distribution to represent the reporting process for suspected cases. The results of this analysis are reported in the Supplementary Information (SI section 4). The numbers of confirmed or suspected cases in pregnant women, *C*_*P,i*_ or *S*_*P,i*_, were represented by binomial distributions dependent on the number of symptomatic infections in pregnant women. The infection attack rate and probability of an infection being symptomatic in pregnant women were assumed to be the same as that in the entire population. The probabilities that a symptomatic infection was reported as a suspected or confirmed Zika case, 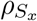 and 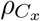, where *x* represents either the entire population (*T*) or pregnant women only (*P*), were assumed to differ between administrative units within a country or territory. To estimate this within-country variation in reporting rates, we assumed that the probability of a symptomatic infection being reported in administrative unit *i* followed a beta distribution with hyperparameters 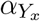 and 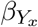 such that 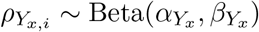, where *Y* denotes confirmed (*C*) or suspected (*S*).

**Figure 1:**
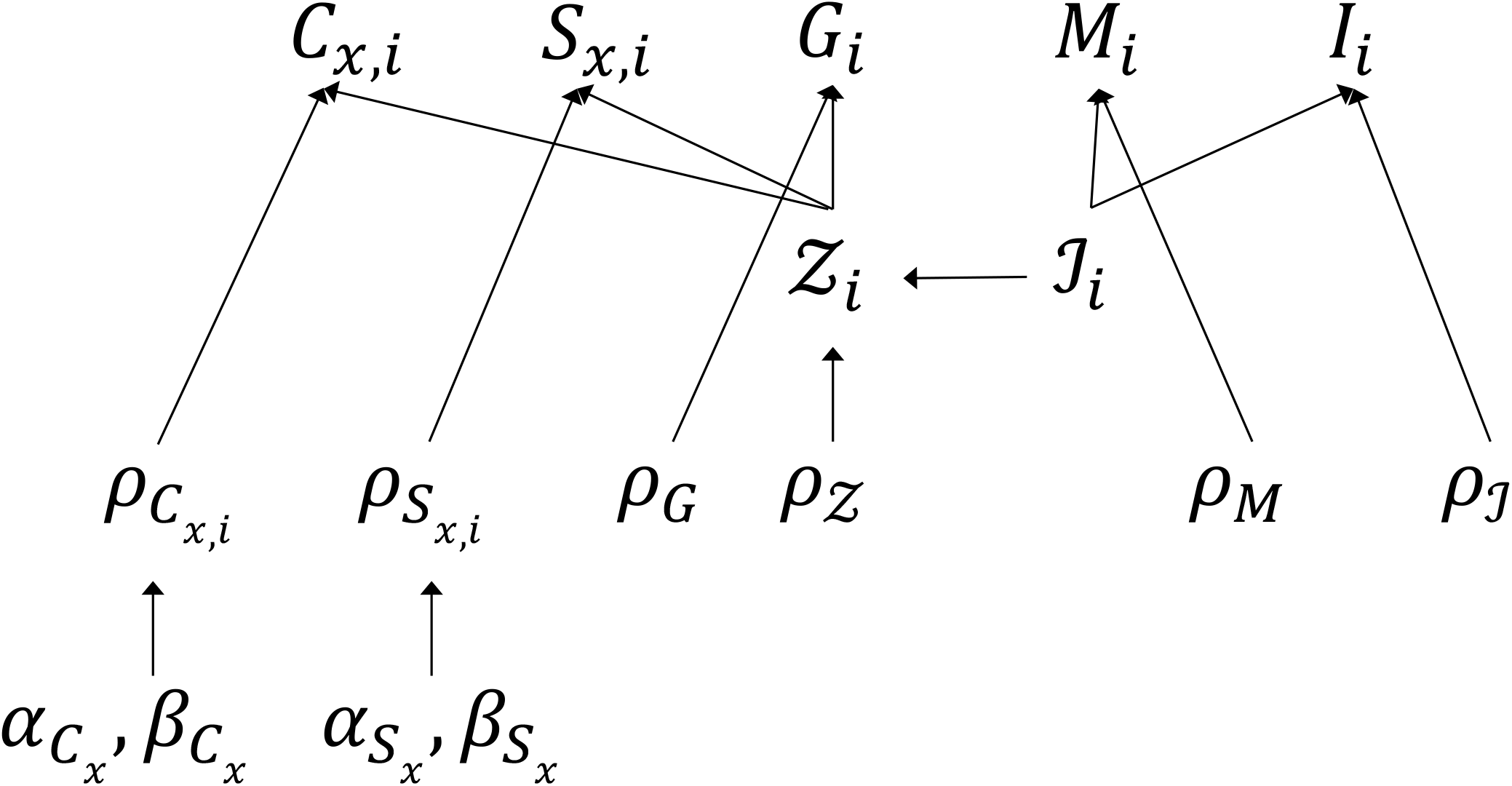
Model schematic. See text and SI Table (2) for description of model parameters and variables.

The number of reported Zika-associated GBS cases was dependent on the number of symptomatic infections, *G*_*i*_ ∼ Bin(*Ƶ*_*i*_, *ρ*_*G*_), where *ρ*_*G*_ is the probability that a symptomatic infection results in a reported GBS case. The number of microcephaly cases associated with Zika was dependent on the total number of births in the population, *B*_*i*_, and the IAR, such that 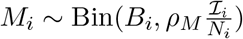, where *ρ*_*M*_ is the probability that an infection during pregnancy results in a reported microcephaly case. The number of blood donors with an active ZIKV infection, *I*_*i*_, within a population of screened blood donors, *D*_*i*_, was related to the IAR by the parameter, *ρ*_*I*_, which corrects for the fact that blood donation is only possible when an individual is asymptomatic and adjusts for the mean duration of viremia in asymptomatic individuals (including pre-symptomatic individuals) relative to the duration of the collection period (Chevalier et al., 2017). As a result, the number of infections among blood donors, *D*_*i*_, is assumed to follow a Poisson distribution, 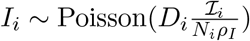 (see SI section 2 for details).

Subnational IARs were estimated for each country and territory using available data types. The national-level IAR was calculated from the total number of subnational infections divided by the national population size, 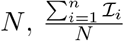. For Puerto Rico, Zika IARs were estimated at the municipality-level (*n* = 78), due to availability of data at that scale. In addition, several datasets there were aggregated at the regional level (*n* = 8), due to their availability at that scale. For these data types, the regional-level IARs were estimated from the total number of infections within all municipalities in a given region.

### Prior assumptions

The IAR reported in previous Zika outbreaks has been as high as 73% on the Micronesian island of Yap (Duffy et al., 2009). The IARs of *Aedes*-transmitted viruses in larger geographical areas tend to be lower than in smaller island environments, such as Yap, because spatial heterogeneity in the presence and abundance of *Aedes* limits transmission potential within a portion of the region (Keeling, 1999). Studies from several different Zika outbreaks have estimated basic reproduction numbers (*R*_0_) of 1.4 to 6.0 (Kucharski et al., 2016; Towers et al., 2016; Lourenço et al., 2017; Shutt et al., 2017; Villela et al., 2017). Based on the theoretical relationship between *R*_0_ and the final epidemic size (Keeling and Rohani, 2011), these *R*_0_ values would correspond to IARs of 0.286–0.833. However, IARs are typically lower at a given *R*_0_ value in populations with heterogeneous contact patterns (Andreasen, 2011), as is typical with transmission by *Ae. aegypti* mosquitoes (Liebman et al., 2014). To lightly constrain our ZIKV IAR estimates without precluding the possibility of values anywhere between 0 and 1, we used a Beta(1, 2) prior for the probability of an individual being infected (i.e., the IAR). This prior distribution had a median value of 0.292 (95% range: 0.013–0.842). We also performed an analysis with a uniform prior for the IARs. A comparison of posterior IAR estimates with and without the Beta(1, 2) prior is included in the Supplementary Information (SI section 4).

Estimates of the probability of a ZIKV infection being symptomatic, *ρ*_*Ƶ*_, have varied considerably across studies (Duffy et al., 2009; Subissi et al., 2017; Mitchell et al., 2019). One recent study estimated the symptomatic probability for three different locations (Yap Island, French Polynesia, and Puerto Rico), taking into account assay senstivity and specificity, as well as the possibility of Zika-like symptoms due to other causes (Mitchell et al., 2019). Median estimates from that study ranged from 27% in Yap to 50% in Puerto Rico. To generate a single prior distribution for *ρ*_*Ƶ*_ in our model, we used the model and data provided in Mitchell et al. (2019) to recreate the posterior estimates of *ρ*_*Ƶ*_ from their analysis, and then fitted a beta distribution to the combined posteriors using the ‘fitdistrplus’ package in R (Delignette-Muller and Dutang, 2015). The resulting distribution was Beta(3.88, 5.34), which has a median of 0.41 and a 95% range of 0.14–0.73, and was used as a prior for each country. The hyperparameters 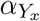 and 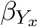 for the reporting probabilities 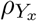, where *Y* = *C* or *S* and *x* = *T* or *P*, were specified as Cauchy(0, 25) priors. This distribution provides a weakly informative prior with the distribution peaked at 0 and a long right tail. We assumed non-informative priors for the probabilities that a symptomatic infection results in a reported Guillan-Barré case or that an infected mother will give birth to a child with reported microcephaly. These probabilities represent not just the probability that an infection leads to a syndromic case, but also that such a case ends up being reported through the surveillance system.

The prior distribution of the incidence correction factor for the probability of infection in asymptomatic blood donors, *ρ*_*I*_, was estimated using the distributions for the incubation period and viremic period described by Chevalier et al. (2017) and fitted to a gamma distribution using the ‘fitdistrplus’ package in R (Delignette-Muller and Dutang, 2015). This resulted in a Gamma(15.97, 0.42) prior. Full details of the estimation of the incidence correction factor are provided in SI section (2).

### Model implementation

Each country or territory model was fitted using the ‘rstan’ version 2.18.2 package in R (Stan Development Team, 2018). For each country or territory, four chains of 5,000 iterations each were run with a burn-in interval of 2,500 iterations. Convergence was assessed using the Gelman-Rubin convergence diagnostic (Gelman et al., 2013). The full model for Peru failed to converge, so the reporting rates of confirmed cases 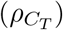 and confirmed cases in pregnant women 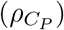 were estimated as single parameters for all administrative units, rather than drawing 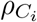 from hyperparameters *α*_*C*_ and *β*_*C*_. We considered this simplification to be justifiable because all ZIKV confirmations were handled by either the national CNS laboratory or one of two regional laboratories, so confirmation rates likely did not vary as widely between administrative units as the reporting rates for suspected cases. Model diagnostic results for each country and territory are provided in SI section (3).

## Model validation

### Posterior predictive checks

Posterior predictive checks were performed by comparing the empirical data to simulated data from the posterior parameter distributions. Posterior predictive data was generated at each iteration, *k*, of the MCMC, with 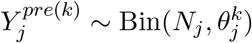 for data type *j* ∈ {*C*_*T*_, *S*_*T*_, *C*_*P*_, *S*_*P*_, *M, G, I*} and its associated parameters *θ*_*j*_. At each iteration, the observed national total, *Y*_*j*_, was compared to the predicted national total, 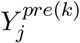. This test statistic was used to calculate a Bayesian p-value 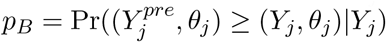, which indicates whether the distribution of the model-generated data was more extreme than the observed data (Gelman et al., 2013). We also compared the observed national totals for each data type to the predicted model output to determine whether the observed data (across all case types and territories) fell within the 95% credible interval (CrI) of the corresponding posterior distributions from the model.

### Holdout analysis

To determine the sensitivity of model estimates to the inclusion of different data types, we fitted the model while holding out one data type at a time. This analysis was restricted to countries where all data types were available at a subnational level (Guatemala and Dominican Republic) or at most one data type was only available at the national-level (Panama). For these countries, we also fitted the model to one data type at a time to assess the benefit of using multiple data types in the estimation process.

### Seroprevalence estimates

As an additional check, we compared modeled IAR estimates to published seroprevalence estimates from the Americas, as these quantities should be comparable. Seroprevalence estimates from at least one location were available for five countries or territories from ten published studies (SI Table 6). The majority of these studies were conducted in a single city and not across a larger adminstrative area, in which case we compared the seroprevalence data to the IAR estimate from the first-level administrative unit where that city was located.

### Applying the model to settings with no spatially disaggregated data

Estimates from the 15 country-specific models were used to make predictions about IARs in the 33 other countries and territories in Latin America and the Caribbean where subnational data was not available. The January, 4 2018 Zika report from PAHO included cumulative data from 52 countries and territories in the Americas (Pan American Health Organization (PAHO), 2018). Canada, Bermuda, and Chile reported no locally-acquired cases, and the United States reported only a few hundred locally-acquired cases in a limited geographic area, so these countries were excluded from the analysis. For the remaining 48 countries and territories (including the 15 modeled territories), the cumulative numbers of confirmed cases, suspected cases, and cases of microcephaly were used to estimate the national IAR. For each of the 48 countries and territories, a national IAR estimate was obtained by drawing from the posterior distributions of the different reporting parameters from each of the 15 country models. This allowed us to draw from across the full range of estimated reporting rates from these 15 countries and territories in predicting the IARs in the remaining countries and territories. For a given country or territory model, *k*, the probability of a given IAR value in country or territory *j* was derived from the joint probability for each of the different data types (*C*_*j*_, *S*_*j*_, and/or *M*_*j*_) that were used to fit that model. The combined probability density function for IAR in country *j* was then taken as the sum of the probability density functions using the parameter estimates from all *K* = 15 models. These IAR estimates were used to calculate the total number of ZIKV infections that may have occurred during the epidemic. Initially, the infections arising in the 15 modeled territories were derived from country- or territory-specific model estimates and only the infections from the remaining 33 countries and territories were estimated from this pooled analysis. The territory-specific model estimates were compared to the pooled model estimates for each of the 15 modeled territories to assess the plausibility of the pooled estimates in the non-modeled countries and territories (SI section 5).

## Results

### Infection attack rate estimates

Estimated Zika infection attack rates at the national level ranged from 0.084 (95% CrI: 0.067 − 0.096) in Peru to 0.361 (95% CrI: 0.214 − 0.514) in Ecuador (Figure 2, SI Table 4). There was considerable heterogeneity in IARs (Figure 3). In Brazil, at the subnational level, the IAR varied from 0.016 (95% CrI: 0.01-0.025) in the State of Paraná to 0.766 (95% CrI: 0.569-0.942) in the State of Sergipe (Figure 3). In Mexico, the IAR ranged from 0 (95% CrI: 0-0) in the Federal District to 0.793 (95% CrI: 0.524-0.963) in the State of Yucatán (SI Figure 17). Subnational IAR estimates for all 15 modeled countries and territories are presented in SI figures (7-21).

**Figure 2:**
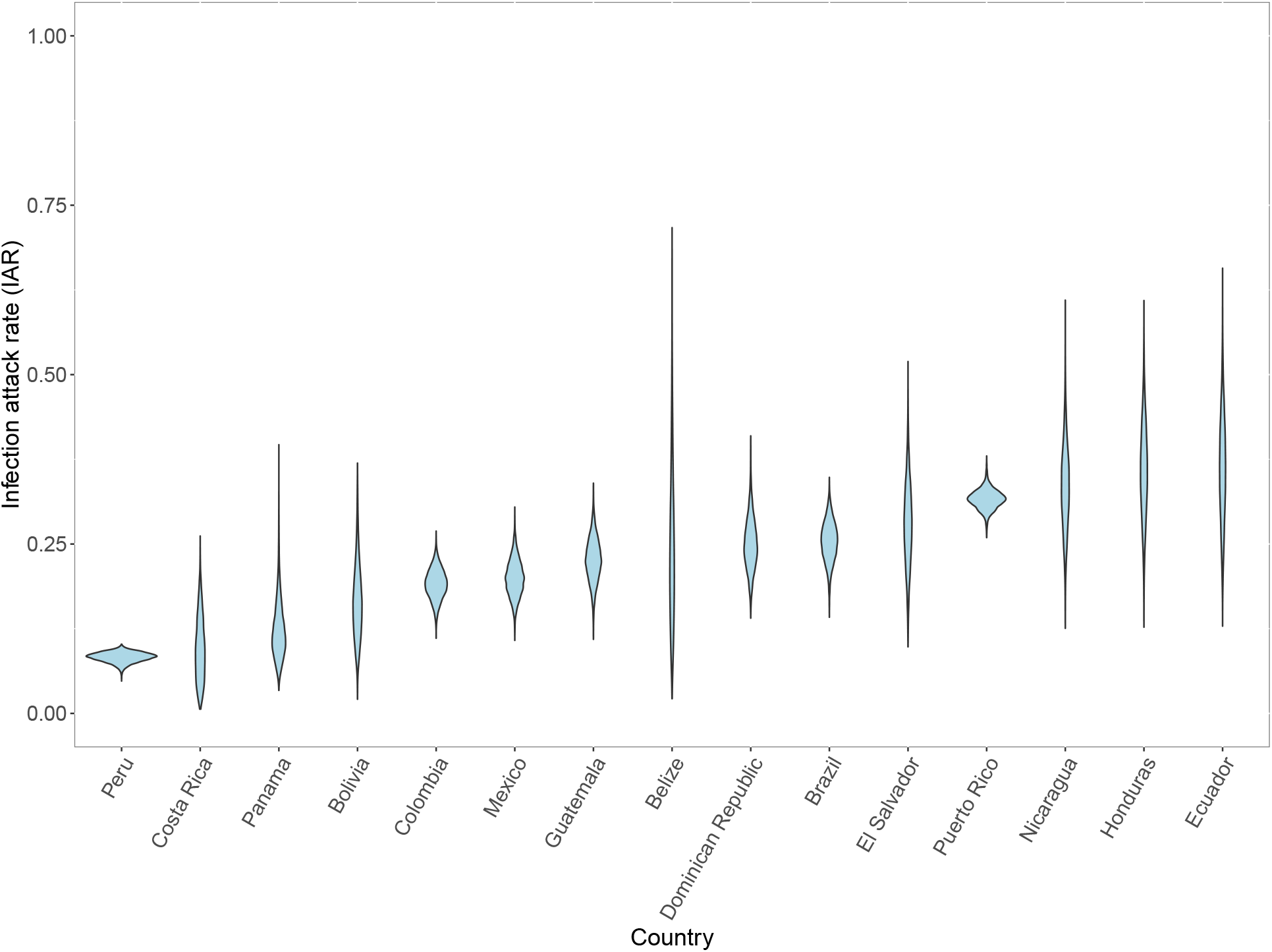
Posterior distribution of national ZIKV infection attack rate (IAR) for 15 different countries and territories.

**Figure 3:**
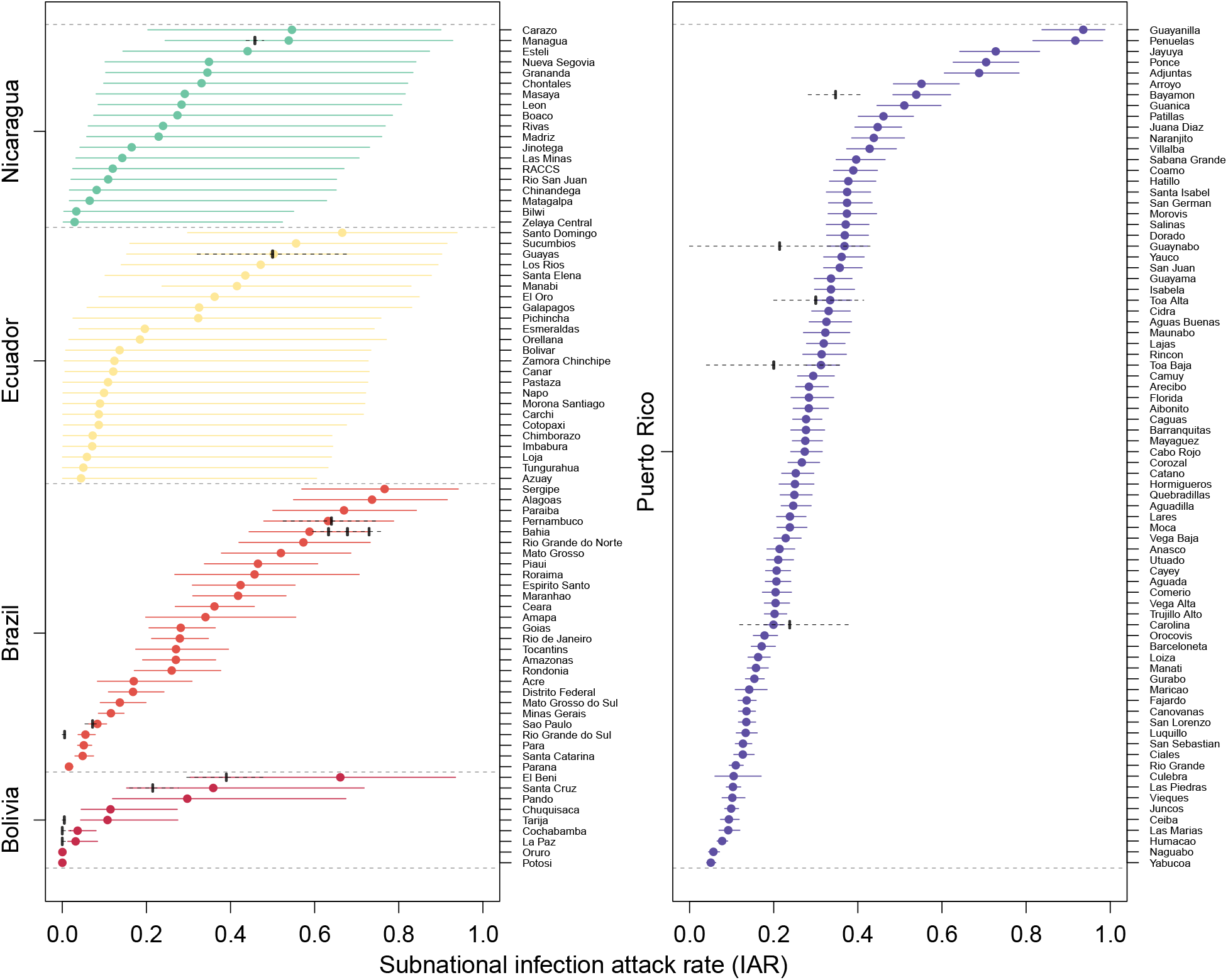
Posterior distribution of subnational ZIKV infection attack rates (IAR) for five different territories (Bolivia, Brazil, Ecuador, Nicaragua, and Puerto Rico). Colored circles and whiskers are the median and 95% credible intervals for each administrative unit. Black circles with dashed lines are seroprevalence estimates from the literature (see SI Table 6). The dashed lines are the 95% confidence intervals for the seroprevalence estimates assuming a binomial distribution with the exception of the 95% CI estimate from Rodriguez-Barraquer et al. (2019) for Bahia, Brazil which was taken directly from their analysis.

Mean estimated IARs in countries and territories where subnational data were not available ranged from a low of 0.003 (95% CrI: 0.000-0.013) in Uruguay to a high of 0.979 (95% CrI: 0.591-1.000) in Saint Martin and 0.979 (95% CrI: 0.616-1.000) in Saint Barthélemy (SI Table 4, SI Figures 22-25). Summing these IAR estimates across all countries and territories in the Americas, there were a total of 132, 278, 856 (95% CrI: 111, 305, 999-170, 156, 510) ZIKV infections across Latin America and the Caribbean. The majority of these infections, 114, 112, 765 (95% CrI: 99, 394, 311-128, 358, 749), were from the 15 modeled countries and territories, while the other 33 countries and territories accounted for an additional 16, 160, 273 (95% CrI: 5, 426, 679-51, 709, 310) infections. There were an estimated 53, 388, 206 (95% CrI: 40, 819, 653-64, 922, 428) ZIKV infections in Brazil and 25, 599, 729 (95% CrI: 19, 237, 945-32, 690, 560) in Mexico. Venezuela had the largest number of ZIKV infections (6, 633, 999; 95% CrI: 350, 008.3-31, 518, 000) out of the countries that were not explicitly modeled (SI Table 4).

### Parameter estimates

The median probability of a ZIKV infection being symptomatic across all countries was 0.15 (95% CrI: 0.02-0.55), which was lower than the prior estimate of 0.41 (95% CrI: 0.14–0.73). The estimated symptomatic probability ranged from 0.03 (95% CrI: 0.01-0.17) in Guatemala to 0.33 (95% CrI: 0.15-0.6) in Colombia (Figure 4A). This shift in the posterior estimate relative to the prior estimate of the symptomatic rate may be the result of identifiability issues between the symptomatic rate and the reporting rate parameters for the different data types. Misdiagnosis of symptomatic ZIKV infections as a different disease, such as dengue fever, could also lower the estimate of the symptomatic rate.

**Figure 4:**
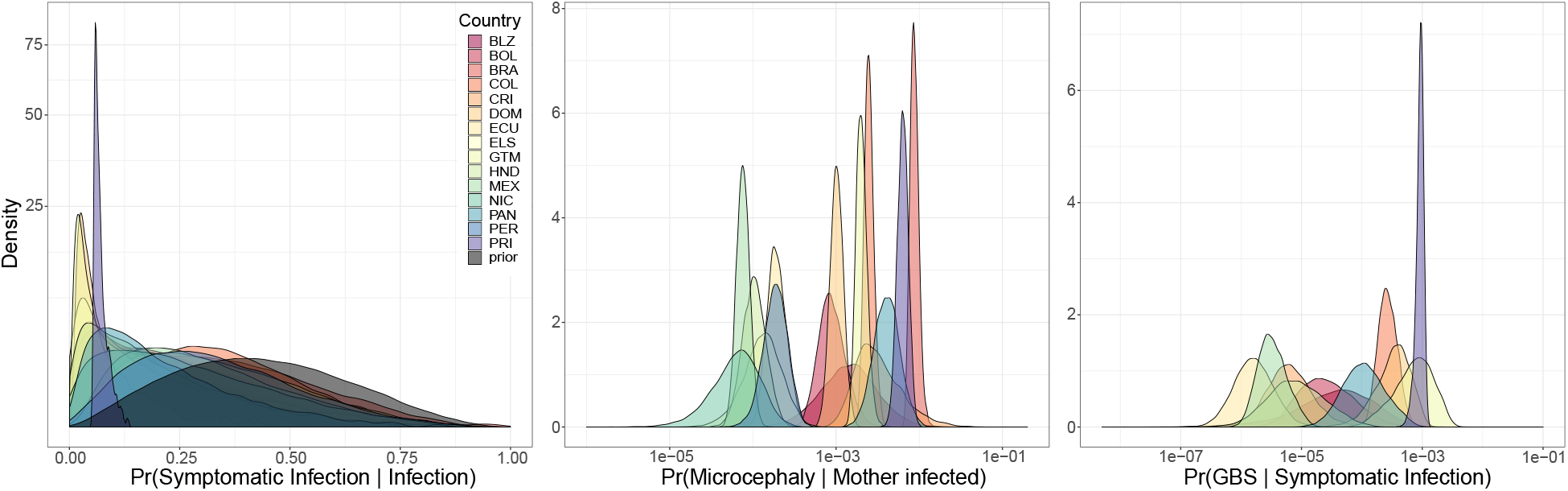
(A) Posterior and prior symptomatic probability estimates for each country or territory. (B) Posterior estimates from each country and territory of the probability that a ZIKV infection in a pregnant woman results in a reported case of microcephaly. (C) Posterior estimates from each country and territory of the probability that a symptomatic infection results in a reported Guillan-Barré syndrome (GBS) case.

Reported rates of microcephaly in every country and territory were lower than recent estimates of the risk of microcephaly based on studies of ZIKV infection during pregnancy (Figure 4B). The probability that an infection in a pregnant woman resulted in a reported case of microcephaly ranged from a low of 0.07 per 1,000 infections (95% CrI: 0.01-0.19) in Nicaragua to 8.7 per 1,000 infections (95% CrI: 7.13-11.39) in Brazil. The probability that a symptomatic infection would result in a reported Guillan-Barré case varied from a low of 0.16 per 100,000 symptomatic infections (95% CrI: 0.04-0.7) in Ecuador to a high of 93.06 per 100,000 symptomatic infections (95% CrI: 63.22-115.2) in Puerto Rico (Figure 4C). The large variability between countries in reporting rates for microcephaly and GBS cases could be due to differences in case definitions among countries, differences in surveillance, or differences in underlying dengue immunity that may have impacted the severity of ZIKV infections due to some form of cross-reactive response (Bardina et al., 2017; Andrade and Harris, 2018; Rodriguez-Barraquer et al., 2019).

The probability of a symptomatic infection being reported as a suspected or confirmed Zika case was higher for pregnant women than the general population for all countries where separate data on pregnant women were available, with the exception of suspected cases in El Salvador and Honduras (SI Figures 26-29). The countrywide reporting rate in pregnant women was as low as 0.002 (95% CrI: 0-0.012) for confirmed cases in El Salvador (SI Figure 29), and as high as 0.289 (95% CrI: 0.041-0.989) for confirmed cases in Costa Rica and 0.276 (95% CrI: 0.056-0.727) for suspected cases in Dominican Republic (SI Figure 28). As with the variability in reporting rates for GBS and microcephaly, this variability in confirming ZIKV infections in pregnant women indicates that there were considerable differences in surveillance and testing efforts among countries during the epidemic.

Countrywide reporting of suspected cases in the entire population ranged from 0.012 (95% CrI: 0.006-0.041) in Peru to 0.933 (95% CrI: 0.632-0.999) in Puerto Rico (SI Figure 26). Countrywide reporting of confirmed cases ranged from 0.0004 (95% CrI: 0.0001-0.0023) in El Salvador to 0.504 (95% CrI: 0.341-0.54) in Puerto Rico (SI Figure 26). The variation in reporting rates among administrative units within a country or territory was largest in Colombia for suspected cases (70.2% of variance was between administrative units vs. 29.8% due to within-unit variance), suspected cases in pregnant women (63.8%), and confirmed cases in pregnant women (67.7%). The largest between-administrative unit variance in reporting rates for confirmed cases in the total population occurred in Puerto Rico (61.4% of total variance). Several countries showed little variability in reporting rates among administrative units, with *<* 1% of total variance explained (SI Figures 30-33).

## Model validation

### Posterior predictive checks

Our model did not generate predicted cases that were more extreme than the numbers of observed cases—as indicated by Bayesian p-values between 0.1 and 0.9—for each observed data type in all countries and territories except Puerto Rico. For Puerto Rico, the total number of reported confirmed cases (*p*_*B*_ = 0.990), suspected cases (*p*_*B*_ = 0.999), GBS cases (*p*_*B*_ = 0.004), and infections in blood donors (*p*_*B*_ = 0.020) were outside the range of model predictions, but the number of microcephaly cases was not (*p*_*B*_ = 0.566) (Figure 5). The underestimation of confirmed and suspected cases—and overestimation of GBS cases and infections in blood donors—in Puerto Rico indicates that our model was not capable of reconciling estimates based on different data types when the subnational reporting rates for these different data types did not vary consistently. Therefore, the municipality-level IAR estimates for Puerto Rico should be viewed with caution. The observed number of microcephaly cases, confirmed cases in pregnant women, and suspected Zika cases in pregnant women fell within the 95% posterior predictive interval (PPI) of the model for every country and territory that reported these data types (Figure 5). Beyond Puerto Rico, the number of confirmed cases in Guatemala was higher than the 95% PPI (1,032 vs. 811-963), the number of suspected cases in Honduras was higher than the 95% PPI (32,385 vs. 29,659-30,583), and the number of suspected cases in Brazil was lower than the 95% PPI (231,725 vs. 240,240-243,040). Comparisons of the observed data to subnational PPIs for each country or territory are provided in the project Github repository https://github.com/mooresea/Zika_IAR.

**Figure 5:**
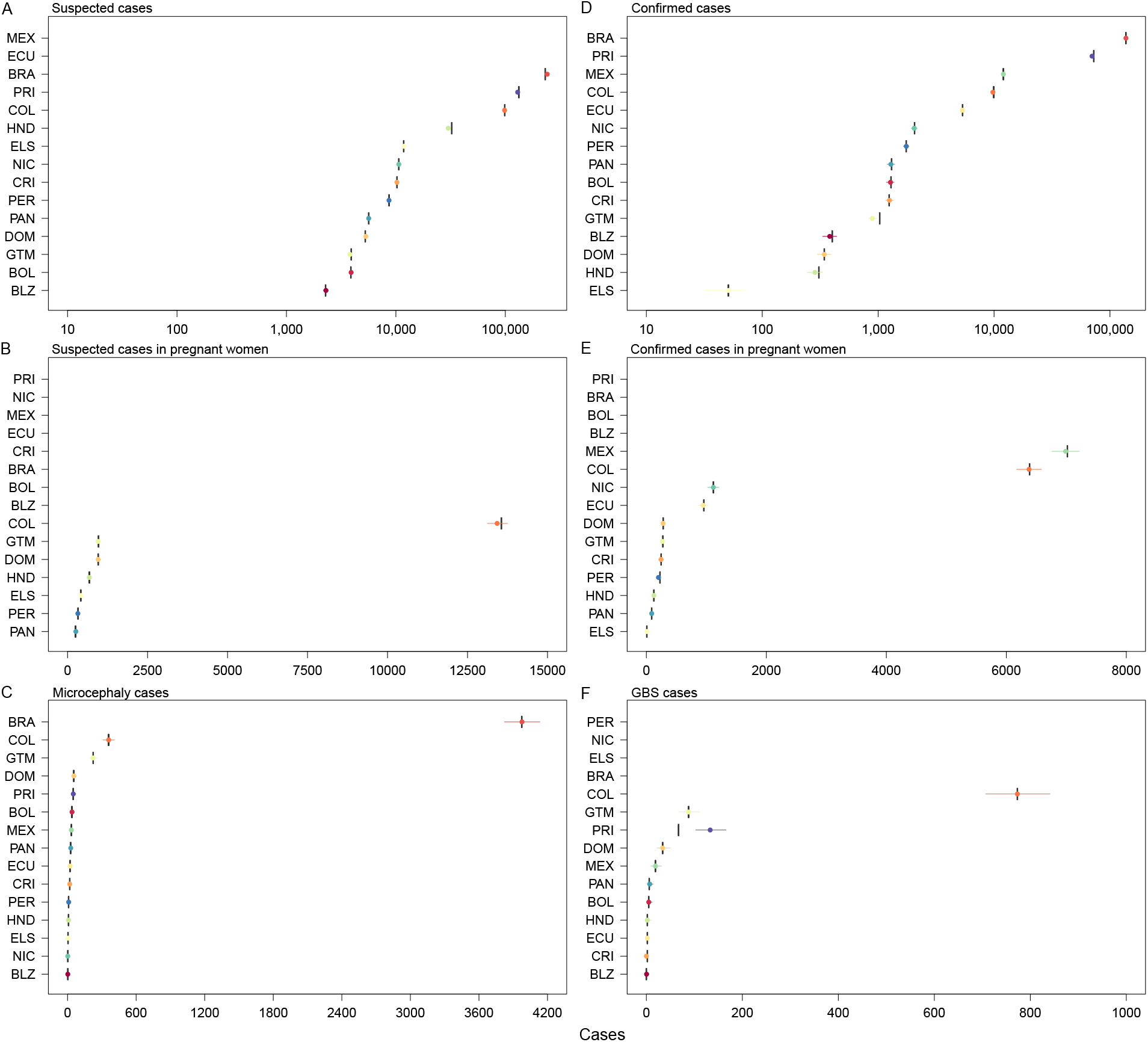
Posterior predictive checks at the national level for each data type used in the Bayesian models. Vertical lines are the observed cases and circles are the predicted number of cases with 95% credible intervals for each country and data type.

### Holdout analysis

Modeled IAR estimates were not overly sensitive to any one data type. Removing any one data type from the estimation process did not significantly alter the national IAR estimate for Guatemala, with median estimates within 1.0% of the median estimate from the full model of 22.6% (SI Figure 34). For the Dominican Republic, the median national IAR estimate varied from 0.217 to 0.314 depending on which data type was withheld, but these median model estimates were all within the 95% CrI for the full model of 0.180-0.331 (SI Figure 35). For Panama, the national median estimates with different data types withheld were all slightly higher (0.127-0.150) than the median estimate of the full model (0.117), but still within the 95% CrI of 0.060-0.222 (SI Figure 36). When models were fitted with a single data type, national and subnational IAR estimates varied widely depending on which data type was used to fit the model (SI Figures 37-39). The single-data-type models overestimated the median national IAR relative to the full model for three countries or territories (Guatemala, Panama, and Puerto Rico), and had increased uncertainty at both the national and subnational levels.

### Comparison to seroprevalence studies

Our median IAR estimates for Managua, Nicaragua (0.538; 95% CrI: 0.244-0.928) and Guayas, Ecuador (0.503 ; 95% CrI: 0.153-0.902) were close to seroprevalence estimates from those regions that were withheld from the fitting process (Figure 3). Overall, the 95% CI for 15 of 18 seroprevalence estimates from published studies overlapped with the 95% CrI of the IAR estimates from our model for the corresponding administrative unit. These two measures are not necessarily identical for every country or territory, because many of the seroprevalence studies were either conducted in only part of the administrative unit for which we estimated the IAR (e.g., the city of Salvador within Bahia State, Brazil or the city of Santos within São Paulo State, Brazil) or were conducted well before the end of the epidemic (Bolivia). The three estimates that did not overlap were Tarija, Bolivia (seroprevalence: 0.005; 95% CI: 0.000-0.015 vs. IAR: 0.108; 95% CrI: 0.043-0.275), Rio Grande do Sul, Brazil (seroprevalence: 0.006; 95% CI: 0.000-0.016 vs. IAR: 0.055; 95% CrI: 0.038-0.078), and Bayamon, Puerto Rico (seroprevalence: 0.347; 95% CI: 0.292-0.412 vs. IAR: 0.539; 95% CrI: 0.483-0.620).

## Discussion

Our results show that the vast majority of ZIKV infections in the Americas went unreported during the recent epidemic. PAHO reported fewer than one million suspected or confirmed Zika cases, whereas we estimate that 132, 278, 856 (95% CrI: 111, 305, 999-170, 156, 510) people were infected. This discrepancy is due to the high number of asymptomatic infections, as well as estimated reporting rates below 1% for both suspected and confirmed Zika cases in many countries. The majority of ZIKV infections (114, 112, 765; 95% CrI: 99, 394, 311-128, 358, 749) occurred in the fifteen countries and territories that we modeled subnationally, while an additional 16, 160, 273 (95% CrI: 5, 426, 679-51, 709, 310) occurred in the remaining American territories where national IAR estimates were extrapolated from the estimated reporting rates from the modeled countries. These estimates of the national and subnational IAR and total ZIKV infections represent the most extensive attempt to date to estimate the size of the Zika epidemic in the Americas.

Our results show significant within- and between-country variability in IARs. This large variability in IARs could reflect spatial heterogeneity in habitat suitability for *Ae. aegypti* (Kraemer et al., 2015). Early projections of the epidemic also predicted that the IAR would be highly heterogeneous due to the geographic distribution of *Ae. aegypti* as well as other factors likely to influence transmission intensity, such as temperature and economic prosperity (Perkins et al., 2016). The historical pattern of virus introduction and spread may have also contributed to the high level of within- and between-country variability in IAR (Zhang et al., 2017). Local outbreaks in geographic regions or municipalities that began near the end of the local transmission season may have ended before epidemic fadeout would be expected due to herd immunity (Ferguson et al., 2016; Huber et al., 2018). Determining the extent to which this spatial heterogeneity is the result of ZIKV epidemiology versus historical contingencies will be important for identifying current and future at-risk populations.

Although our IAR estimates closely matched estimates from seroprevalence studies in some areas (e.g., Managua, Nicaragua, Guayas, Ecuador, and Pernambuco, Brazil), other estimates diverged from published seroprevalence estimates. One potential reason for the discrepancy between our IAR estimates and serosurvey-based estimates is a mismatch in the spatial scale of the region that the IAR estimate covers. For example, our estimate for the Brazilian state of Bahia (0.514; 95% CrI: 0.388-0.631), was slightly lower than the seroprevalence estimates of 0.63 (95% CI: 0.595-0.671),0.68 (95% CI: 0.600-0.744), and 0.73 (95% CI: 0.70–0.76) from Salvador, the largest city in Bahia (Netto et al., 2017; Moreira-Soto et al., 2018; Rodriguez-Barraquer et al., 2019). However, the overall IAR in Bahia might be lower than the IAR in Salvador if rural areas outside of Salvador experienced a lower infection attack rate. Serosurvey-based estimates might also underestimate IAR if the sensitivity and specificity of the serological assay are not taken into account. Only Rodriguez-Barraquer et al. (2019) reported seroprevalence estimates adjusted for assay sensitivity and specificity, which increased their estimate from 0.63 (95% CI: 0.60-0.65) to 0.73 (95% CI: 0.70-0.76) for Salvador, Brazil. Another potential cause of discrepancies is that several of the serosurveys were conducted prior to completion of the ZIKV epidemic. For example, the published seroprevalence estimates for Bolivia from Villarroel et al. (2018) were conducted from December, 2016 until May, 2017, but Bolivia had significant ZIKV transmission activity throughout 2017 and 2018. As a result, our IAR estimates which incorporated data from 2017 and 2018 were higher than those reported by Villarroel et al. (2018). Finally, posterior predictive checks showed that Puerto Rico was the only territory for which our model did not accurately predict several of the observed data types, which may explain why our IAR estimate for Bayamon, Puerto Rico was significantly different than the seroprevalence estimate from Lozier et al. (2018). Puerto Rico has more administrative level-one areas (78) than the other modeled countries and territories, which suggests that our model may produce overly precise estimates when there is a large sample size and variation in the reporting rates of different case types that is not spatially consistent. Conducting further serosurveys in a range of locations across the Americas would be an important step in determining current levels of immunity and would further validate our model estimates.

The estimated reporting rates for microcephaly in each territory were significantly lower than the reported risk of microcephaly or CZS during pregnancy from several different studies. Our microcephaly detection estimates are significantly lower than estimates from epidemiological studies because we are estimating both the probability of a ZIKV infection resulting in microcephaly, and the probability that a microcephaly case was diagnosed and reported. Our estimates also depended on estimates of the annual number of births in a region, but this could represent an overestimate of the number of births at risk in many regions because the duration of the epidemic in most localities was less than one year. Overestimating the number of births at risk would lead to an underestimate of the microcephaly rate in a region or country. Whether this underestimate in the microcephaly rate would also affect the estimate of IAR would depend on the influence of the other data types on the total likelihood. The risk of microcephaly or CZS has been estimated as 0.95% (95% CI: 0.34-1.91%) during the first trimester in French Polynesia (Cauchemez et al., 2016), 2.3% (2 out of 86 births) in Colombia (Rodriguez-Morales et al., 2018), and 5% (95% CI: 4-7%) among fetuses and infants born to women with laboratory-confirmed ZIKV infection in the U.S. territories (Shapiro-Mendoza, 2017). In a cohort study in Rio de Janerio, 42% of children born to ZIKV-infected mothers were found to have central nervous system abnormalities, although only 4 of 117 were diagnosed with microcephaly (Brasil et al., 2016). Our estimates ranged from 0.1 per 1,000 infections (95% CrI: 0-0.2) in Nicaragua to 8.7 per 1,000 infections (95% CrI: 7.1-11.4) in Brazil. Changes in the reporting rates of both ZIKV infections and microcephaly over the course of the epidemic could also complicate the estimation of the true risk of CZS (Hay et al., 2018).

Rates of GBS also varied significantly, ranging from 0.04 to 8.51 per 100,000 total ZIKV infections. A recent review estimated a rate of 20 GBS cases per 100,000 ZIKV infections (95% CrI: 5-45 per 100,000), which is higher than our estimates (Mier-y Teran-Romero et al., 2018). However, that analysis also estimated the IAR for each location included in their study and estimated lower IARs for Bahia, Brazil (0.02), Salvador, Brazil (0.08), Colombia (0.09), El Salvador (0.15), and Honduras (0.04) than our model. Their IAR estimates for Bahia State and Salvador in Brazil were also much lower than seroprevalence estimates from three studies (Netto et al., 2017; Moreira-Soto et al., 2018; Rodriguez-Barraquer et al., 2019) that more closely matched our IAR estimates for Bahia State. Underestimating the number of ZIKV infections would lead to an overestimate of GBS incidence per infection. For Puerto Rico, where our estimate for IAR (0.316; 95% CrI: 0.288 − 0.345) is within the 95% CrI of the IAR estimate (0.17; 0.08-0.46) by Mier-y Teran-Romero et al. (2018), our estimate of 8.51 (95% CrI: 6.94-11.15) GBS cases per 100,000 ZIKV infections is also within the range of their estimate of 14 (95% CrI: 4-25) GBS cases per 100,000 ZIKV infections. This overlap suggests that our method of simultaneously estimating IAR, reporting rates, and the risk of GBS produces reasonable estimates for GBS risk and reporting.

The current status of population-level immunity throughout the Americas has important implications for the future of Zika epidemiology in the region (Ferguson et al., 2016; Lessler et al., 2016; Perkins, 2017). Our IAR estimates suggest that a number of areas have high levels of herd immunity, which has also been suggested by other studies (O’Reilly et al., 2018). However, the high degree of heterogeneity in subnational IARs suggests that certain areas in Central and South America that are suitable for ZIKV transmission may still contain a considerable number of susceptible individuals. While some areas experienced a low IAR because of environmental conditions that limited local transmission, such as high-elevation locations in Colombia and Peru, there are other areas that had a low IAR where conditions appear favorable for ZIKV transmission. For example, the Brazilian states of Pará and Amazonas had median IAR estimates of 0.077 and 0.164, even though both were believed to be high-risk locations (Messina et al., 2016; Perkins et al., 2016) based on high suitability for *Ae. aegypti* (Brady et al., 2014; Kraemer et al., 2015) and historical dengue incidence (Bhatt et al., 2013; Siqueira et al., 2005). If the remaining population-at-risk is large in these areas, then local outbreaks in the near future could still be possible. Modeling studies also suggest that the presence of multiple locations with incomplete herd immunity increases the chances that ZIKV becomes endemic in the region in the next decade as new births replenish the susceptible population (Ferguson et al., 2016). Although Zika incidence in the Americas in 2019 appears to be much lower than during the epidemic, limited transmission has been reported in multiple countries and recent Zika outbreaks have also been reported in Africa and Asia (ProMED-mail, 2019), leaving open the possibility that ZIKV could be re-introduced into vulnerable populations. However, it is also possible that there was significant transmission in these states prior to the detection of ZIKV transmission in the Americas, in which case the actual IAR could be higher than we estimate. Genomic data suggests that ZIKV was present in northeast Brazil by early 2014 and spread to multiple countries before Zika surveillance began (Faria et al., 2017). Microcephaly data can re-construct some of this missed portion of the epidemic due to the lag between maternal infection and birth, but even this data will miss infections that occurred more than a year prior to the first case reports.

Because of the high variation in estimated reporting rates among modeled countries, there was a high degree of uncertainty in our projections of IAR in the non-modeled territories. Previous studies have also found high variation in the probability of a ZIKV infection being reported between countries (O’Reilly et al., 2018). This resulted in a fair amount of uncertainty in our estimates of the total number of ZIKV infections (95% CrI: 111, 305, 999-170, 156, 510). However, because most of the non-modeled territories were relatively small in population size (the total estimated population size in the 15 modeled territories is 507.1 million versus 116.6 million in the 33 non-modeled territories) the median estimate is largely a reflection of infections in the modeled countries. Because we were unable to precisely estimate IAR without estimating territory-specific reporting rates, the publication of subnational totals for different types of Zika cases (suspected, microcephaly, etc.) would help greatly in refining these estimates. The publication of this data in conjunction with seroprevalence data would be particularly valuable for examining differences in surveillance systems and the geographic variation in IAR throughout the entire region (Ferguson et al., 2016).

The Zika epidemic in the Americas was a major public health concern that drew considerable attention from both the public health sector and the general public. Despite this attention, the majority of infections were not detected by public health surveillance systems. Using a hierarchical Bayesian model with empirically-informed priors, we were able to leverage Zika case reporting to simultaneously estimate national and subnational reporting rates, the fraction of symptomatic infections, and subnational IARs. Our results indicate that fewer than 1% of ZIKV infections in the Americas were reported during the recent epidemic. In the absence of detailed seroprevalence data, our subnational and national IAR estimates are an important first attempt at assessing current levels of ZIKV immunity throughout the Americas. Current levels of herd immunity are a critical determinant of the probability of further Zika outbreaks—and potentially the likelihood of dengue outbreaks as well—in this region in the next decade.

## Data Availability

All data used in our analysis, along with model code, is provided on our project github: https://github.com/mooresea/Zika_IAR.
In addition, all of the reports from which the data were collated are publicly available from the various government agencies that released the reports (see References).

## Acknowledgements

We would like to thank the Epidemiology Unit of the Belize Ministry of Health for providing Zika reports upon request. We also thank all of the governments in North and South America that made their Zika surveillance reports or data publicly available online.

## Supplementary Information

### 1 Zika data

We collected publicly available Zika data for the 15 countries or territories where we were able to find subnational case counts for at least one data type (SI Table 1). The potential data types considered were confirmed Zika cases (either lab or clinically confirmed), suspected Zika cases, microcephaly cases associated with a ZIKV infection in the mother, Zika-associated cases of Guillan-Barré syndrome (GBS), and the proportion of blood donors with a positive ZIKV infection (Puerto Rico only). Many territories also reported the number of suspected or confirmed Zika cases in pregnant women. Where available, these counts were treated as independent observations from the population-wide observations. Although cases in pregnant women and in the entire population are treated as independent data points, we assume that the infection attack rate (IAR) is the same in both populations. A graphical representation of how IAR was estimated from the observed data is provided in Figure (1) of the main text. Short descriptions for each of the variables and parameters included in the model are presented in SI Table (2).

Where available, we obtained Zika data at the first administrative level (e.g., province or state) within a country or territory. Where lower level data was available, it was aggregated to the first administrative level. The majority of datasets were obtained either from governmental websites or from country reports produced by PAHO (Pan American Health Organization (PAHO), 2017a). Details on the time period covered by each dataset and data sources are provided in SI Table (1). Where data totals were provided by epidemiological week, the start and end dates for these reporting periods were obtained from the governmental report or assumed to match the dates of WHO epidemiological weeks. Although we list the start and end periods of reporting for each data type, in some instances there may have been gaps in reporting during this period.

**Supplementary Table 1:**
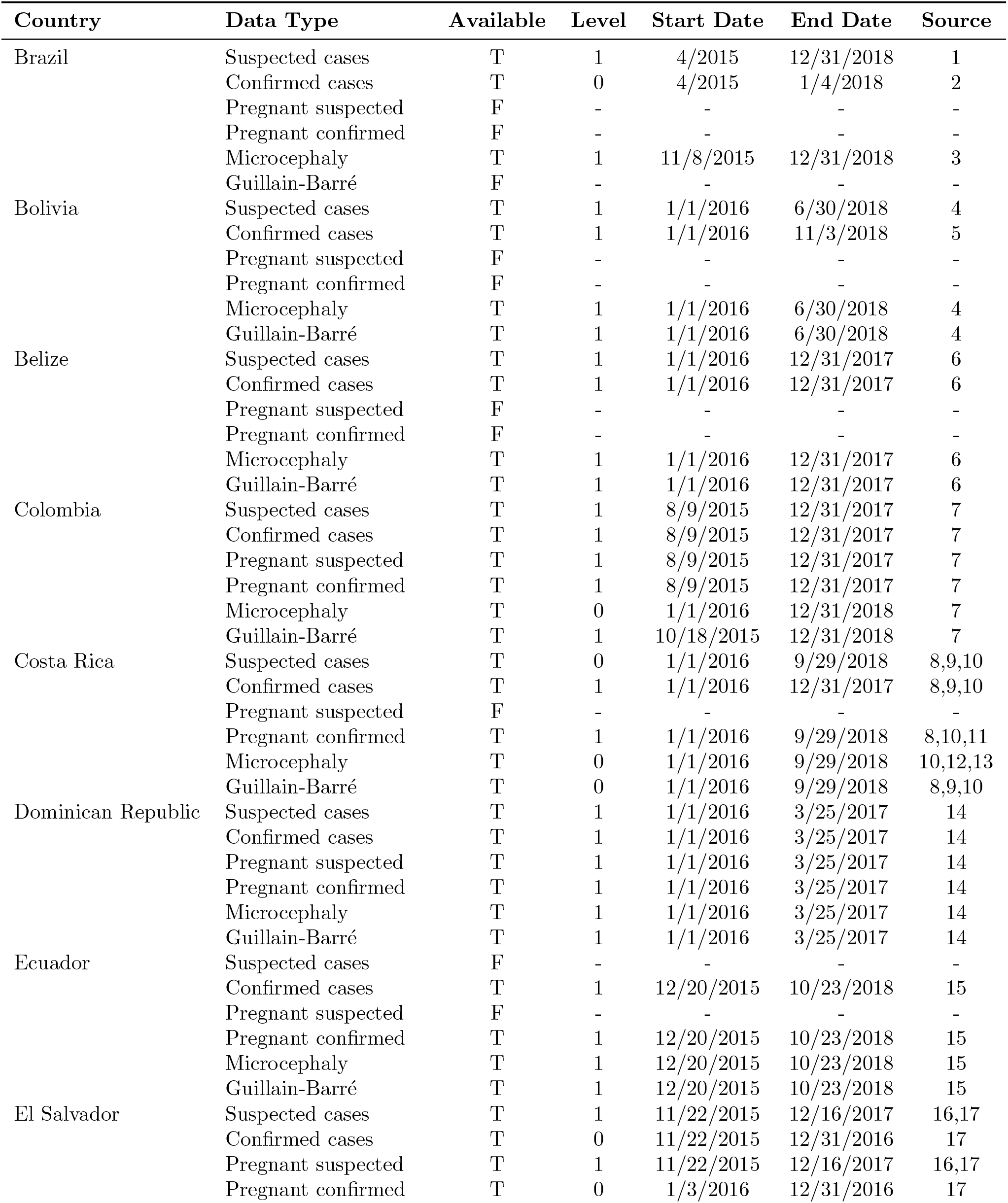
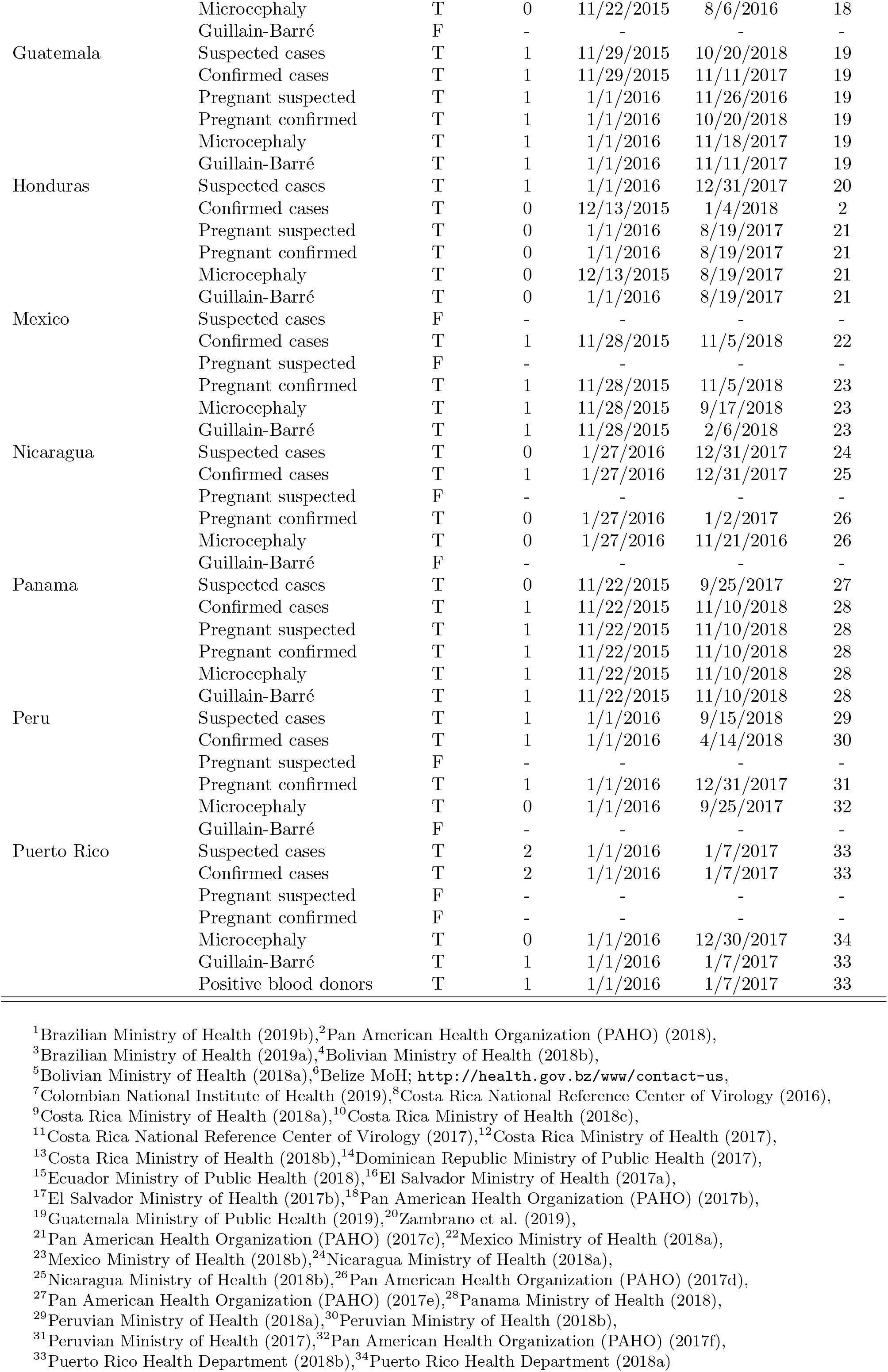
Summary of data availability at the national or subnational level for each country and territory.

**Supplementary Table 2:** Description of each of the variables and parameters used in the models.

| Variable or Parameter | Description |
| --- | --- |
| $\mathcal{I}$ | Number of infected individuals |
| $\mathcal{Z}$ | Number of symptomatic infections |
| $S_T$ | Number of suspected cases in total population |
| $C_T$ | Number of confirmed cases in total population |
| $S_P$ | Number of suspected cases in pregnant women |
| $C_P$ | Number of confirmed cases in pregnant women |
| $G$ | Number of Guillan-Barré syndrome (GBS) cases |
| $M$ | Number of microcephaly cases |
| $D$ | Number of blood donors |
| $I$ | Number of infections in blood donors |
| $\rho_Z$ | Proportion of infections that are symptomatic |
| $\rho_{S_T}$ | Fraction of symptomatic infections that are reported as a suspected case |
| $\rho_{C_T}$ | Fraction of symptomatic infections reported as a confirmed case |
| $\rho_{S_P}$ | Fraction of symptomatic infections in pregnant women that are reported as a suspected case |
| $\rho_{C_P}$ | Fraction of symptomatic infections in pregnant women that are reported as a confirmed case |
| $\rho_G$ | Fraction of symptomatic infections that result in a reported GBS case |
| $\rho_M$ | Fraction of infections in pregnant women that result in a reported microcephaly case |
| $\rho_I$ | Correction factor for fraction of blood donors that are infected at donation based on length of viremia and asymptomatic rate |
| $\alpha_{S_T}$ | Hyperparameter from beta distribution for $\rho_{S_T}$ |
| $\beta_{S_T}$ | Hyperparameter from beta distribution for $\rho_{S_T}$ |
| $\alpha_{S_T}$ | Hyperparameter from beta distribution for $\rho_{C_T}$ |
| $\beta_{S_T}$ | Hyperparameter from beta distribution for $\rho_{C_T}$ |
| $\alpha_{S_P}$ | Hyperparameter from beta distribution for $\rho_{S_P}$ |
| $\beta_{S_P}$ | Hyperparameter from beta distribution for $\rho_{S_P}$ |
| $\alpha_{C_P}$ | Hyperparameter from beta distribution for $\rho_{C_P}$ |
| $\beta_{C_P}$ | Hyperparameter from beta distribution for $\rho_{C_P}$ |

### 2 Blood donor infections

To prevent transfusion-associated transmission of ZIKV, Puerto Rico began screening all blood donations for ZIKV in April, 2016 (Chevalier et al., 2017). Because this method has the ability to detect asymptomatic infections in contrast to most of the other surveillance methods included in our analysis, we used this data to estimate IAR in Puerto Rico. This data was available at the regional level (n=8) through January 7, 2017 from Puerto Rico Health Department (2018b). The number of blood donors with an active ZIKV infection, *I*_*i*_, within a population of screened blood donors *D*_*i*_, was related to the IAR by the parameter, *ρ*_*I*_, which corrects for the fact that blood donation is only possible when an individual is asymptomatic and adjusts for the mean duration of viremia in asymptomatic individuals (including pre-symptomatic individuals) relative to the duration of the collection period. As a result, the number of infections among blood donors is assumed to follow a Poisson distribution, 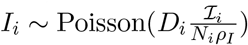.

We followed the methodology of Chevalier et al. (2017) in estimating a prior distribution for the incidence correction factor, *ρ*_*I*_, with a few modifications to reflect the timeframe of our study and additional data available since their study was published. To account for the transient nature of viremia in an infected individual, the number of observed infections in blood donors was scaled by the probability that a blood donor would have a detectable viremia at the time of donation. The duration of collection (*d* = 41 weeks) is therefore divided by the mean duration of ZIKV viremia 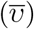, resulting in a crude scaling factor of 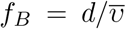 (Busch et al., 2006). This scaling factor is biased because symptomatic individuals are removed during the screening process, meaning that only people with asymptomatic or pre-symptomatic infections will donate blood. The unbiased scaling factor assuming only asymptomatic or pre-symptomatic individuals donate is,

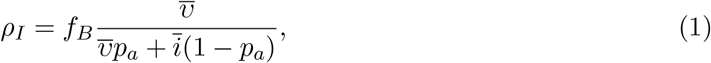

where *p*_*a*_ is the probability an infection is asymptomatic and 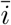 is the mean incubation period. The prior distribution for *ρ*_*I*_ was estimated using the distributions for the incubation period and viremic period described in Chevalier et al. (2017), and the probability of asymptomatic infection in Puerto Rico from Mitchell et al. (2019) and fit to a gamma distribution using the ‘fitdistr’ package in R, resulting in a Gamma(15.97, 0.42) prior.

### 3 Model implementation and diagnostics

Each country or territory model was fitted using the ‘rstan’ version 2.18.2 package in R (Stan Development Team, 2018) using the ‘No-U-turn’ sampling algorithm with four chains of 5,000 iterations and a 50% burn-in period. Smaller step sizes for the sampling algorithm were set by increasing the adapt delta parameter from the default of 0.8 to 0.99. In addition, the maximum tree depth was increased from 10 to 15. Convergence was assessed using the Gelman-Rubin convergence diagnostic, *R*_*c*_ (Gelman et al., 2013). The *R*_*c*_ values for every parameter in each of the 15 county or territory models were *<* 1.05, indicating that each model achieved convergence. Traceplots of the log of the posterior distribution are provided in SI Figure (1).

**Supplementary Figure 1:**
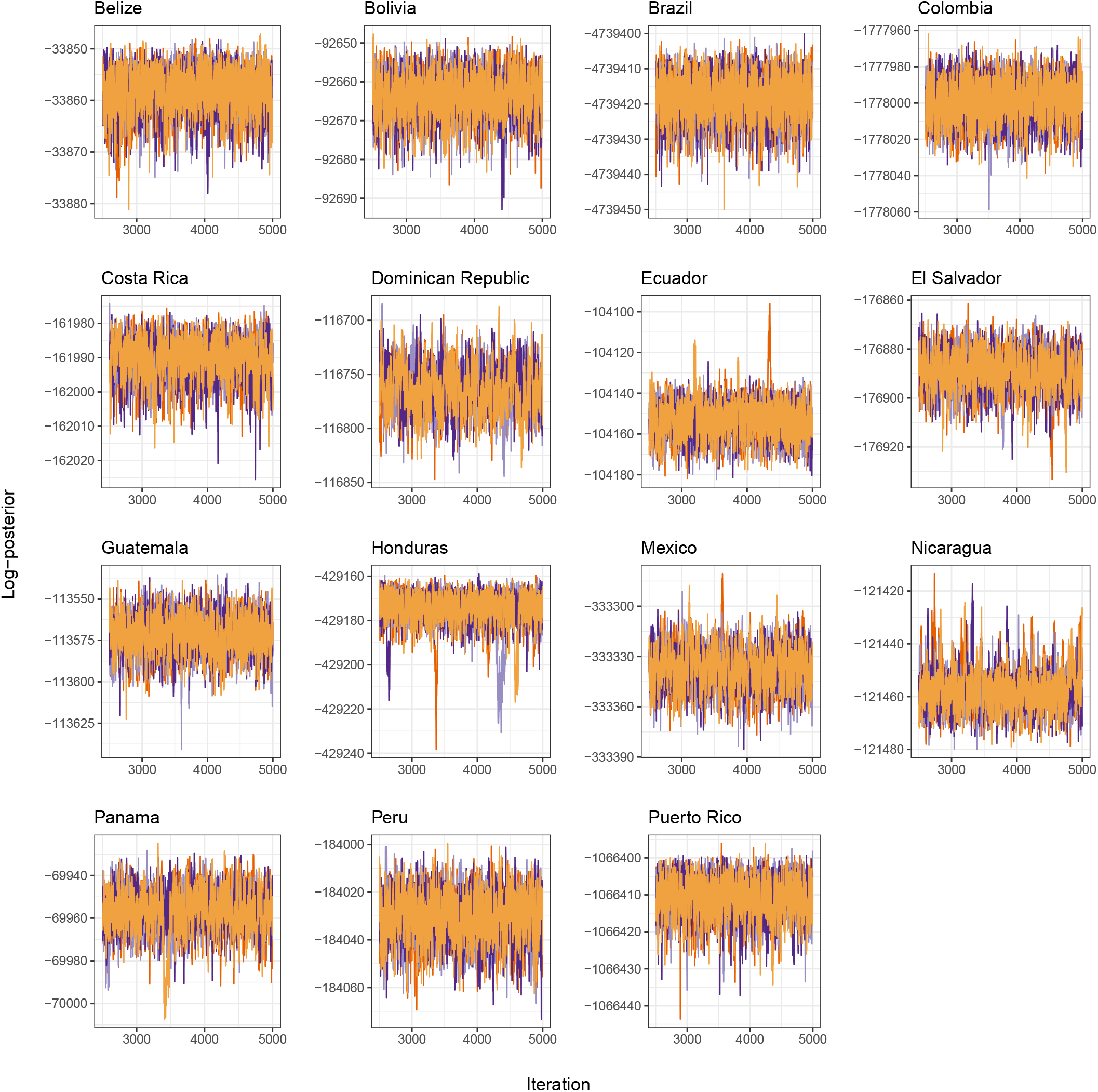
Traceplots of the log probability density of the posterior distribution for each country model after warmup. Colors represent the four separate chains.

### 4 Testing model assumptions

In the analysis presented in the main text, we used a Beta(1, 2) prior for the IAR in each country or territory model. The intention of this prior was to lightly constrain our ZIKV IAR estimates and prevent the model from converging towards extreme estimates without precluding the possibility of values anywhere between 0 and 1. This prior distribution has a median value of 0.292 (95% range: 0.013–0.842). To examine the sensitivity our IAR estimates to this prior assumption, we also ran a model version for each territory with a uniform prior on the IAR. With the uniform prior for the subnational IARs, the posterior IAR estimates at both the subnational and national level were higher for all 15 modeled countries and territories except for Costa Rica (SI Table 3; SI Figure 2).

In the analysis presented in the main text, we assumed that the reporting of symptomatic ZIKV infections, *Ƶ*_*i*_, as suspected cases, *S*_*i*_, followed a binomial distribution, 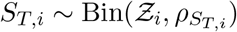. The probability of a symptomatic infection being reported as a suspected case, 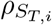, in administrative unit *i* of a country or territory followed a beta distribution with hyperparameters 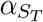 and 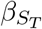. However, because there is considerable overlap between the symptoms of a ZIKV infection and the symptoms of several other arbovirus infections—including dengue and chikungunya—the number of suspected Zika cases could exceed the number of symptomatic ZIKV infections if other arbovirus infections were misdiagnosed as ZIKV during the epidemic. To account for this possibility, we also considered a model where 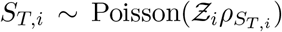. The reporting rate for suspected cases was allowed to range above one by drawing from a gamma distribution, 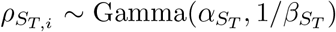. The gamma distribution hyperparameters, 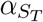 and 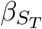, were assigned truncated standard normal prior distributions. These hyperparameter priors result in a mean of 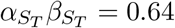, and a variance of 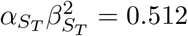 for the prior of the gamma distribution.

The version of the model with a Poisson distribution for suspected cases was run for Costa Rica, Guatemala, Panama, and Puerto Rico. These four countries and territories represented the range of estimated suspected reporting rates that were observed for the model with a binomial distribution, with Guatemala and Panama having relatively low estimates of 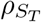, and Costa Rica and Puerto Rico having the second highest and highest estimates of 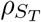, respectively. The values of 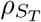 did not vary significantly between the models with binomial or Poisson distributions for Panama (0.092; 95% CrI: 0.024-0.436 vs. 0.074; 95% CrI: 0.021-0.343) or Guatemala (0.027; 95% CrI: 0.001-0.194 vs. 0.040; 95% CrI: 0.0022-0.268) (SI Figure 3). In addition, the IAR estimates for these two countries differed by *<*1% (SI Figure 4). The median estimate of 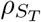 for Costa Rica was lower with the Poisson distribution (0.14; 95% CrI: 0.029 − 0.768) than with the binomial distribution (0.255; 95% CrI: 0.037 − 0.908) (SI Figure 3). This decrease in the estimated reporting rate was only associated with a small increase in the IAR estimate from 0.092 (95% CrI: 0.019 − 0.193) to 0.102 (95% CrI: 0.026 − 0.206) (SI Figures 4-5). Puerto Rico had the highest estimate of 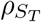 when suspected cases were binomially distributed, with a median of 0.933 and a upper 95% credible interval very close to 1 (95% CrI: 0.632 − 0.999). When we assumed suspected cases followed a Poisson distribution, the median estimate of 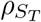 was 0.299 (95% CrI: 0.099−0.958). The marginal posterior distribution for 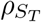 with a Poisson distribution was much broader than with the binomial distribution, and although the 95% credible interval was below 1 the upper estimates from the posterior were greater than 1 (SI Figure 3). The estimated territory-wide IAR in Puerto Rico was higher with a Poisson distribution (0.38; 95% CrI: 0.325 − 0.437) than with the binomial distribution (0.316; 95% CrI: 0.288 − 0.345) (SI Figure 4). A majority of Puerto Rico’s municipalities had higher IAR estimates with a Poisson distribution, but several estimates were lower than they were with a binomial distribution (SI Figure 6).

**Supplementary Table 3:**
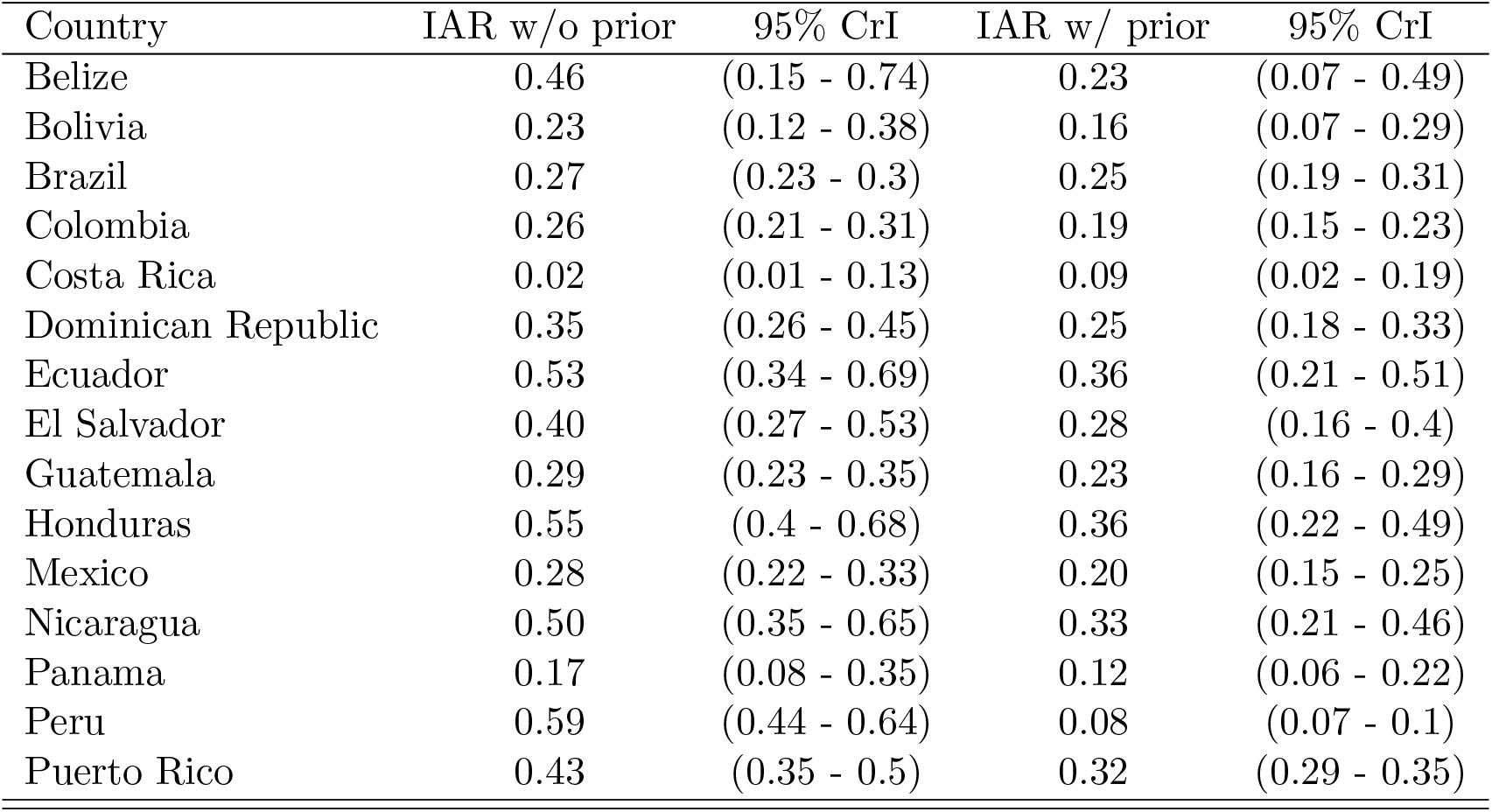
Comparison of national IAR estimates with or without a beta prior for IAR.

| Country | IAR w/o prior | 95% CrI | IAR w/ prior | 95% CrI |
| --- | --- | --- | --- | --- |
| Belize | 0.46 | (0.15 - 0.74) | 0.23 | (0.07 - 0.49) |
| Bolivia | 0.23 | (0.12 - 0.38) | 0.16 | (0.07 - 0.29) |
| Brazil | 0.27 | (0.23 - 0.3) | 0.25 | (0.19 - 0.31) |
| Colombia | 0.26 | (0.21 - 0.31) | 0.19 | (0.15 - 0.23) |
| Costa Rica | 0.02 | (0.01 - 0.13) | 0.09 | (0.02 - 0.19) |
| Dominican Republic | 0.35 | (0.26 - 0.45) | 0.25 | (0.18 - 0.33) |
| Ecuador | 0.53 | (0.34 - 0.69) | 0.36 | (0.21 - 0.51) |
| El Salvador | 0.40 | (0.27 - 0.53) | 0.28 | (0.16 - 0.4) |
| Guatemala | 0.29 | (0.23 - 0.35) | 0.23 | (0.16 - 0.29) |
| Honduras | 0.55 | (0.4 - 0.68) | 0.36 | (0.22 - 0.49) |
| Mexico | 0.28 | (0.22 - 0.33) | 0.20 | (0.15 - 0.25) |
| Nicaragua | 0.50 | (0.35 - 0.65) | 0.33 | (0.21 - 0.46) |
| Panama | 0.17 | (0.08 - 0.35) | 0.12 | (0.06 - 0.22) |
| Peru | 0.59 | (0.44 - 0.64) | 0.08 | (0.07 - 0.1) |
| Puerto Rico | 0.43 | (0.35 - 0.5) | 0.32 | (0.29 - 0.35) |

**Supplementary Figure 2:**
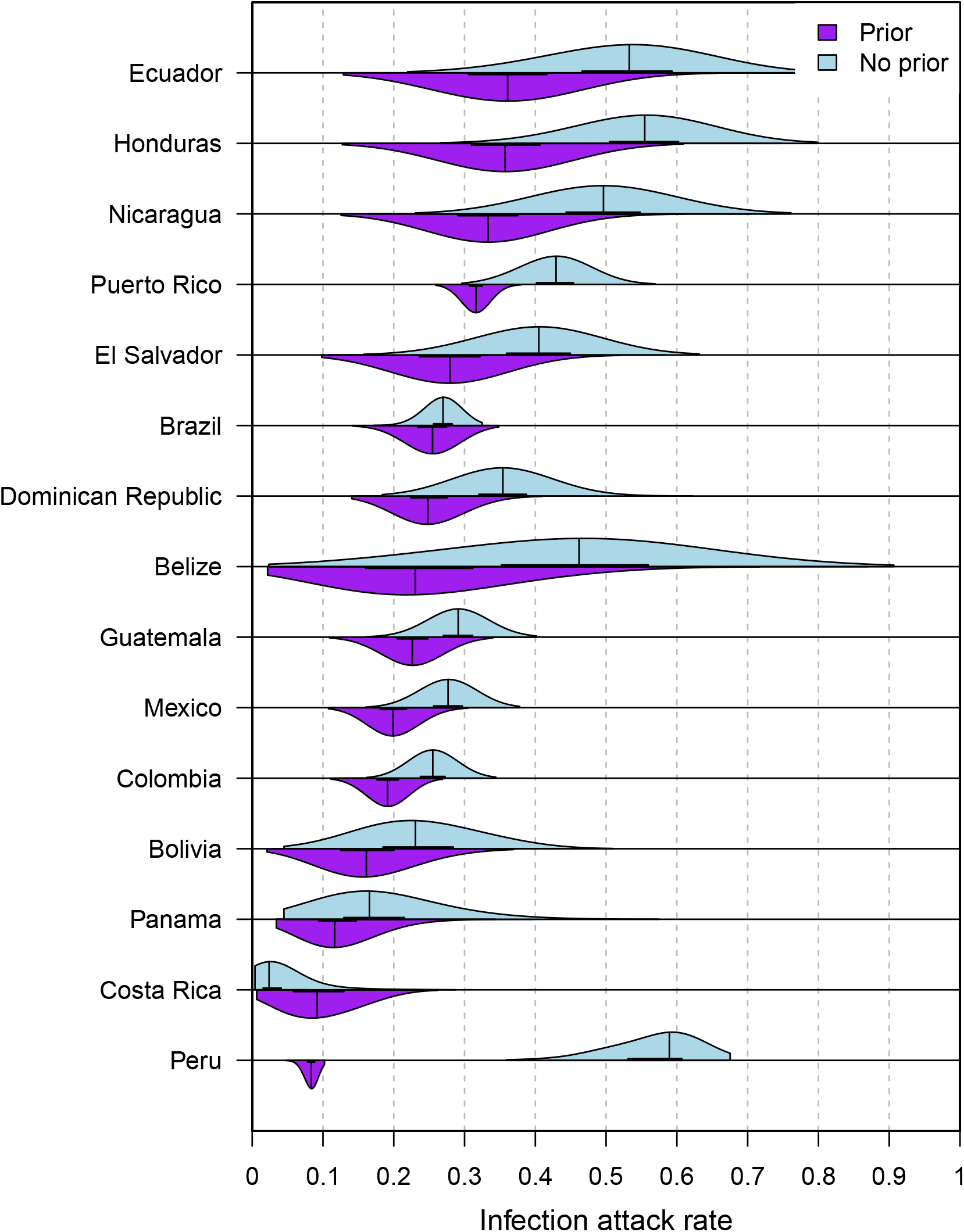
Comparison of posterior national-level IAR estimates for each country or territory with or without a Beta(1, 2) prior for subnational IARs.

**Supplementary Figure 3:**
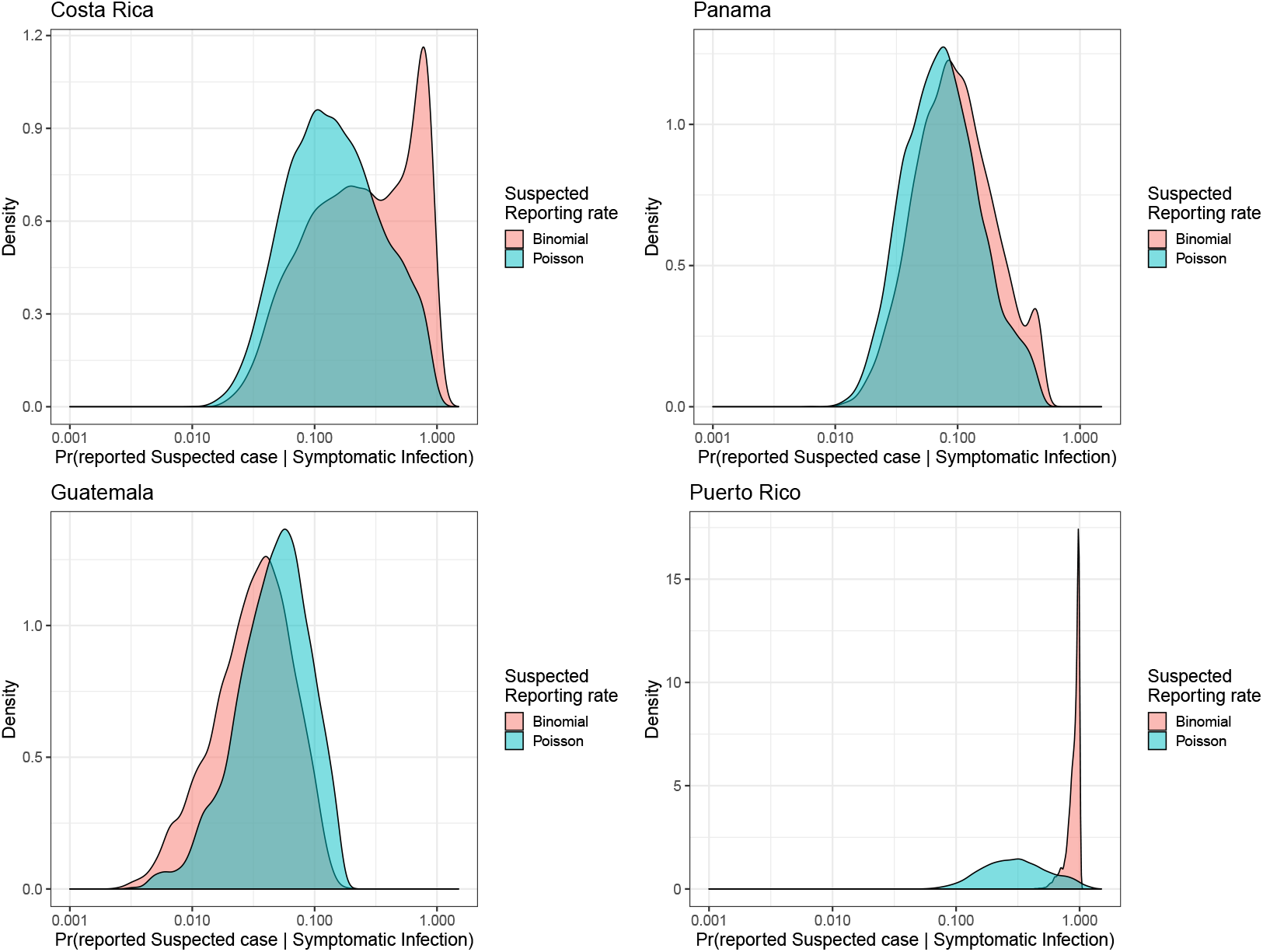
Comparison of posterior estimates from four countries of the probability that a symptomatic infection was reported as a suspected case when the reporting process was modeled using a binomial or Poisson distribution.

**Supplementary Figure 4:**
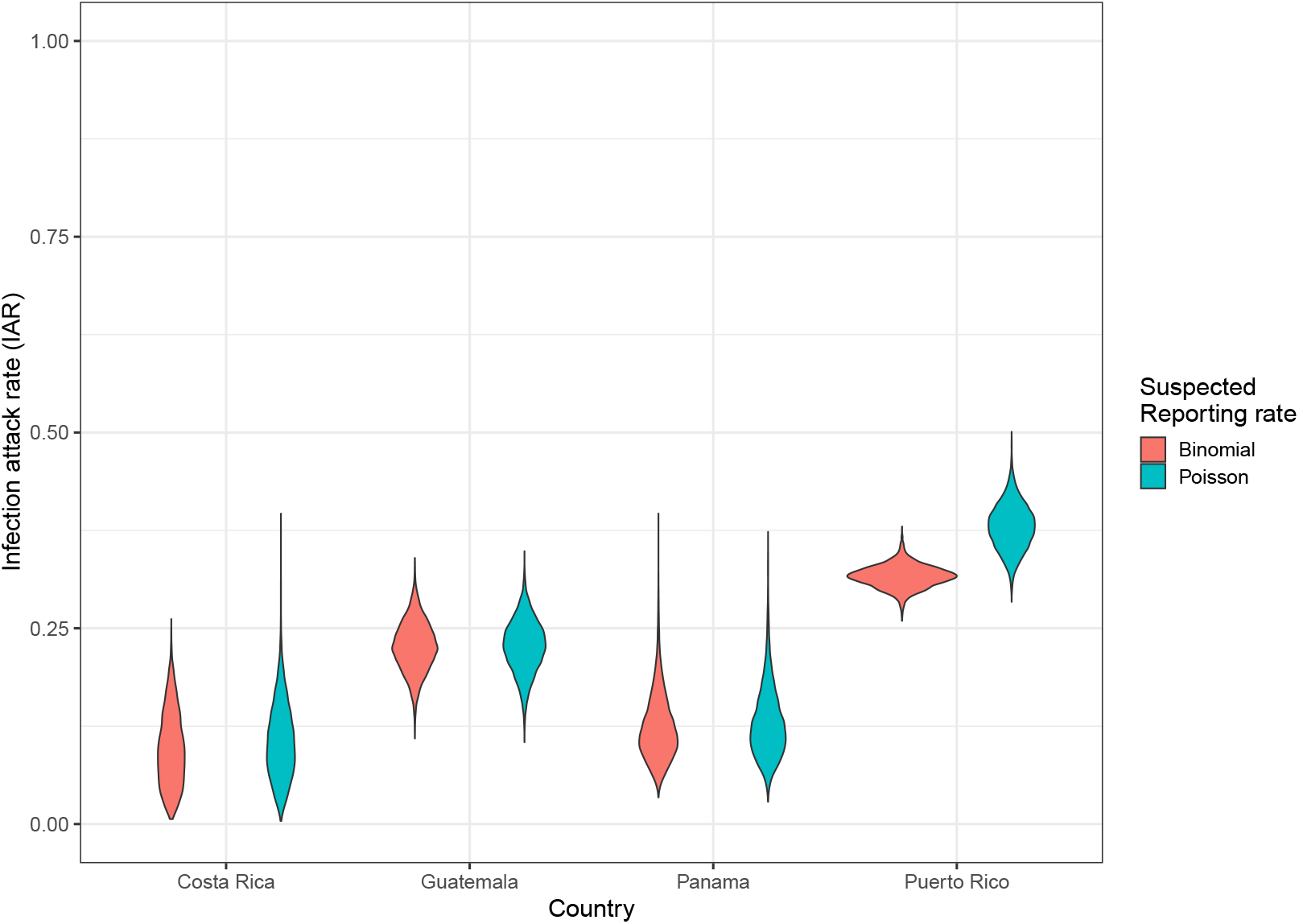
Comparison of posterior national-level IAR estimates for four countries when the reporting process for suspected cases was modeled using a binomial or Poisson distribution.

**Supplementary Figure 5:**
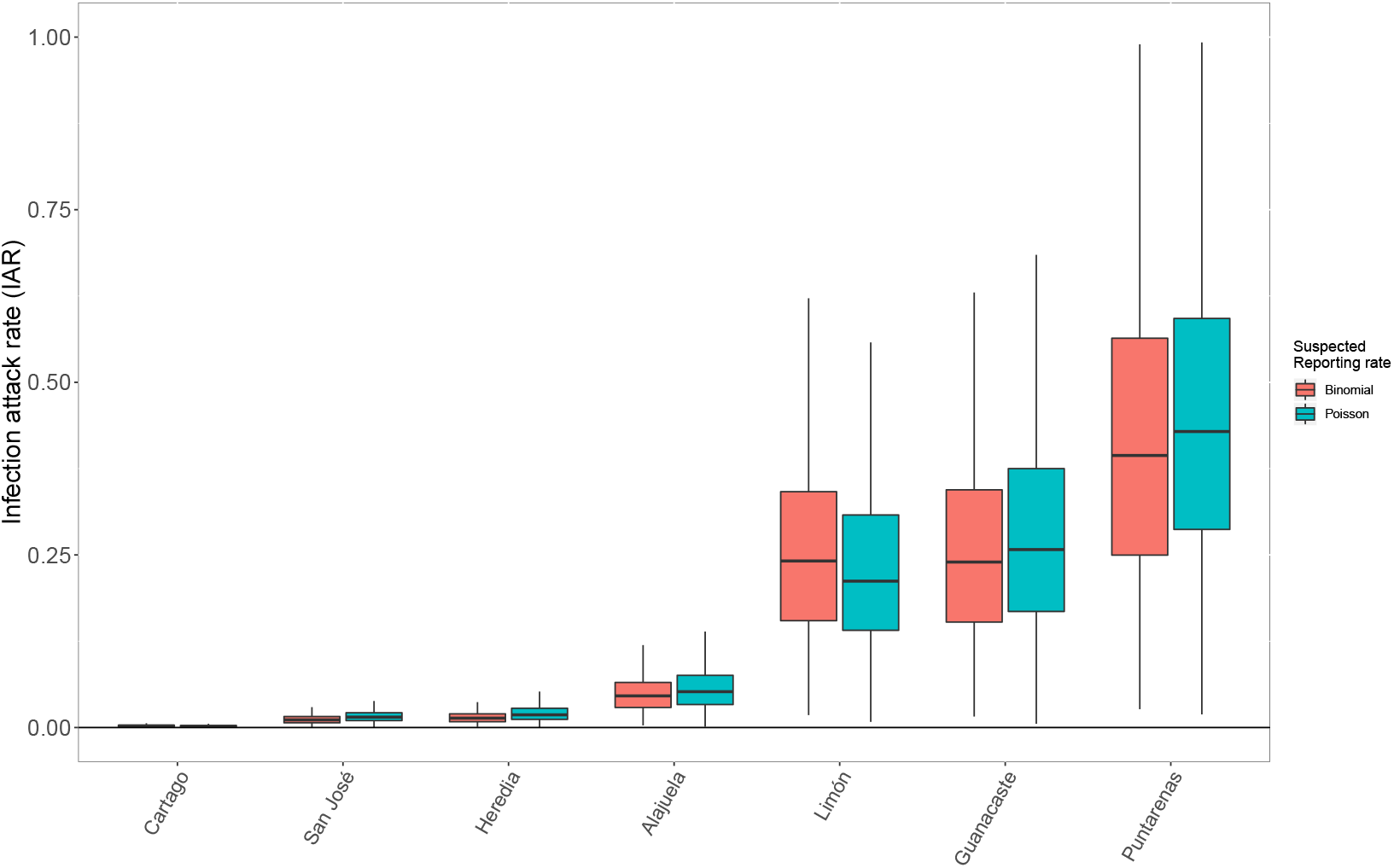
Comparison of posterior national-level IAR estimates for four countries when the reporting process for suspected cases was modeled using a binomial or Poisson distribution.

**Supplementary Figure 6:**
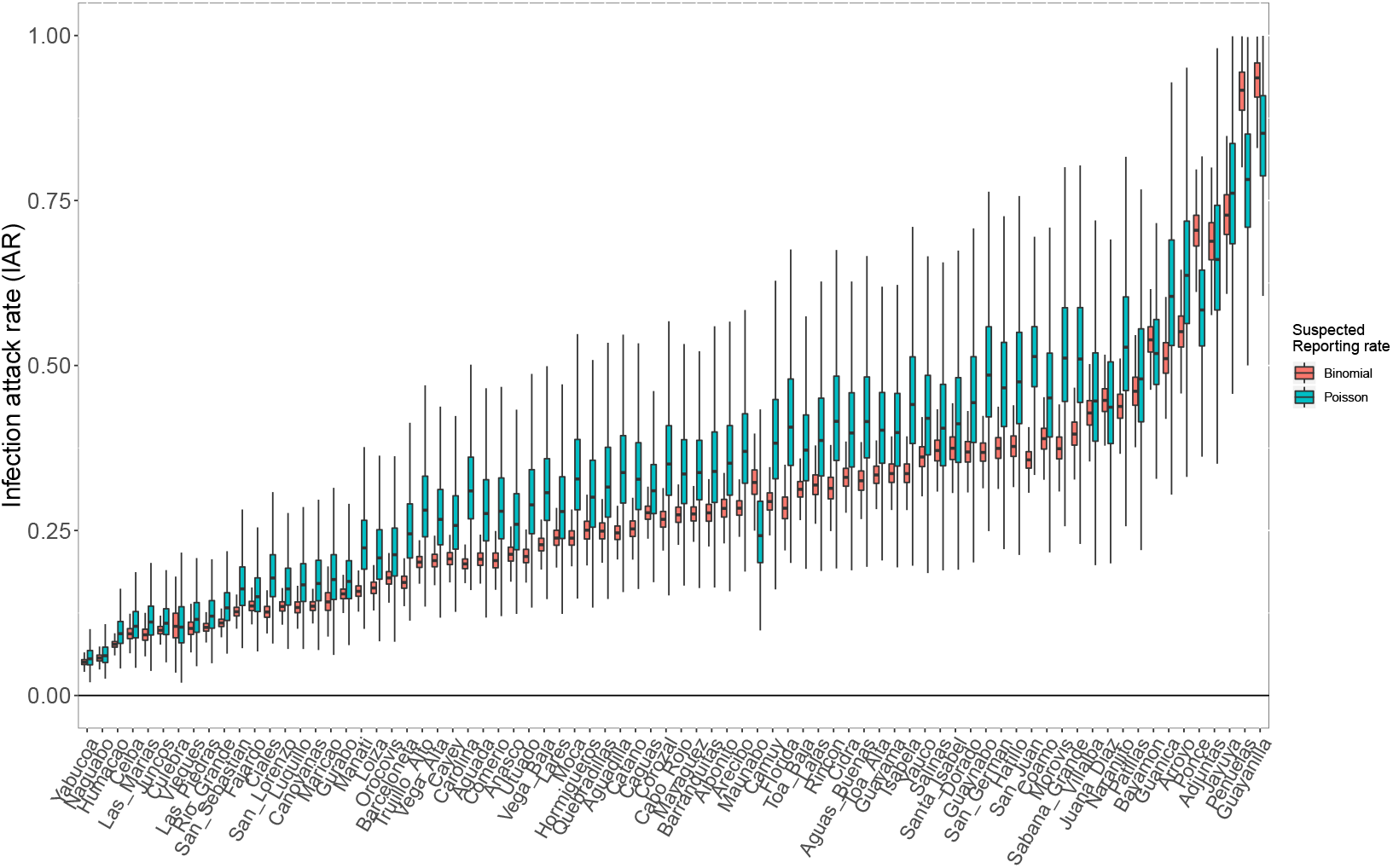
Comparison of posterior national-level IAR estimates for four countries when the reporting process for suspected cases was modeled using a binomial or Poisson distribution.

### 5 Infection attack rate projections

The territory-level IARs and number of ZIKV infections for all of the Americas were estimated by extrapolating parameter estimates from the 15 modeled countries and territories to the remaining countries and territories in the region. ZIKV IAR and the total number of ZIKV infections were estimated for the 33 countries and territories that reported confirmed Zika cases (*C*), suspected Zika cases (*S*), and Zika-associated microcephaly cases (*M*) to the Pan American Health Organization (PAHO) (2018). A national IAR estimate was obtained by drawing from the posterior distributions of the different reporting parameters from each of the 15 country models. This allowed us to draw from across the full range of estimated reporting rates from these 15 countries and territories in predicting the IARs in the remaining territories. For a given model, *k*, the probability of a given IAR value in country *j* was derived from the joint probability of the probability density functions for each of the different data types (*C*_*j*_, *S*_*j*_, and/or *M*_*j*_) that were used to fit that model. The IAR, 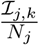, for country *j* from model *k* given *C*_*j*_ was estimated using a binomial distribution 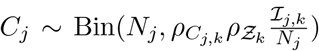, where 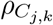 was drawn from 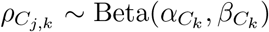 using the posterior distributions for 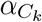 and 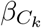 from model *k*. Similarly, 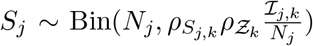 and 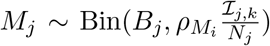. These probability densities were calculated across a range of 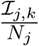 from 0 to 1 based on 1,000 draws from the posterior distribution of model *k*. The combined probability density function for 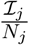 across all *K* = 15 models was 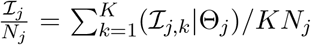.

Examples of the IAR probability distributions are presented in SI figures (22-25). The IAR probability distributions for the remaining countries and territories are located in the Github repository https://github.com/mooresea/Zika_IAR. The median IAR estimates and total number of ZIKV infections in each country and territory are provided in SI Table (4). For the 15 modeled territories, these estimates use the modeled IAR estimates rather than IAR projections based on the range of reporting rates observed across all 15 territories. A comparison of the modeled IAR estimates versus the IAR projections for each of these territories is provided in SI Table (5). The 95% CrIs for the projected IAR estimates include the mean of the model posterior for the national-level IAR for all 15 modeled territories. However, for most territories the 95% CrI for the projections is very broad, with a lower bound near 0 and an upper bound at 1. These broad credible intervals are an inevitable result of the high variability in estimated reporting rates among the modeled territories, which results in a high uncertainty in IARs when sampling reporting rates from across the posterior distributions from every model. The median projected IAR is very similar to the modeled IAR estimate for several countries (e.g., Brazil, Honduras, Nicaragua), but drastically different for several other territories, including Belize and Puerto Rico. These results suggest that the projections for non-modeled territories should be interpreted cautiously. Fortunately, most of the non-modeled countries have small populations and do not contribute significantly to the continent-wide estimate of ZIKV infections.

**Supplementary Table 4:** Infection attacks rates and total infections.

| Country | IAR | 95% CrI | Infections | 95% CrI |
| --- | --- | --- | --- | --- |
| Brazil | 0.25 | (0.19 - 0.31) | 53,388,206 | (40,819,653 - 64,922,428) |
| Mexico | 0.20 | (0.15 - 0.25) | 25,599,729 | (19,237,945 - 32,690,560) |
| Colombia | 0.19 | (0.15 - 0.23) | 9,302,116 | ( 7,133,364 - 11,360,866) |
| Venezuela | 0.21 | (0.01 - 1) | 6,633,999 | ( 350,008 - 31,518,000) |
| Ecuador | 0.36 | (0.21 - 0.51) | 5,956,506 | ( 3,523,747 - 8,484,736) |
| Guatemala | 0.23 | (0.16 - 0.29) | 3,772,441 | ( 2,714,844 - 4,819,782) |
| Honduras | 0.36 | (0.22 - 0.49) | 2,926,168 | ( 1,824,954 - 4,044,226) |
| Peru | 0.08 | (0.07 - 0.1) | 2,672,621 | ( 2,129,464 - 3,054,319) |
| Dominican Republic | 0.25 | (0.18 - 0.33) | 2,657,916 | ( 1,923,663 - 3,538,936) |
| Nicaragua | 0.33 | (0.21 - 0.46) | 2,061,039 | ( 1,320,432 - 2,841,540) |
| Bolivia | 0.16 | (0.07 - 0.29) | 1,769,432 | ( 793,925 - 3,172,282) |
| El Salvador | 0.28 | (0.16 - 0.4) | 1,717,254 | ( 974,790 - 2,482,731) |
| Puerto Rico | 0.32 | (0.29 - 0.35) | 1,164,139 | ( 1,059,133 - 1,270,456) |
| Trinidad and Tobago | 0.62 | (0.01 - 1) | 847,316 | ( 9,648 - 1,366,851) |
| Jamaica | 0.28 | (0.02 - 1) | 787,408 | ( 47,988 - 2,808,000) |
| Suriname | 0.90 | (0.04 - 1) | 495,564 | ( 21,279 - 548,000) |
| Guadeloupe | 1.00 | (0.34 - 1) | 472,000 | ( 158,442 - 472,000) |
| Panama | 0.12 | (0.06 - 0.22) | 465,004 | ( 238,134 - 886,204) |
| Costa Rica | 0.09 | (0.02 - 0.19) | 448,074 | ( 94,060 - 942,369) |
| Martinique | 1.00 | (0.09 - 1) | 396,000 | ( 34,131 - 396,000) |
| French Guiana | 1.00 | (0.23 - 1) | 275,970 | ( 64,676 - 276,000) |
| Haiti | 0.02 | (0 - 1) | 272,412 | ( 6,580 - 10,910,543) |
| Cuba | 0.02 | (0 - 0.98) | 192,194 | ( 2,322 - 11,193,532) |
| Curacao | 1.00 | (0.28 - 1) | 148,974 | ( 41,318 - 149,000) |
| Barbados | 0.43 | (0.02 - 1) | 124,633 | ( 5,303 - 291,947) |
| Argentina | 0.00 | (0 - 0.73) | 122,880 | ( 4,448 - 32,221,973) |
| Aruba | 1.00 | (0.11 - 1) | 113,850 | ( 11,981 - 114,000) |
| Virgin Islands (US) | 1.00 | (0.13 - 1) | 102,910 | ( 12,980 - 103,000) |
| Saint Lucia | 0.62 | (0.04 - 1) | 102,142 | ( 5,781 - 164,984) |
| Belize | 0.23 | (0.07 - 0.49) | 85,497 | ( 24,747 - 181,433) |
| Saint Vincent and the Grenadines | 0.82 | (0.04 - 1) | 83,376 | ( 3,914 - 101,986) |
| Paraguay | 0.01 | (0 - 0.99) | 74,808 | ( 4,172 - 6,638,558) |
| Dominica | 1.00 | (0.11 - 1) | 73,802 | ( 7,838 - 74,000) |
| Grenada | 0.58 | (0.03 - 1) | 64,499 | ( 2,770 - 110,971) |
| Bahamas | 0.16 | (0.01 - 1) | 61,828 | ( 3,222 - 394,768) |
| Antigua and Barbuda | 0.65 | (0.03 - 1) | 60,924 | ( 3,163 - 93,985) |
| Saint Kitts and Nevis | 0.98 | (0.06 - 1) | 51,950 | ( 3,397 - 52,996) |
| Guyana | 0.06 | (0 - 0.98) | 48,304 | ( 545 - 757,640) |
| Sint Maarten (Dutch part) | 0.99 | (0.06 - 1) | 41,465 | ( 2,441 - 41,994) |
| Cayman Islands | 0.63 | (0.03 - 1) | 36,408 | ( 1,554 - 57,974) |
| Saint Martin | 1.00 | (0.57 - 1) | 35,989 | ( 20,419 - 36,000) |
| Turks and Caicos Islands | 0.57 | (0.03 - 1) | 29,828 | ( 1,459 - 51,969) |
| Bonaire, St Eustatius and Saba | 1.00 | (0.16 - 1) | 24,939 | ( 4,076 - 25,000) |
| Virgin Islands (UK) | 0.53 | (0.02 - 1) | 18,682 | ( 819 - 34,967) |
| Saint Barthelemy | 1.00 | (0.65 - 1) | 9,990 | ( 6,503 - 10,000) |
| Anguilla | 0.47 | (0.02 - 1) | 7,957 | ( 345 - 16,953) |
| Montserrat | 0.62 | (0.03 - 0.99) | 3,086 | ( 145 - 4,975) |
| Uruguay | 0.00 | (0 - 0.01) | 0 | ( 0 - 38,519) |

**Supplementary Table 5:**
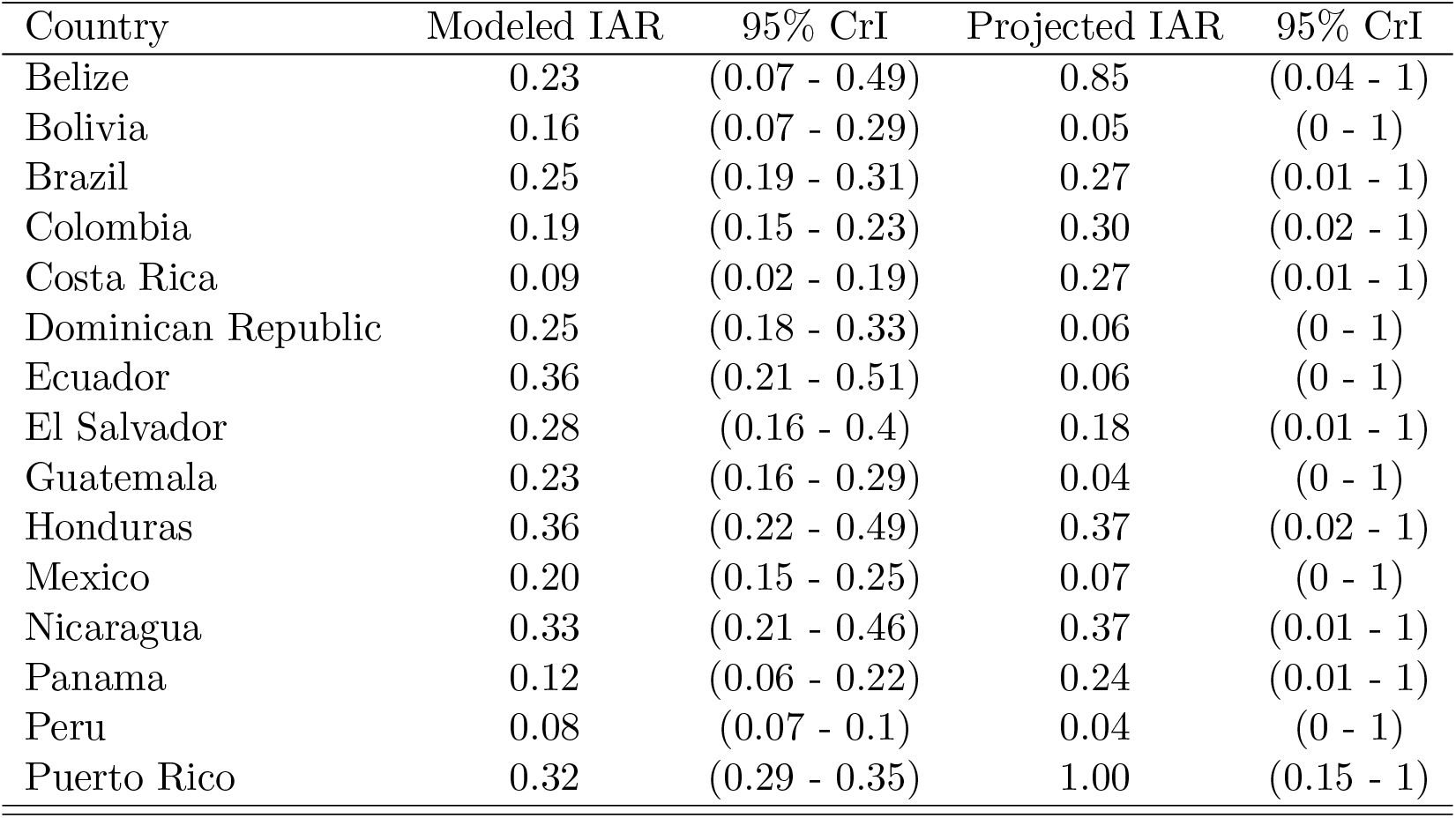
Comparison of model-specific IAR estimates for each modeled territory versus projections of IAR from combined model estimates.

**Supplementary Table 6:**
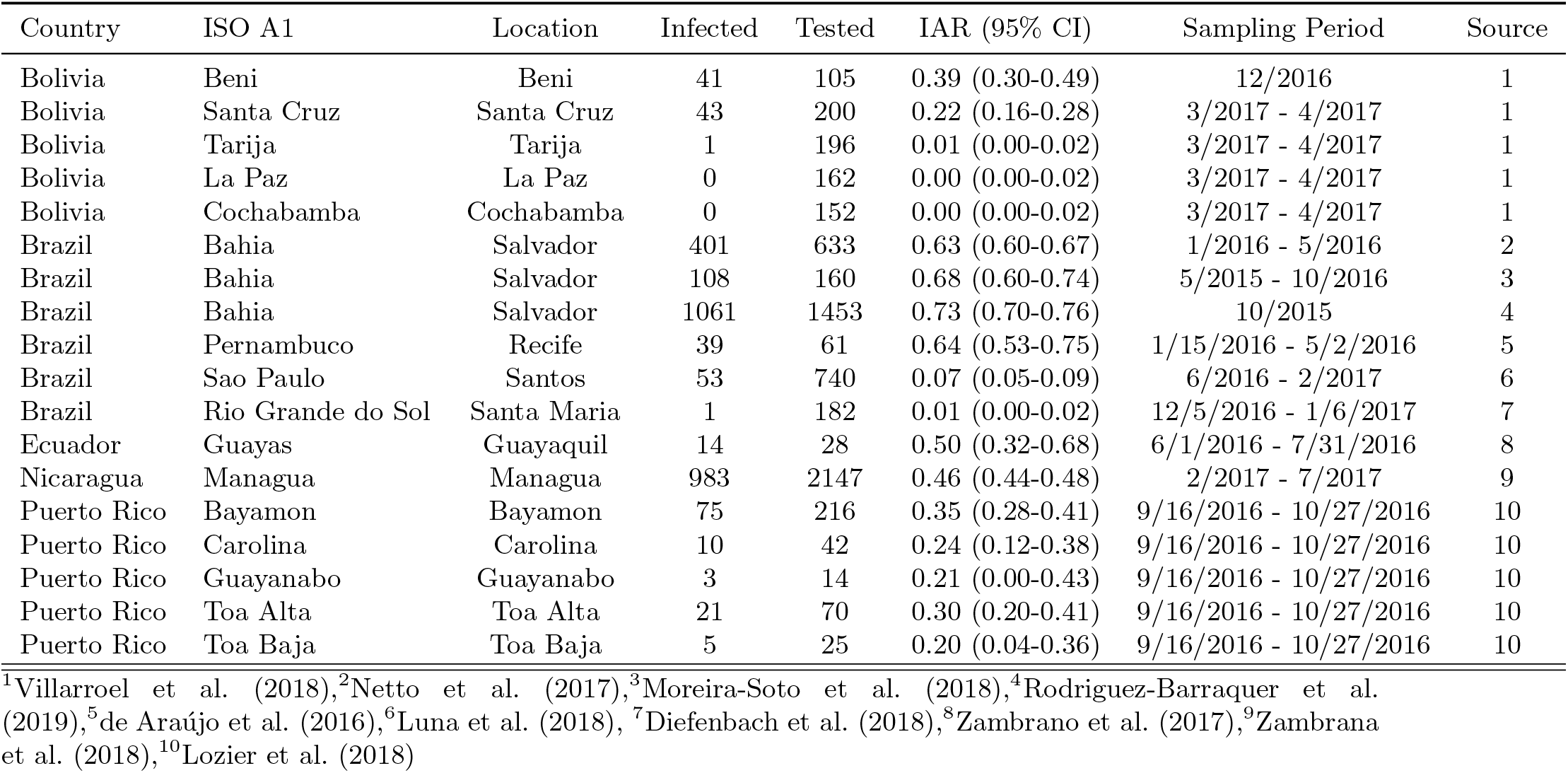
Zika seroprevalence studies. See footnotes for references.

| Country | ISO A1 | Location | Infected | Tested | IAR (95% CI) | Sampling Period | Source |
| --- | --- | --- | --- | --- | --- | --- | --- |
| Bolivia | Beni | Beni | 41 | 105 | 0.39 (0.30-0.49) | 12/2016 | 1 |
| Bolivia | Santa Cruz | Santa Cruz | 43 | 200 | 0.22 (0.16-0.28) | 3/2017 - 4/2017 | 1 |
| Bolivia | Tarija | Tarija | 1 | 196 | 0.01 (0.00-0.02) | 3/2017 - 4/2017 | 1 |
| Bolivia | La Paz | La Paz | 0 | 162 | 0.00 (0.00-0.02) | 3/2017 - 4/2017 | 1 |
| Bolivia | Cochabamba | Cochabamba | 0 | 152 | 0.00 (0.00-0.02) | 3/2017 - 4/2017 | 1 |
| Brazil | Bahia | Salvador | 401 | 633 | 0.63 (0.60-0.67) | 1/2016 - 5/2016 | 2 |
| Brazil | Bahia | Salvador | 108 | 160 | 0.68 (0.60-0.74) | 5/2015 - 10/2016 | 3 |
| Brazil | Bahia | Salvador | 1061 | 1453 | 0.73 (0.70-0.76) | 10/2015 | 4 |
| Brazil | Pernambuco | Recife | 39 | 61 | 0.64 (0.53-0.75) | 1/15/2016 - 5/2/2016 | 5 |
| Brazil | Sao Paulo | Santos | 53 | 740 | 0.07 (0.05-0.09) | 6/2016 - 2/2017 | 6 |
| Brazil | Rio Grande do Sol | Santa Maria | 1 | 182 | 0.01 (0.00-0.02) | 12/5/2016 - 1/6/2017 | 7 |
| Ecuador | Guayas | Guayaquil | 14 | 28 | 0.50 (0.32-0.68) | 6/1/2016 - 7/31/2016 | 8 |
| Nicaragua | Managua | Managua | 983 | 2147 | 0.46 (0.44-0.48) | 2/2017 - 7/2017 | 9 |
| Puerto Rico | Bayamon | Bayamon | 75 | 216 | 0.35 (0.28-0.41) | 9/16/2016 - 10/27/2016 | 10 |
| Puerto Rico | Carolina | Carolina | 10 | 42 | 0.24 (0.12-0.38) | 9/16/2016 - 10/27/2016 | 10 |
| Puerto Rico | Guayanabo | Guayanabo | 3 | 14 | 0.21 (0.00-0.43) | 9/16/2016 - 10/27/2016 | 10 |
| Puerto Rico | Toa Alta | Toa Alta | 21 | 70 | 0.30 (0.20-0.41) | 9/16/2016 - 10/27/2016 | 10 |
| Puerto Rico | Toa Baja | Toa Baja | 5 | 25 | 0.20 (0.04-0.36) | 9/16/2016 - 10/27/2016 | 10 |

<sup>1</sup>Villarroel et al. (2018),<sup>2</sup>Netto et al. (2017),<sup>3</sup>Moreira-Soto et al. (2018),<sup>4</sup>Rodriguez-Barraquer et al. (2019),<sup>5</sup>de Araújo et al. (2016),<sup>6</sup>Luna et al. (2018),<sup>7</sup>Diefenbach et al. (2018),<sup>8</sup>Zambrano et al. (2017),<sup>9</sup>Zambrana et al. (2018),<sup>10</sup>Lozier et al. (2018)

**Supplementary Figure 7:**
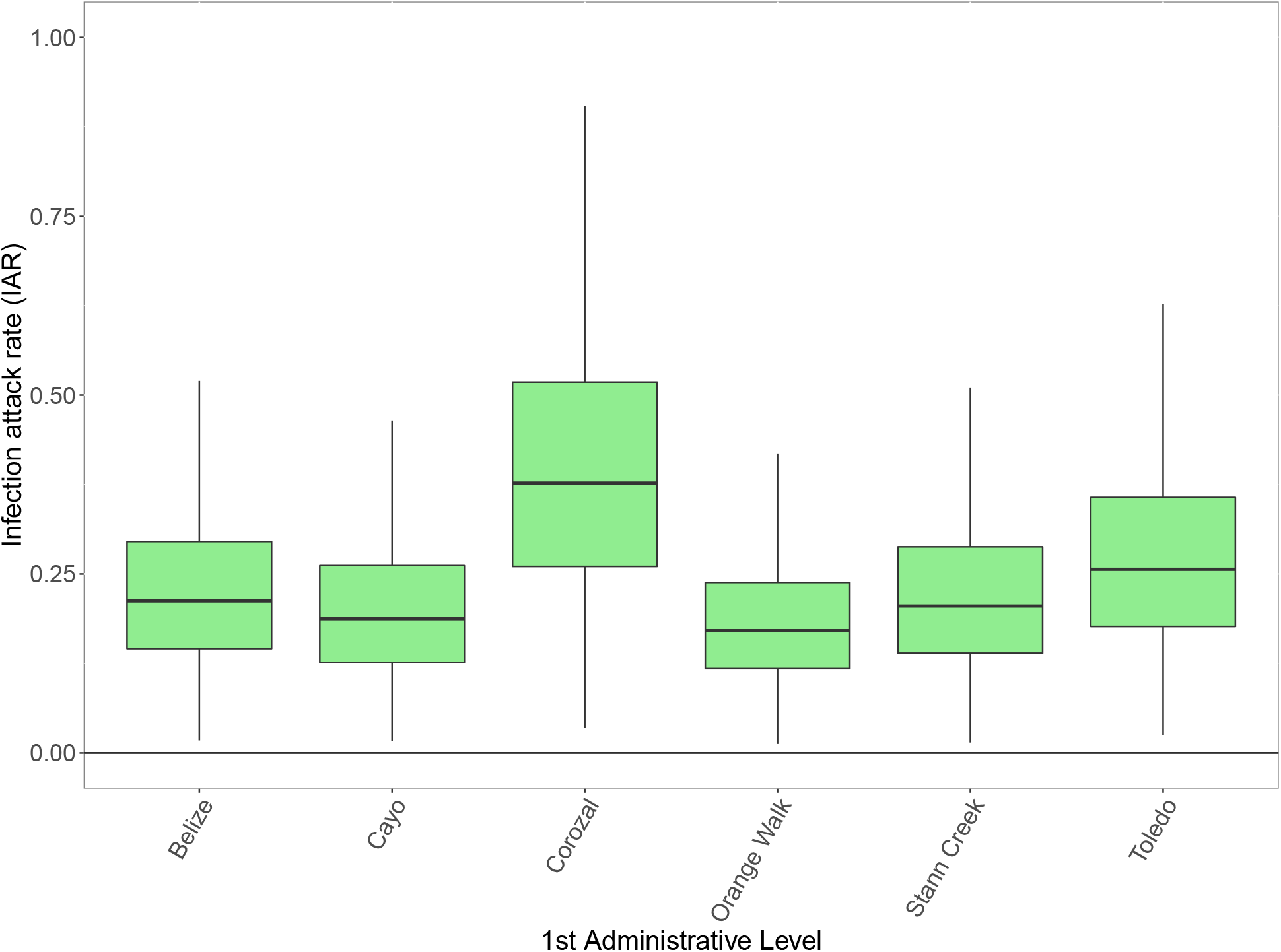
Posterior distribution of subnational infection attack rates (IAR) for Belize.

**Supplementary Figure 8:**
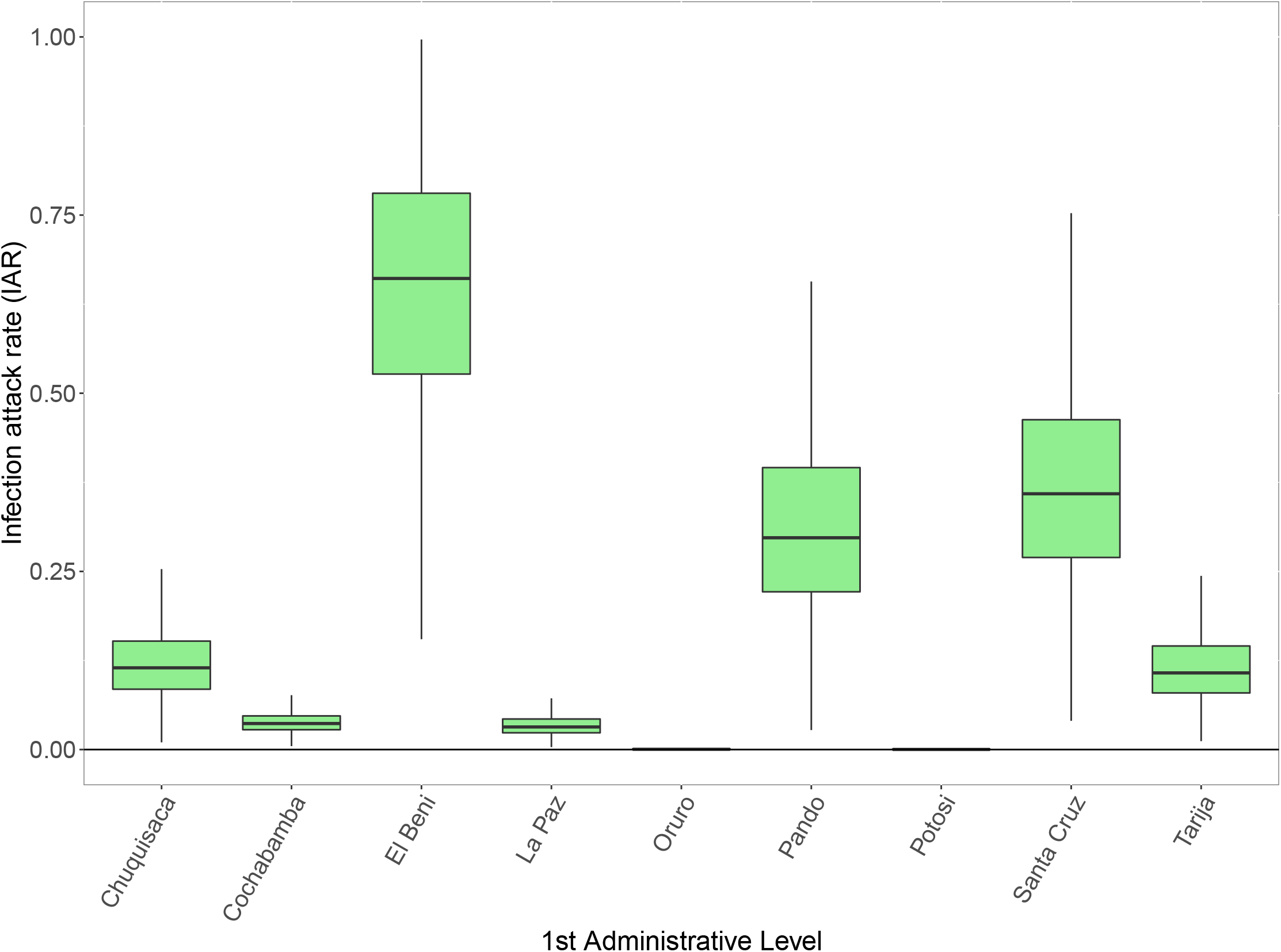
Posterior distribution of subnational infection attack rates (IAR) for Bolivia.

**Supplementary Figure 9:**
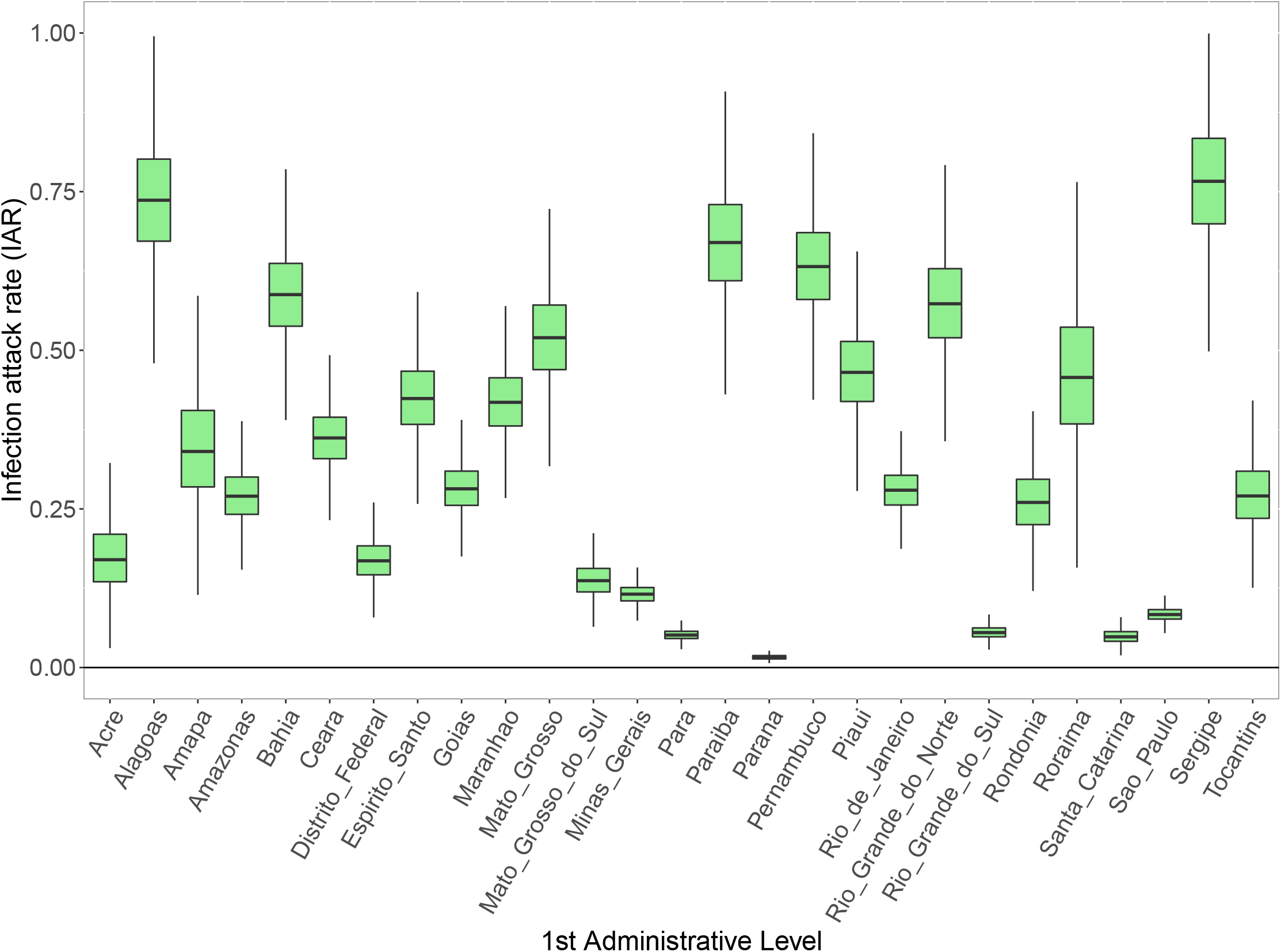
Posterior distribution of subnational infection attack rates (IAR) for Brazil.

**Supplementary Figure 10:**
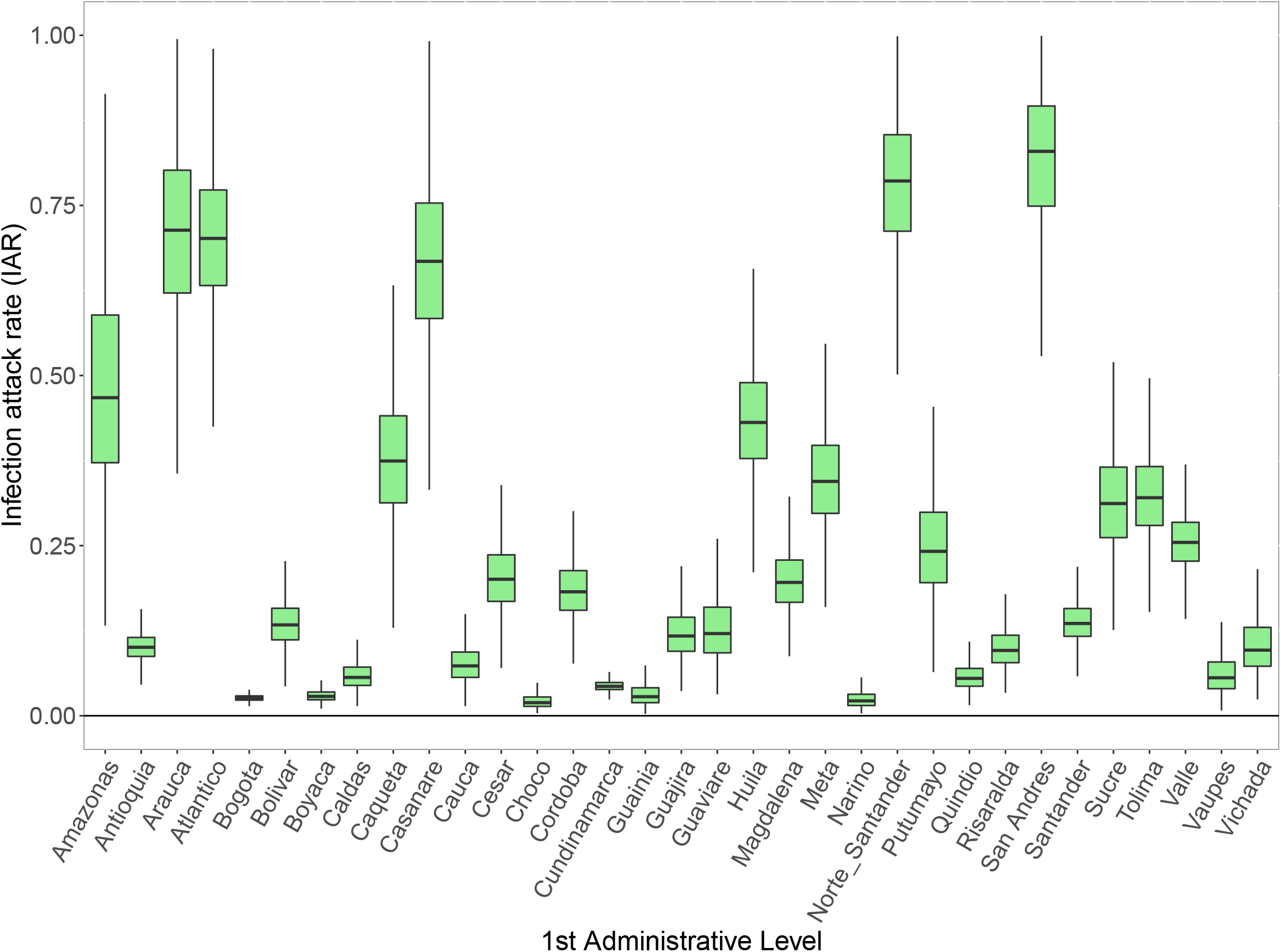
Posterior distribution of subnational infection attack rates (IAR) for Colombia.

**Supplementary Figure 11:**
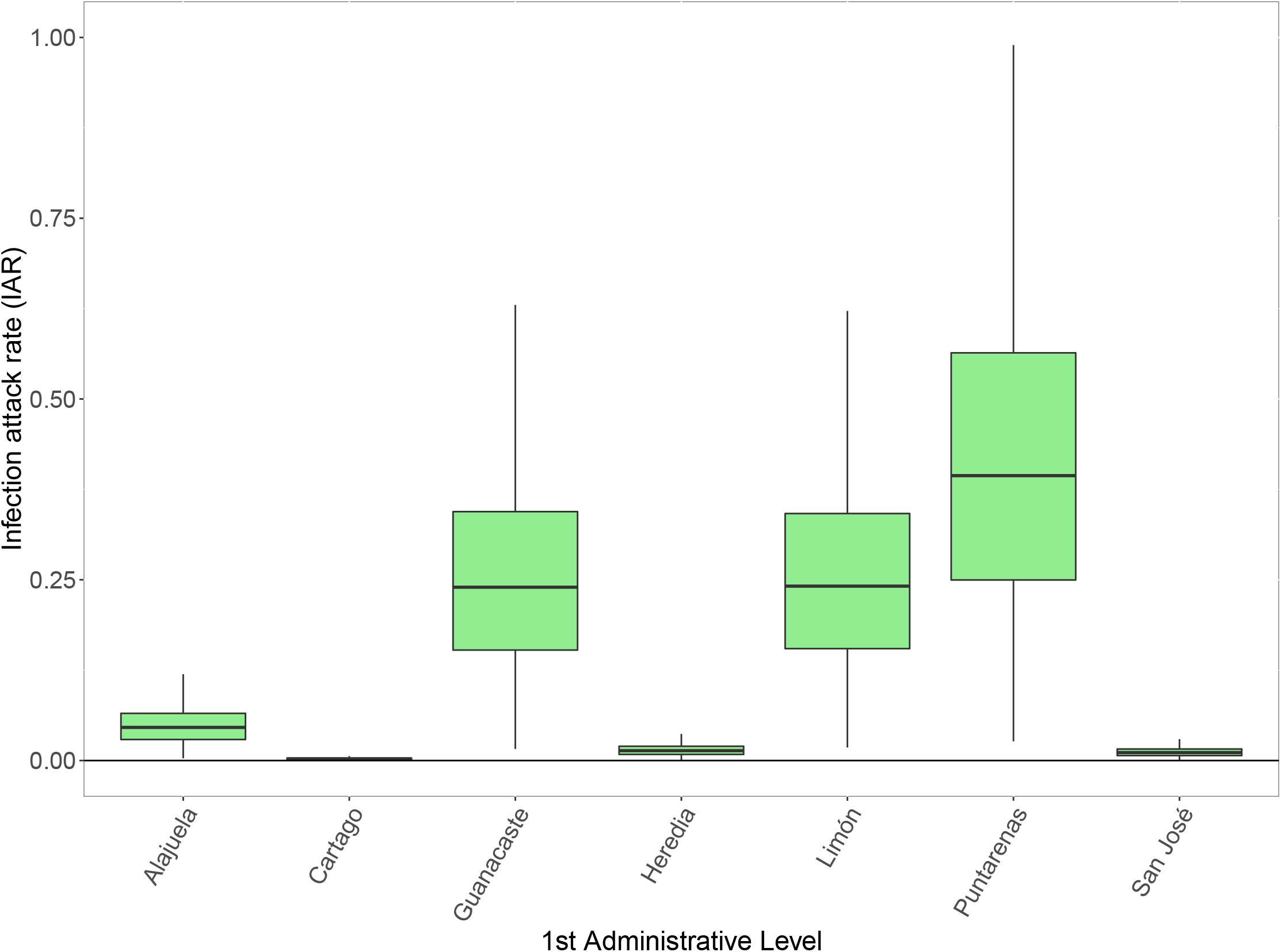
Posterior distribution of subnational infection attack rates (IAR) for Costa Rica.

**Supplementary Figure 12:**
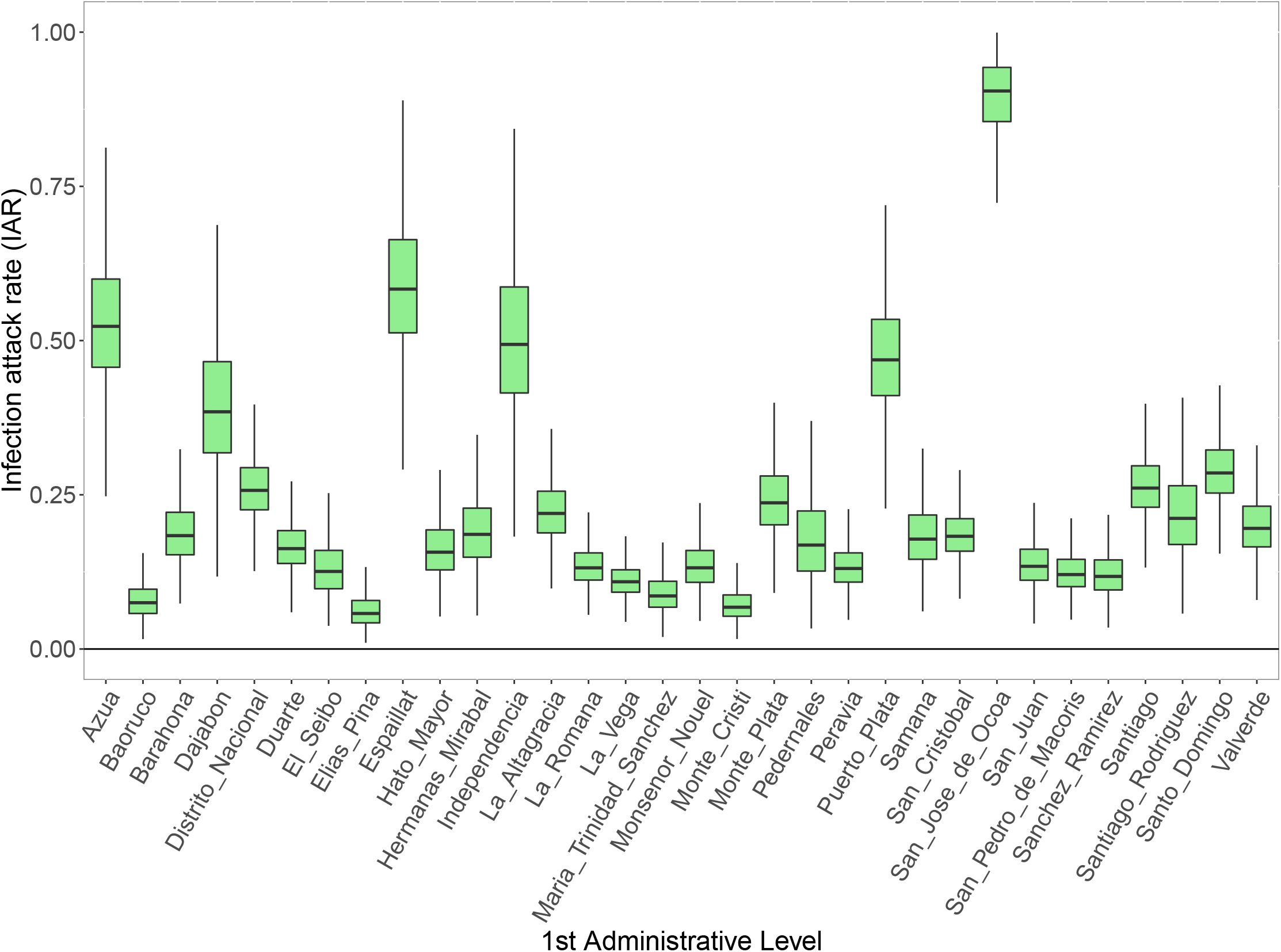
Posterior distribution of subnational infection attack rates (IAR) for the Dominican republic.

**Supplementary Figure 13:**
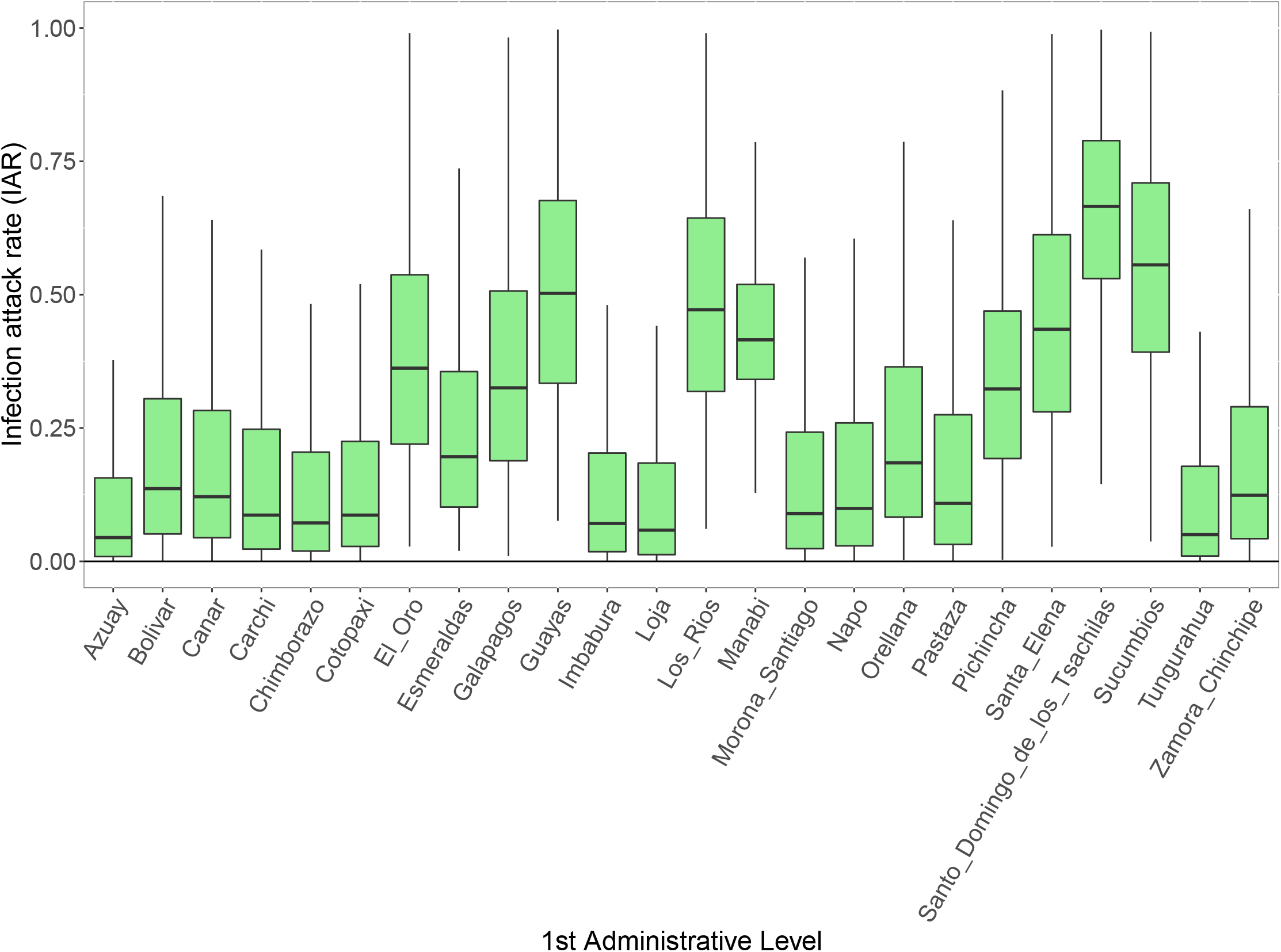
Posterior distribution of subnational infection attack rates (IAR) for Ecuador.

**Supplementary Figure 14:**
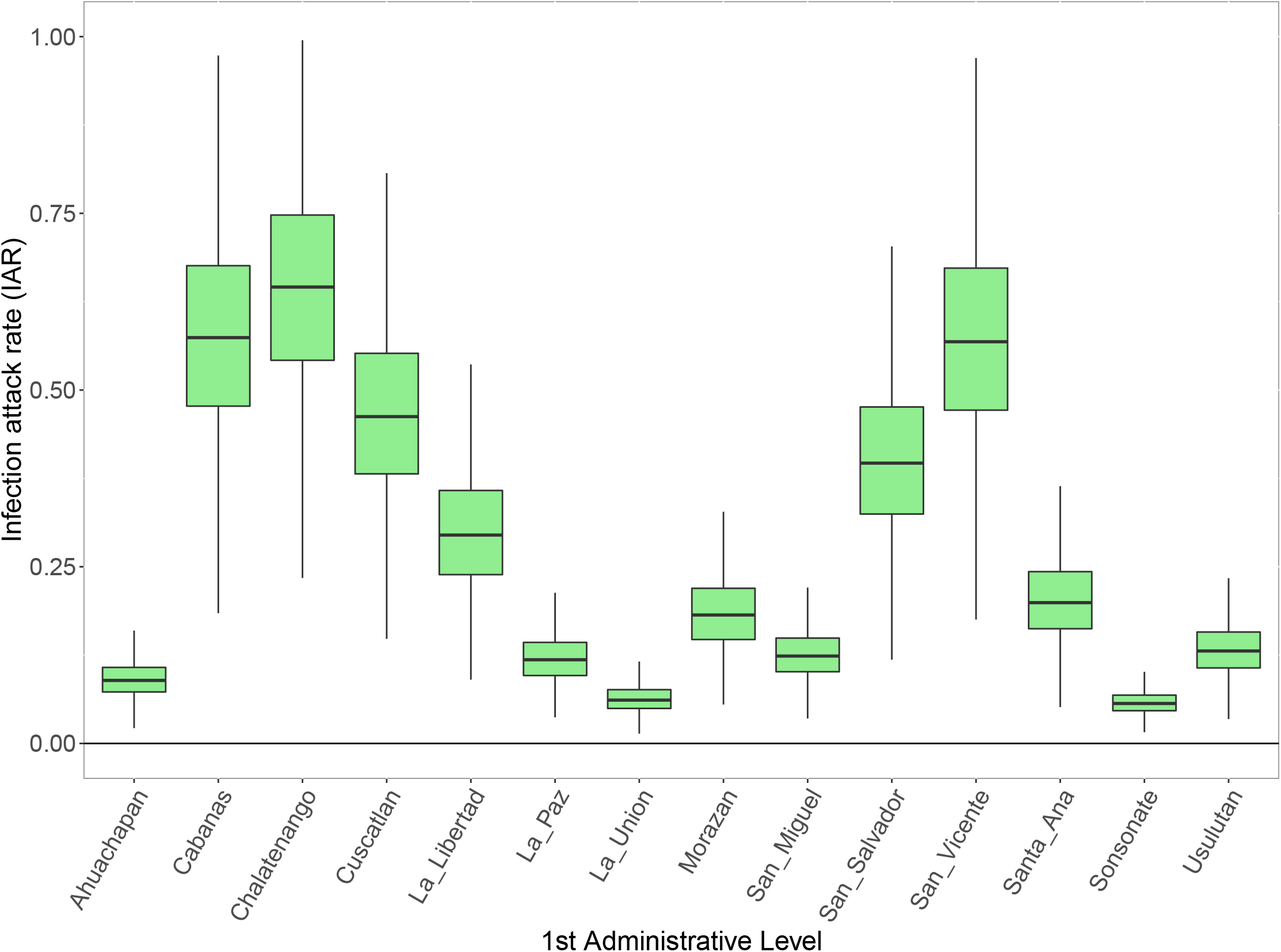
Posterior distribution of subnational infection attack rates (IAR) for El Salvador.

**Supplementary Figure 15:**
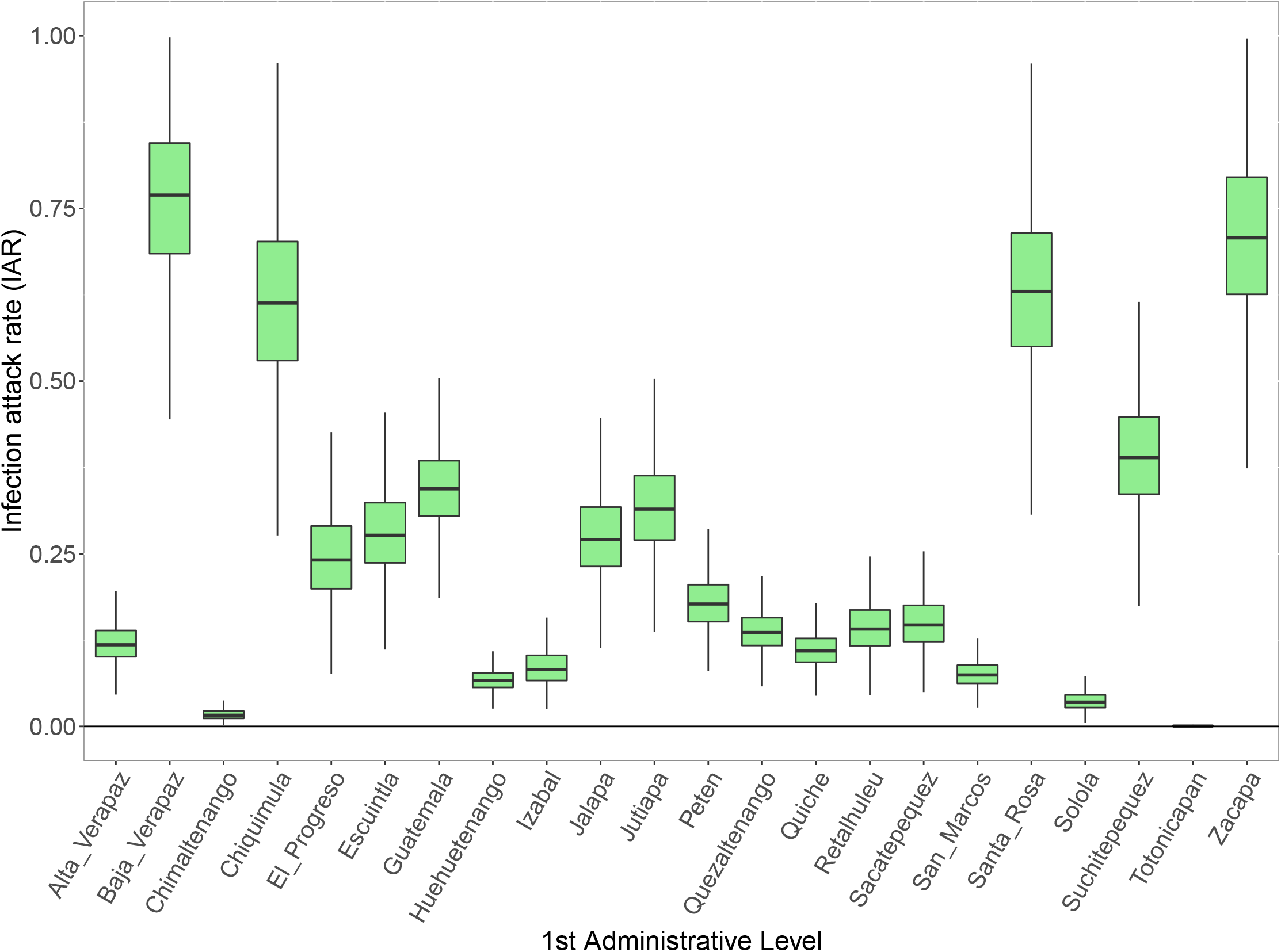
Posterior distribution of subnational infection attack rates (IAR) for Guatemala.

**Supplementary Figure 16:**
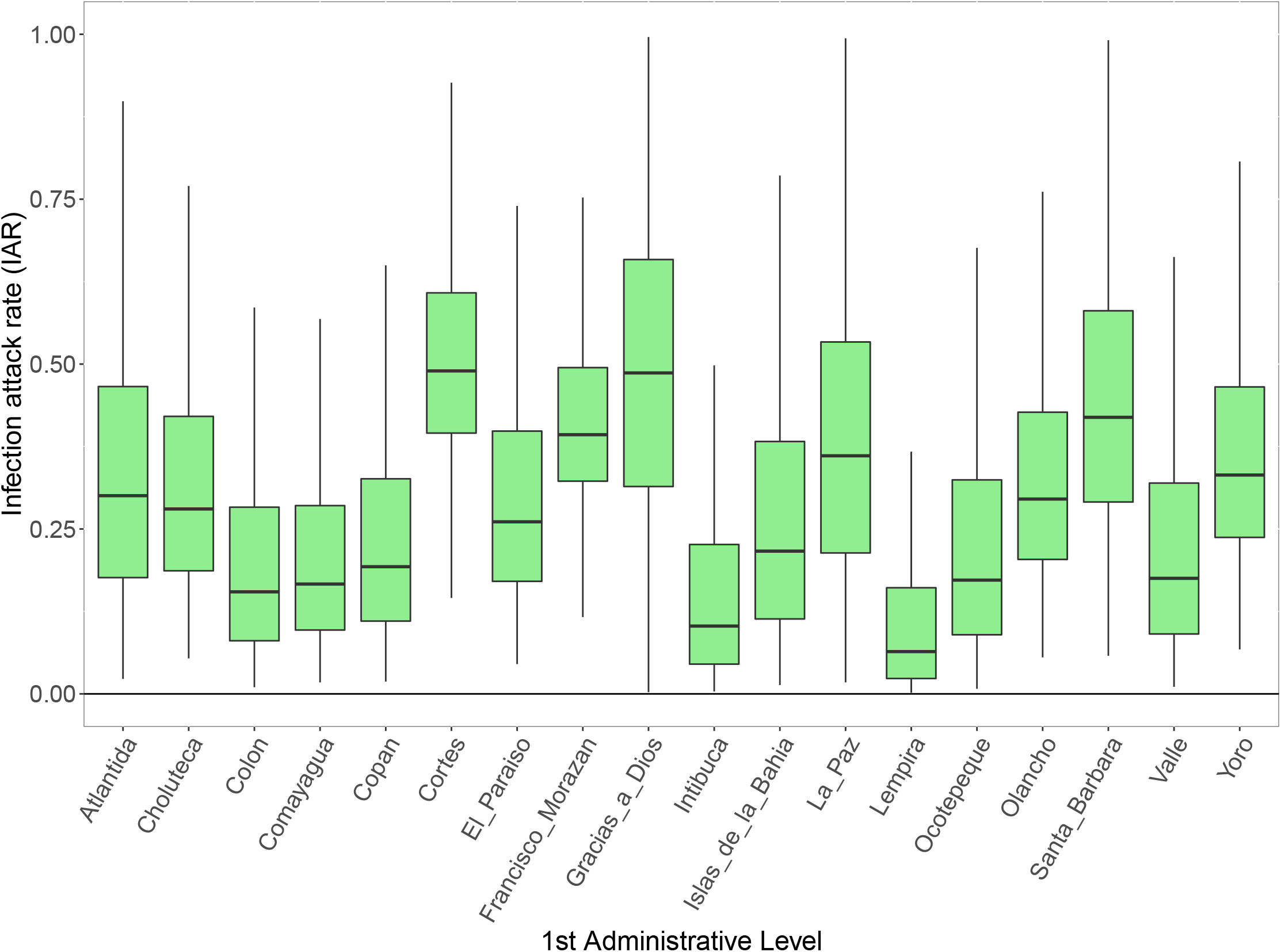
Posterior distribution of subnational infection attack rates (IAR) for Honduras.

**Supplementary Figure 17:**
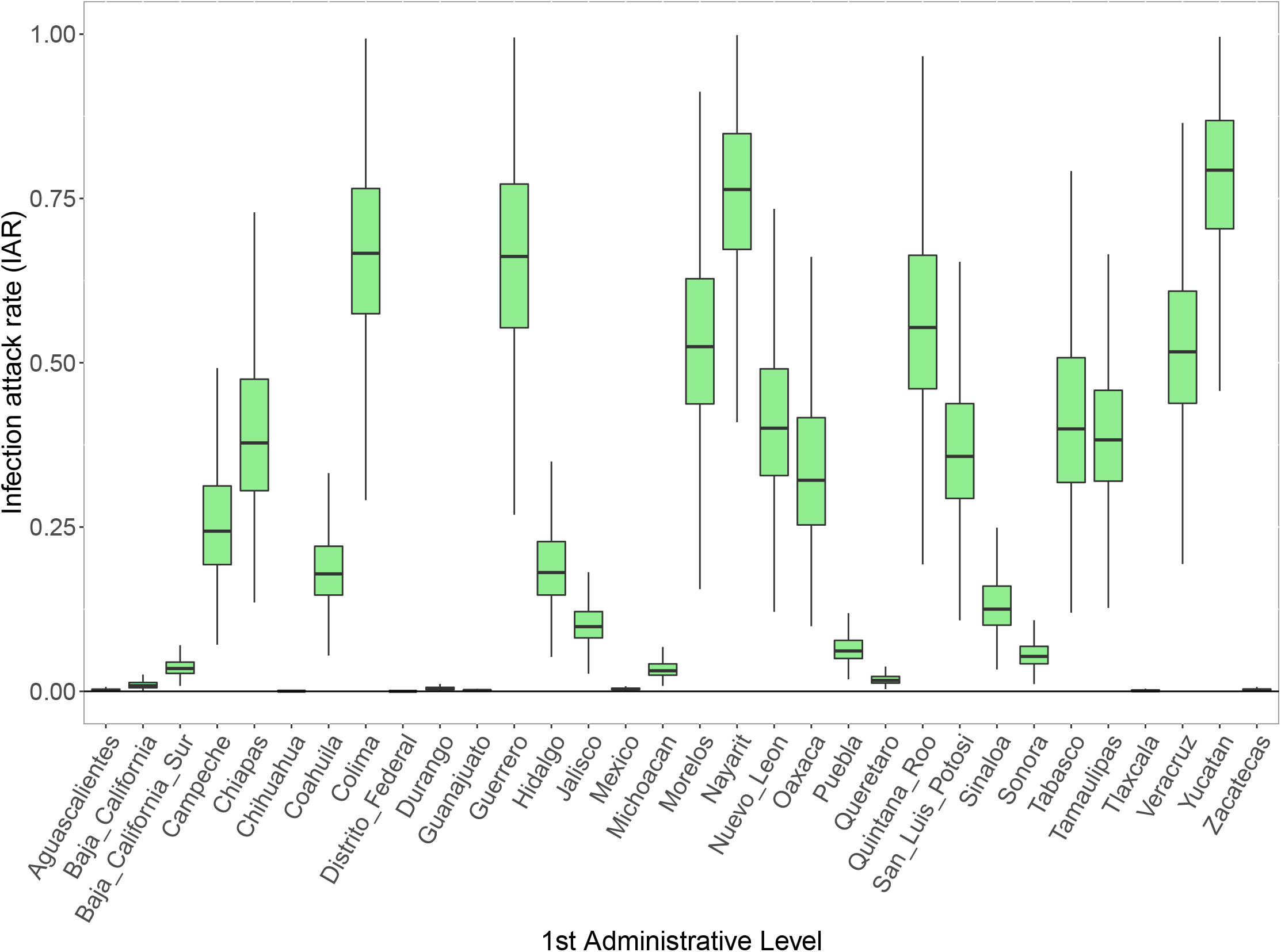
Posterior distribution of subnational infection attack rates (IAR) for Mexico.

**Supplementary Figure 18:**
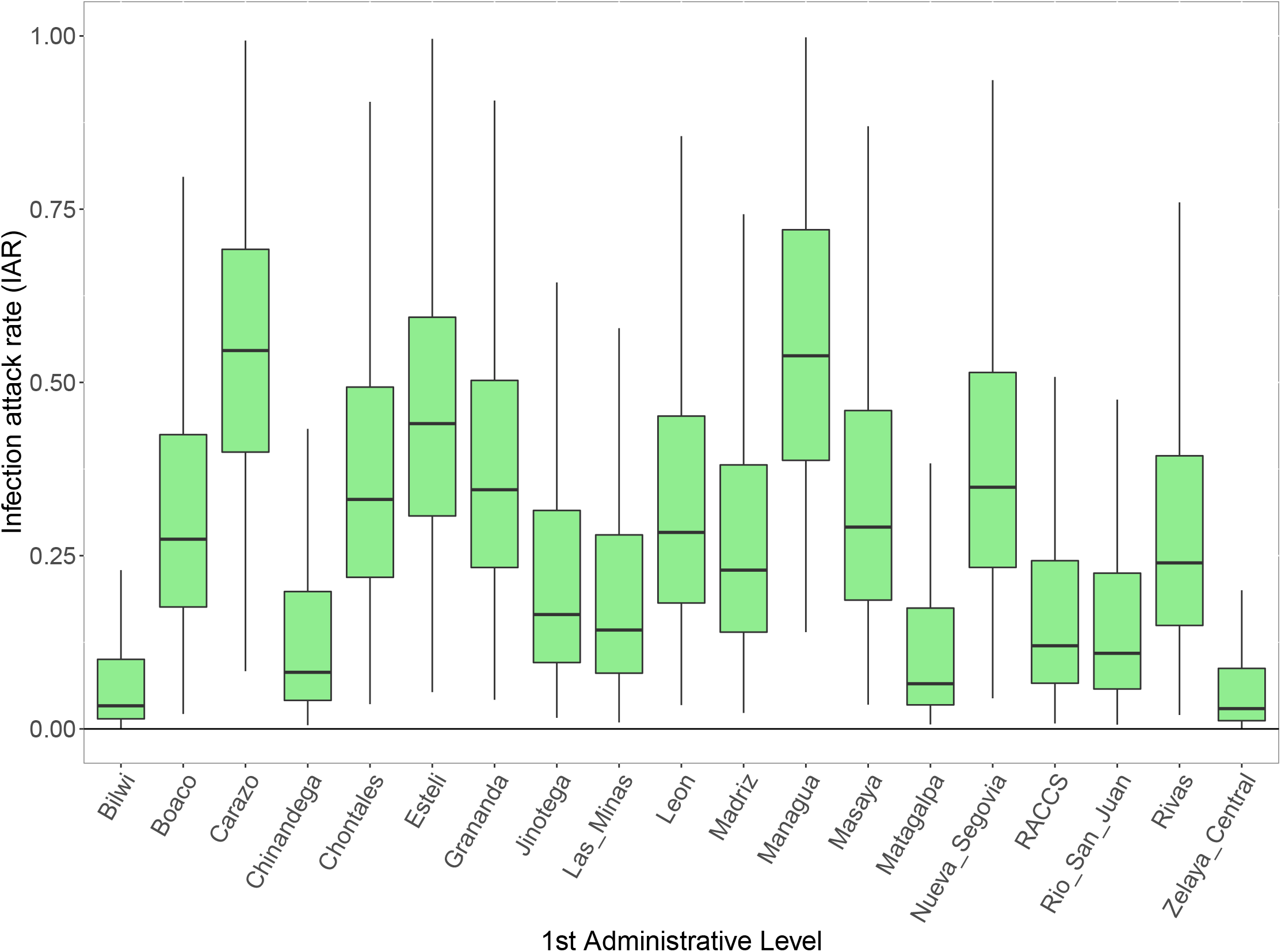
Posterior distribution of subnational infection attack rates (IAR) for Nicaragua.

**Supplementary Figure 19:**
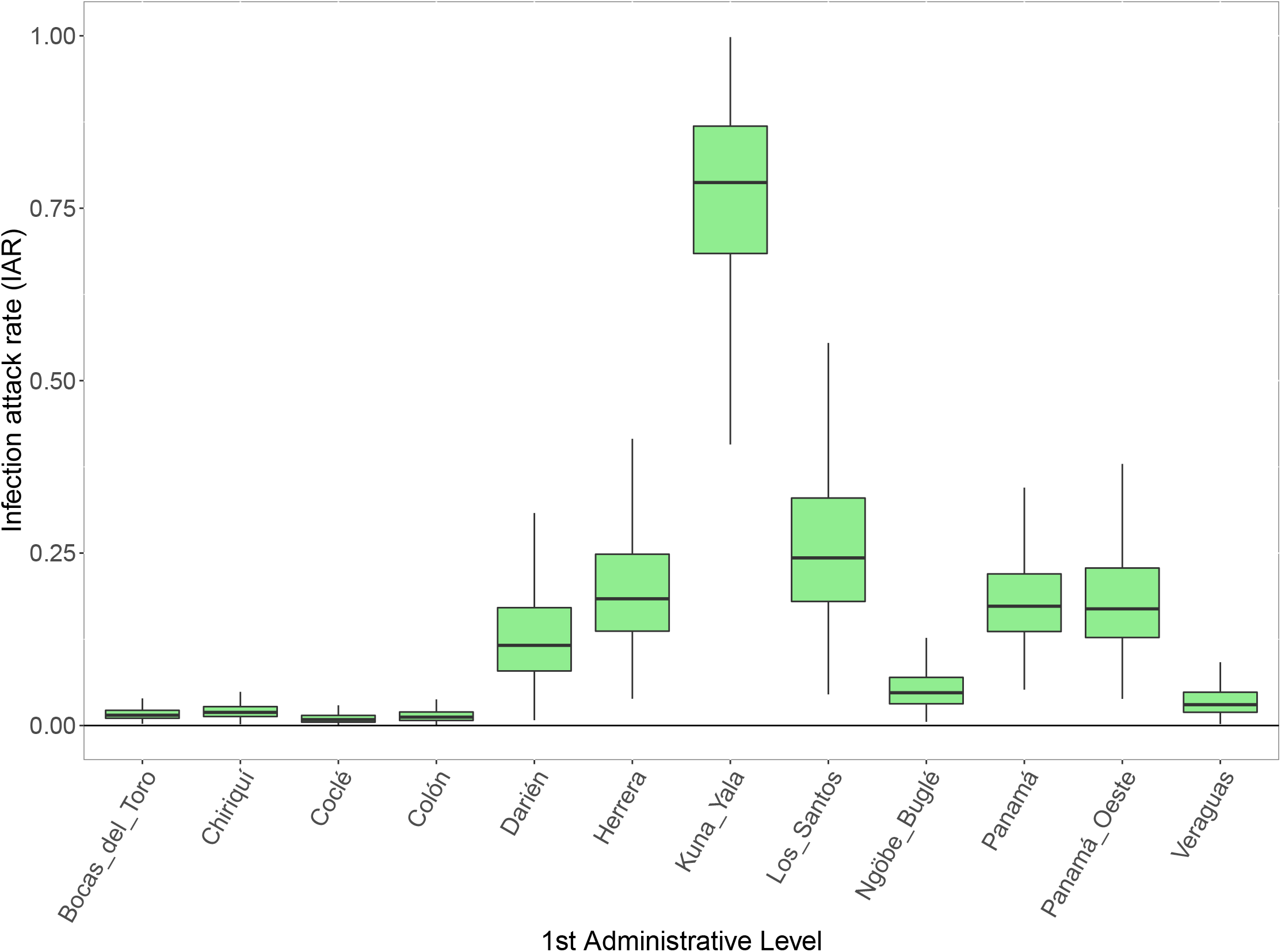
Posterior distribution of subnational infection attack rates (IAR) for Panama.

**Supplementary Figure 20:**
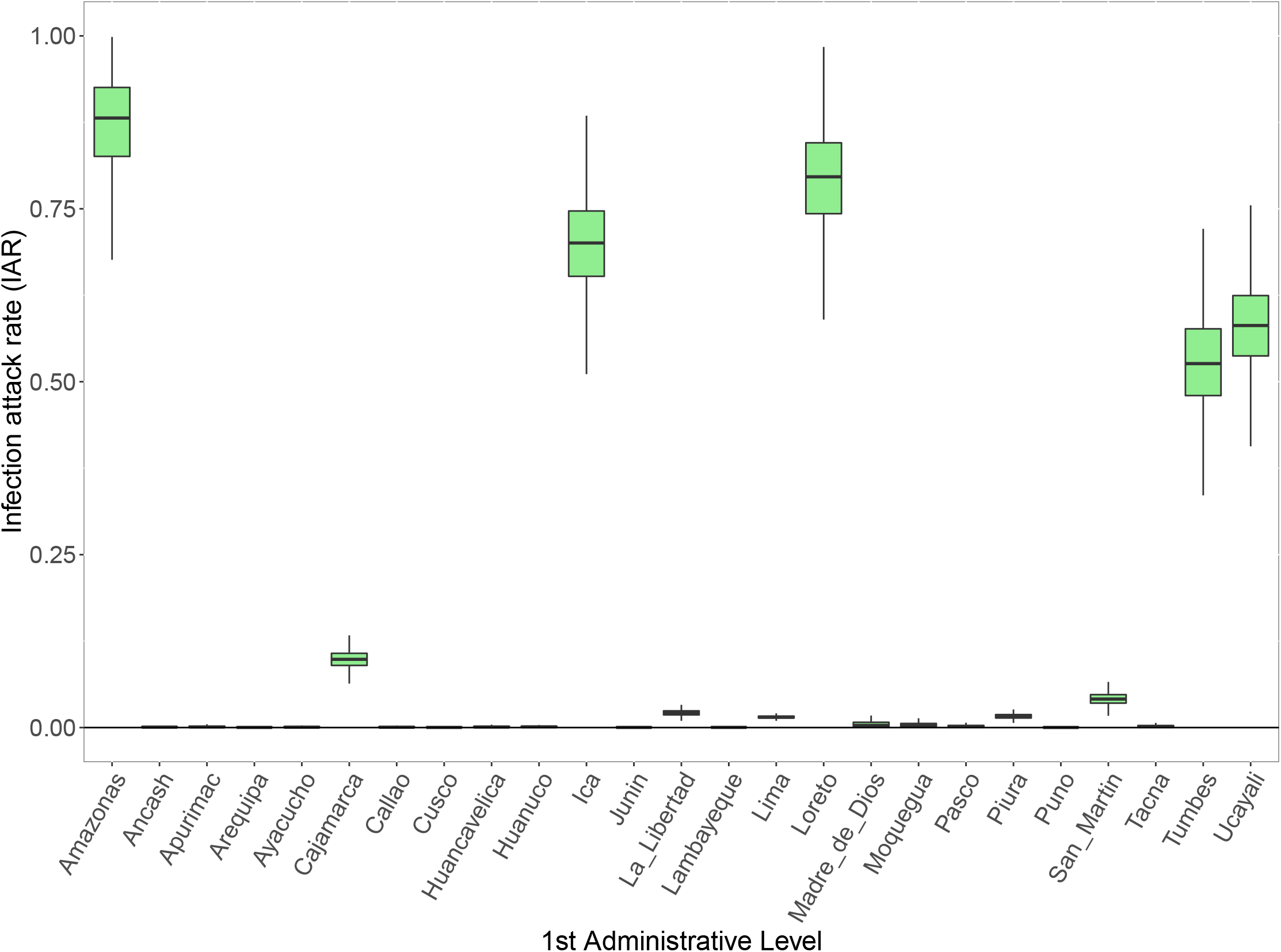
Posterior distribution of subnational infection attack rates (IAR) for Peru.

**Supplementary Figure 21:**
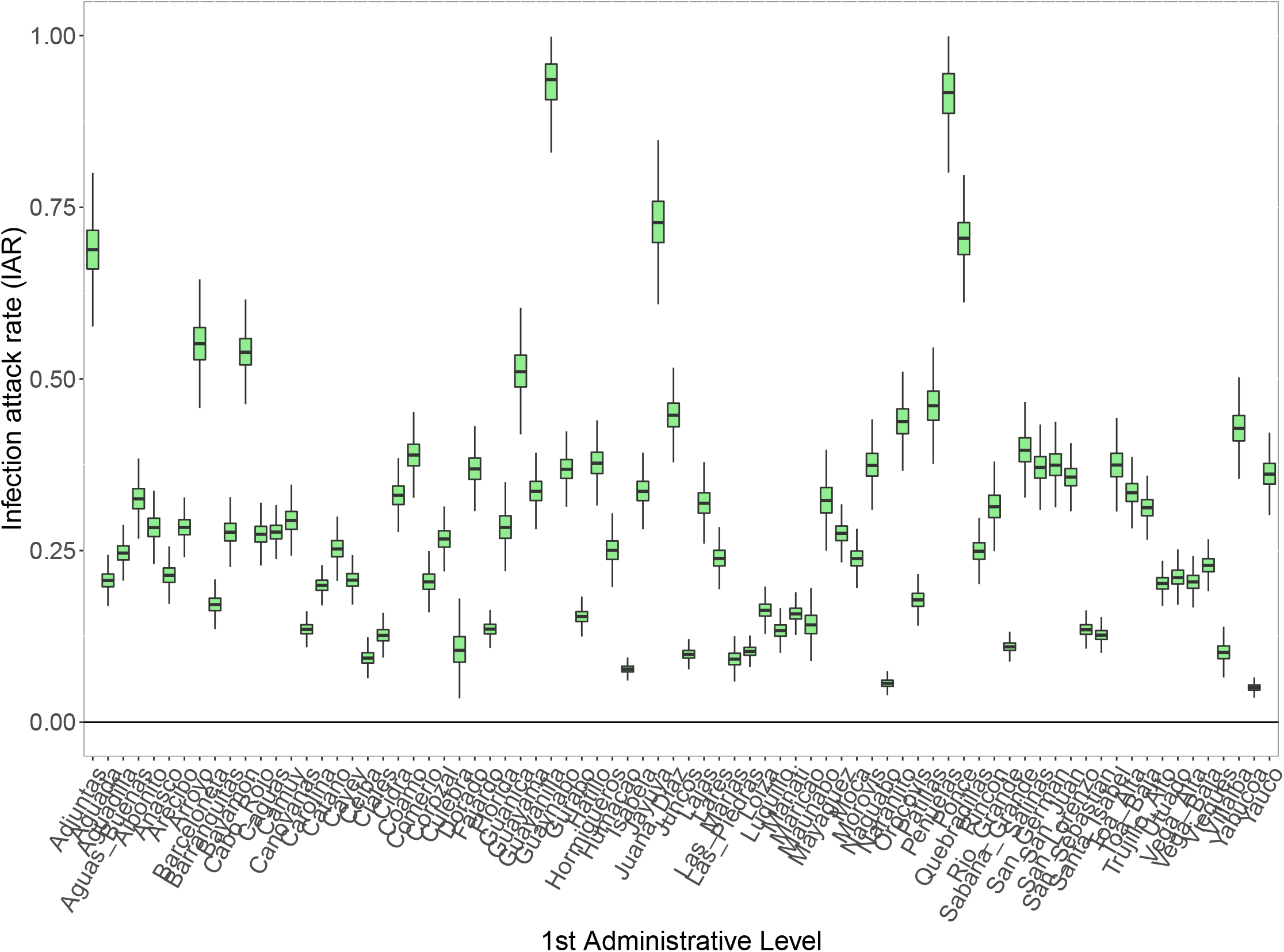
Posterior distribution of subnational infection attack rates (IAR) for Puerto Rico.

**Supplementary Figure 22:**
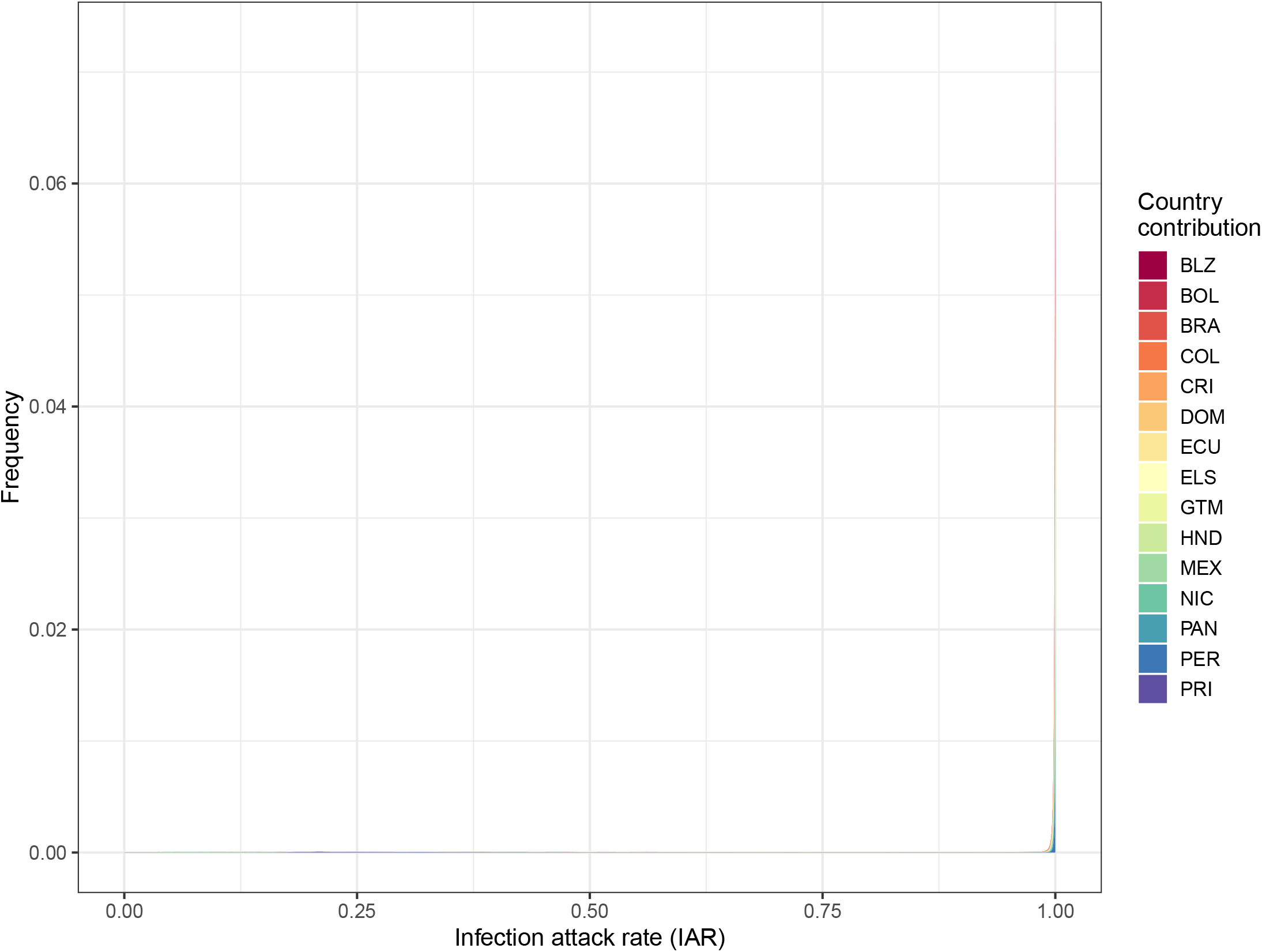
Probability distribution of the infection attack rate (IAR) for Aruba assuming reporting rates estimated from the different national models. The different colors represent the probability distribution of IAR generated from using the estimated reporting rates from each modeled territory.

**Supplementary Figure 23:**
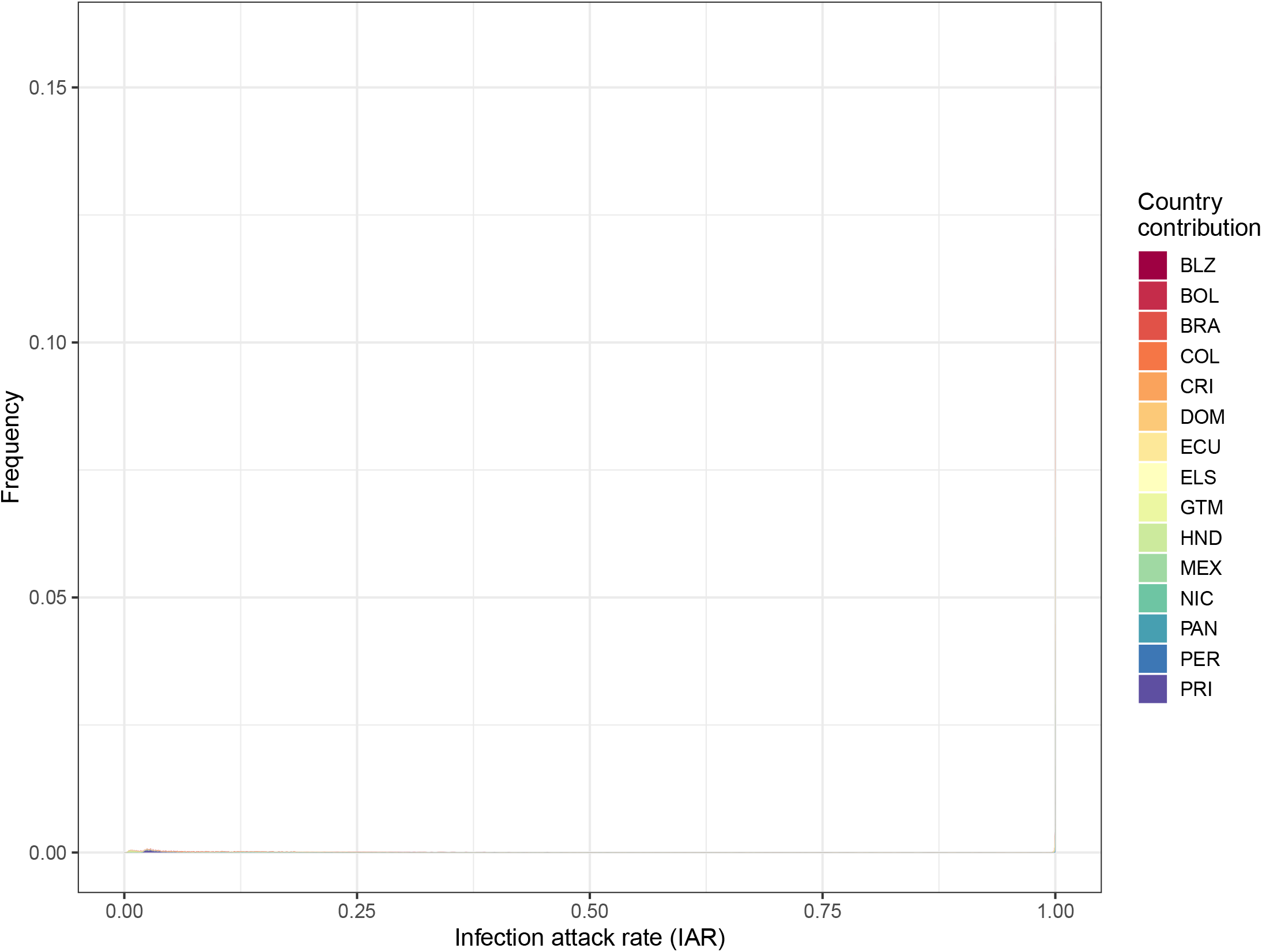
Probability distribution of the infection attack rate (IAR) for Venezuela assuming reporting rates estimated from the different national models. The different colors represent the probability distribution of IAR generated from using the estimated reporting rates from each modeled territory.

**Supplementary Figure 24:**
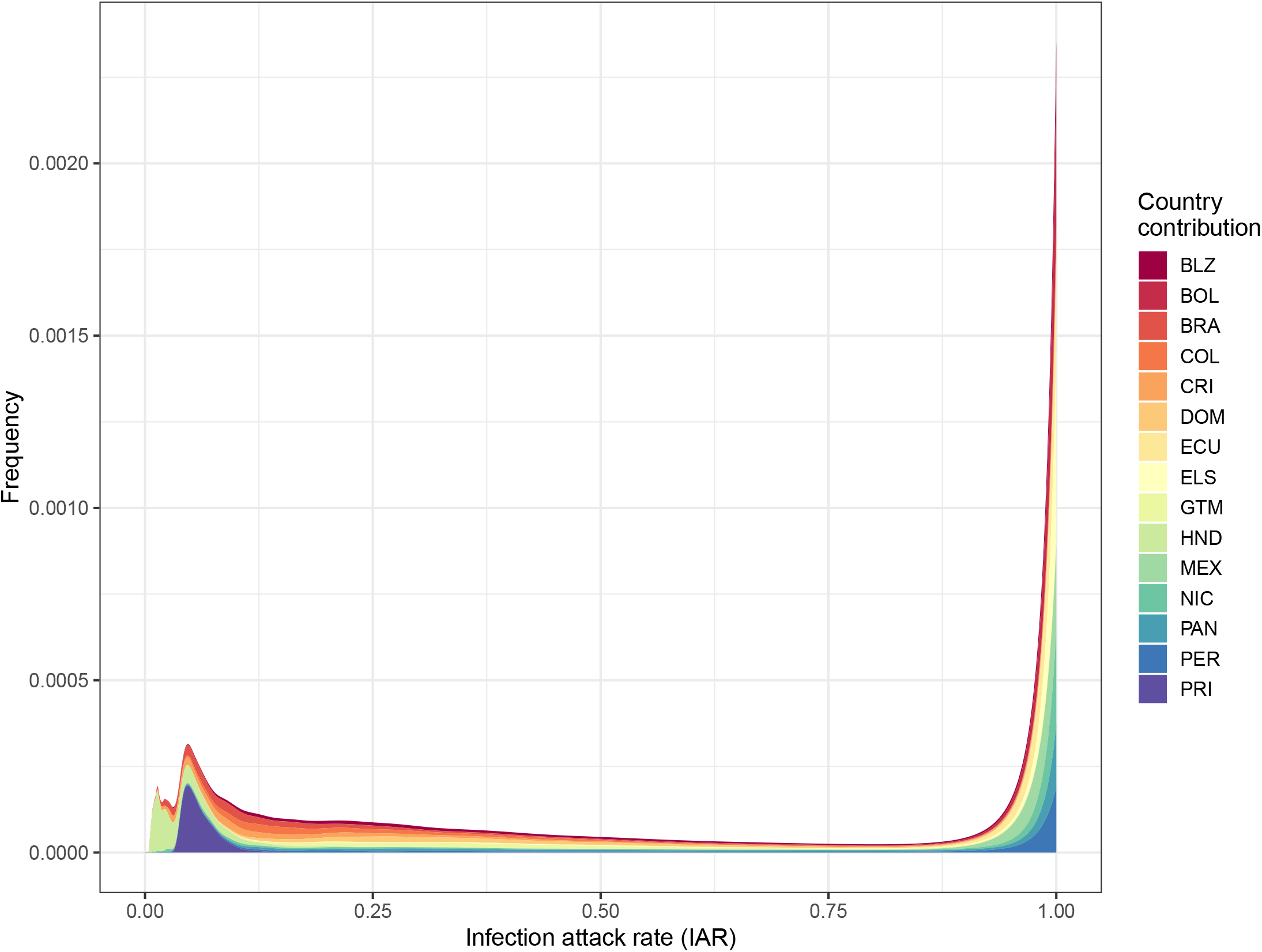
Probability distribution of the infection attack rate (IAR) for the British Virgin Islands assuming reporting rates estimated from the different national models. The different colors represent the probability distribution of IAR generated from using the estimated reporting rates from each modeled territory.

**Supplementary Figure 25:**
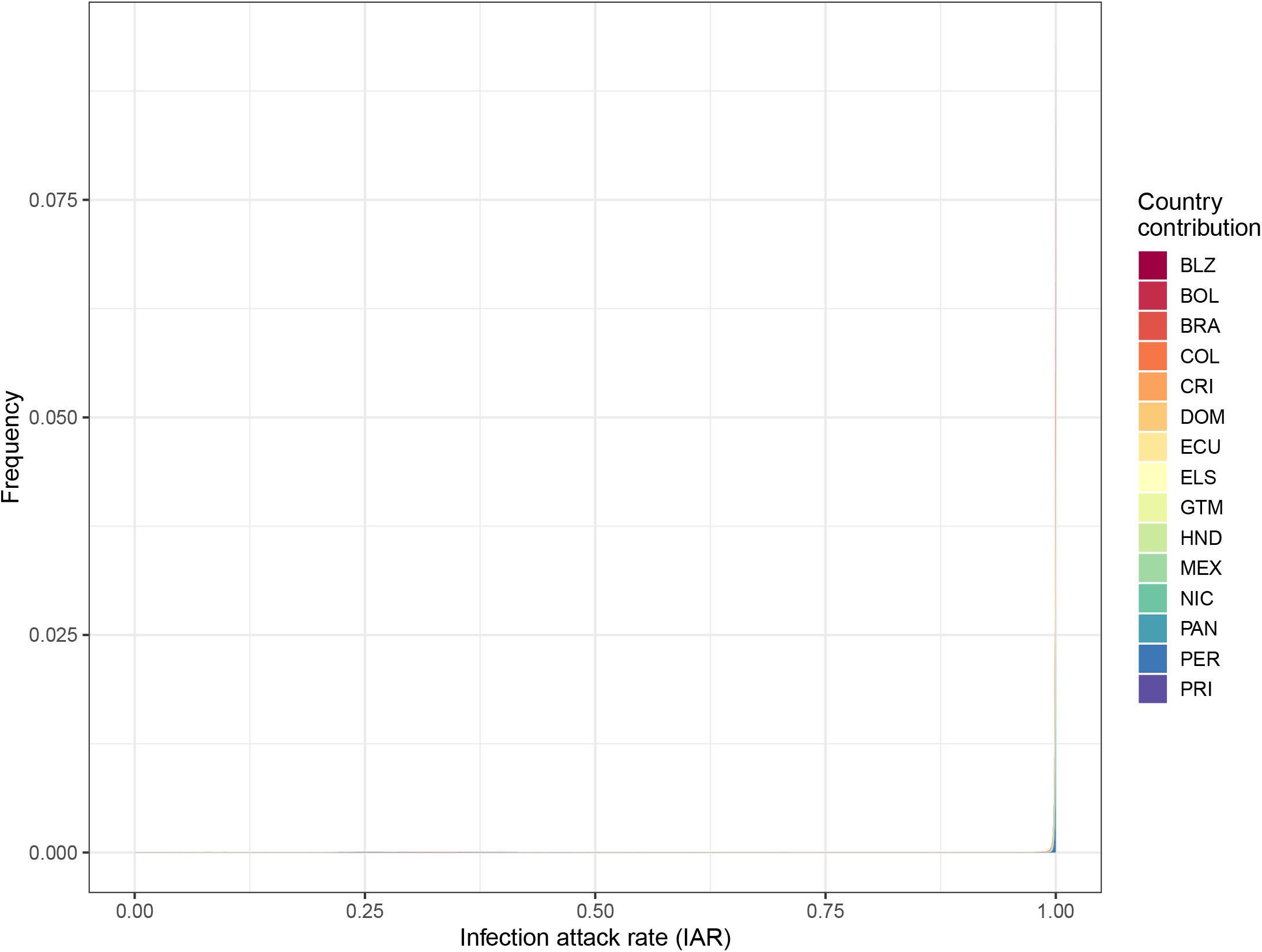
Probability distribution of the infection attack rate (IAR) for the U.S. Virgin Islands assuming reporting rates estimated from the different national models. The different colors represent the probability distribution of IAR generated from using the estimated reporting rates from each modeled territory.

**Supplementary Figure 26:**
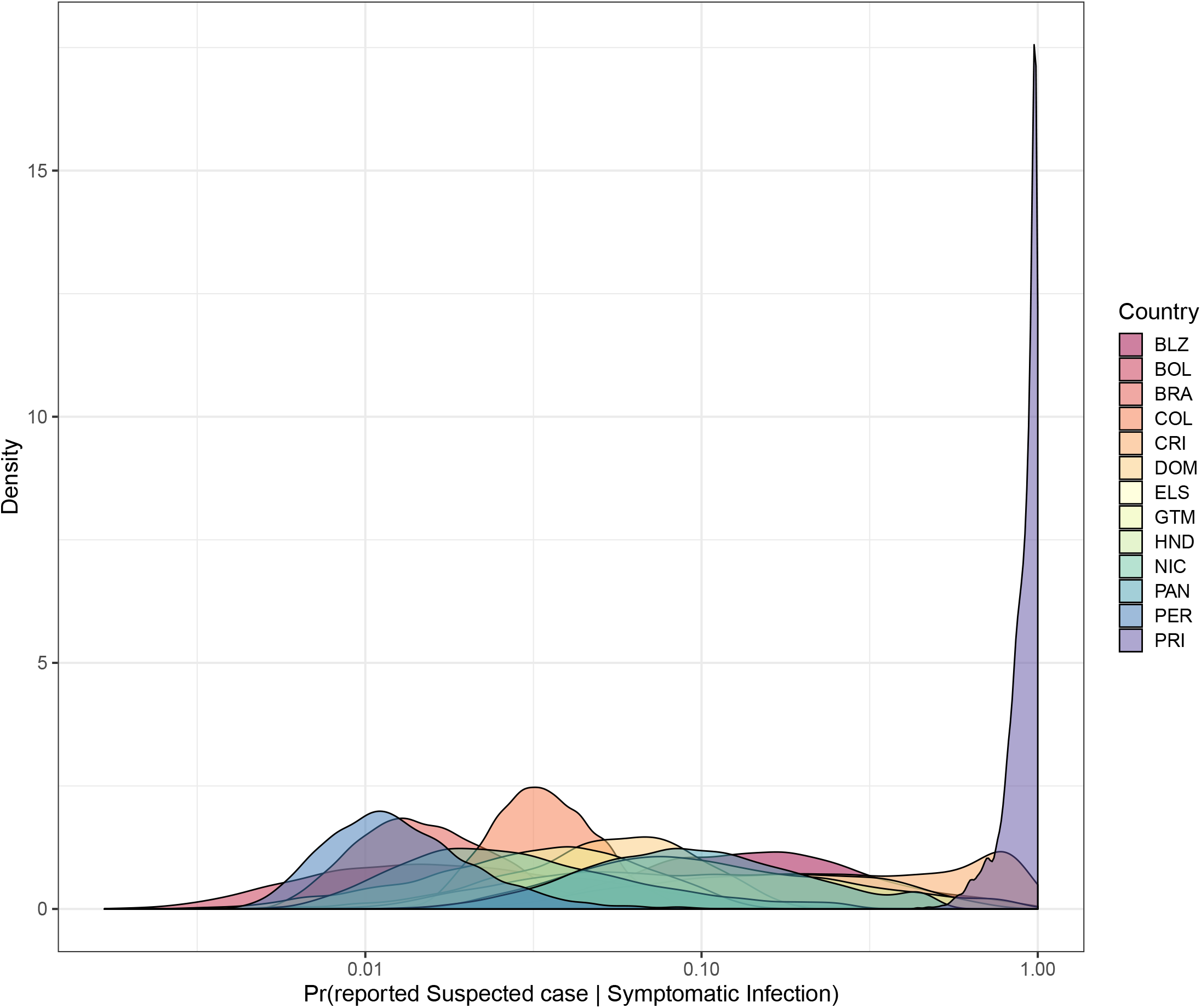
Posterior estimates from each country and territory of the probability that a symptomatic ZIKV infection is reported as a suspected Zika case.

**Supplementary Figure 27:**
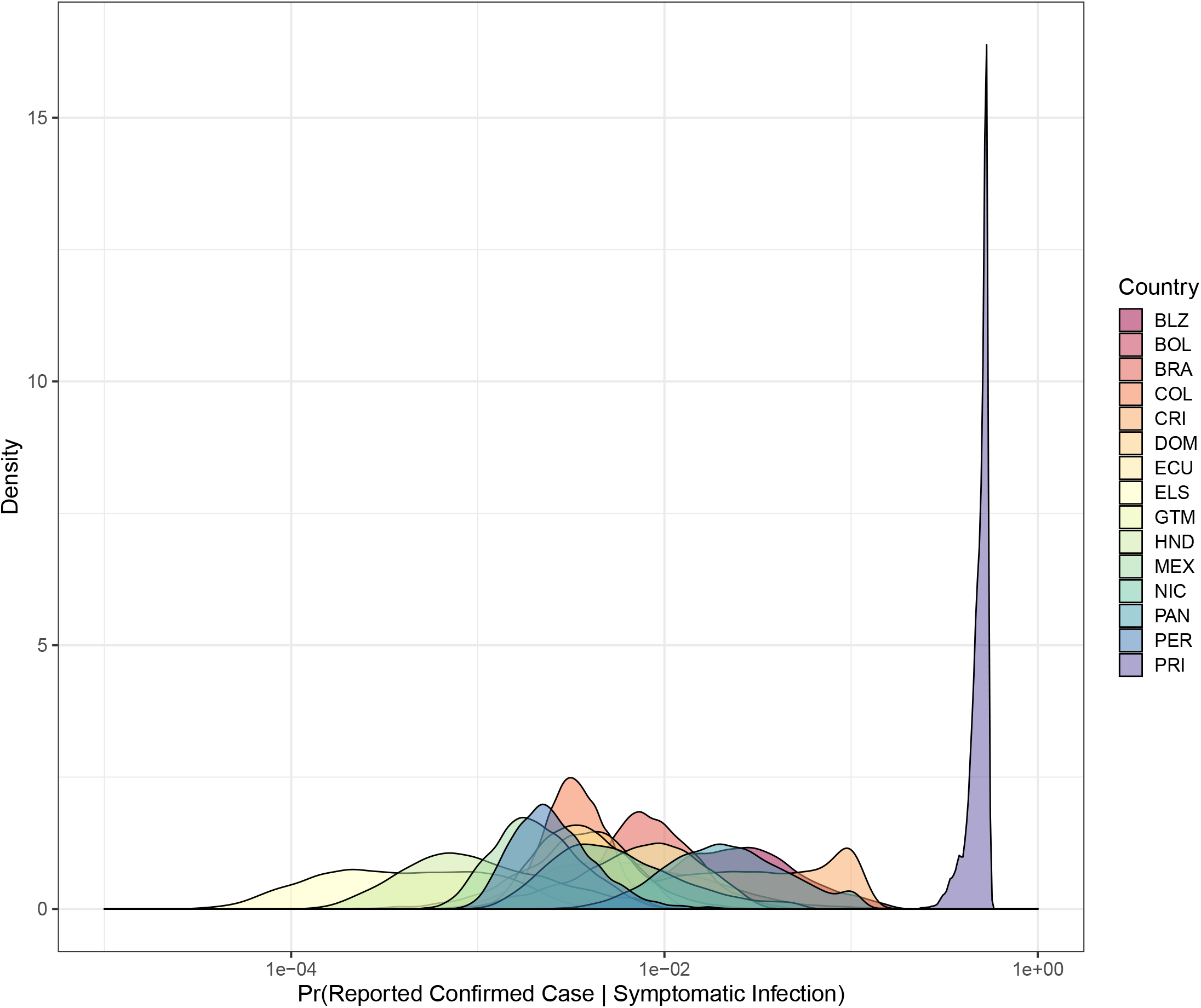
Posterior estimates from each country and territory of the probability that a symptomatic ZIKV infection is reported as a confirmed Zika case.

**Supplementary Figure 28:**
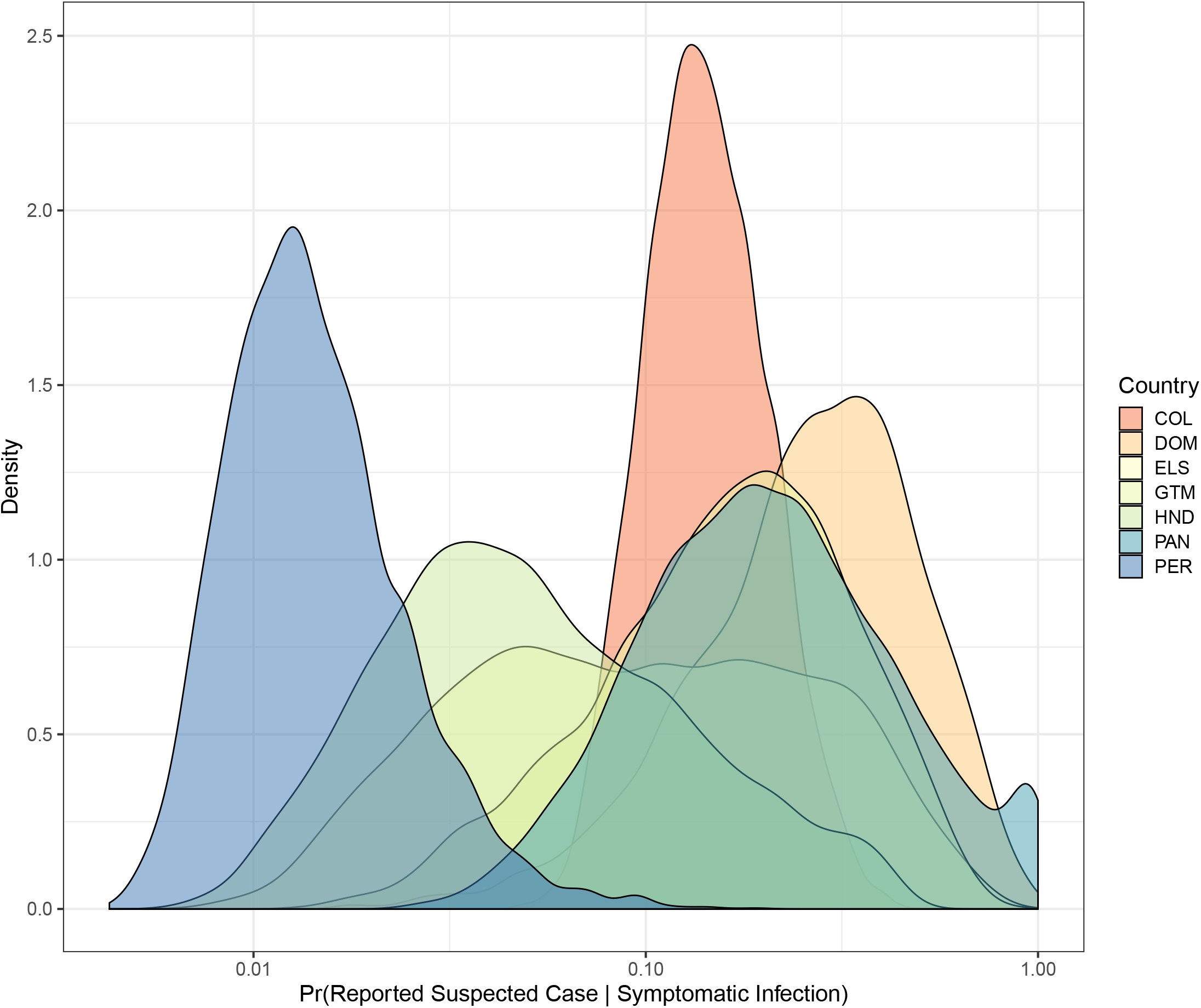
Posterior estimates from each country and territory of the probability that a symptomatic ZIKV infection in a pregnant woman is reported as a suspected Zika case.

**Supplementary Figure 29:**
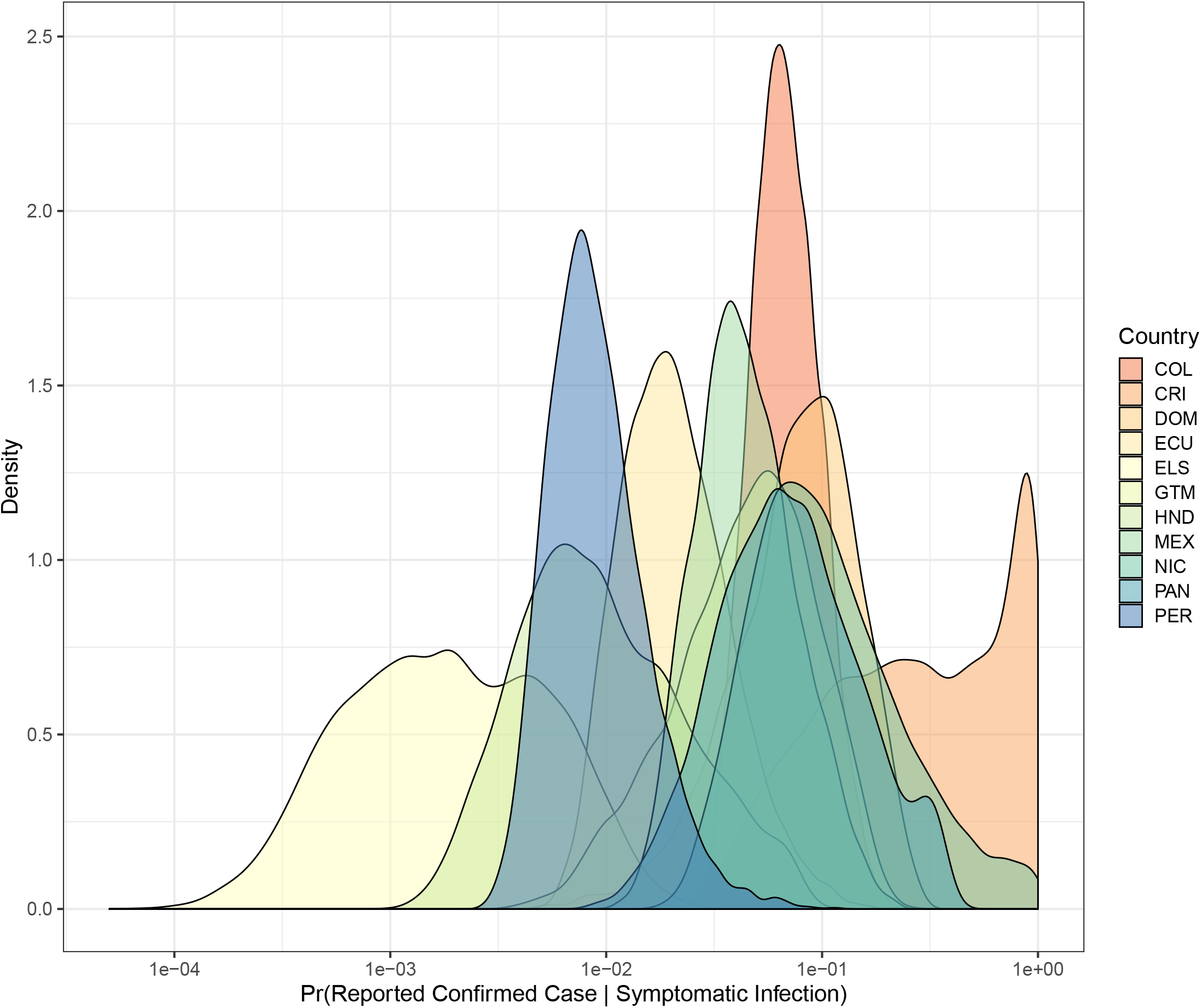
Posterior estimates from each country and territory of the probability that a symptomatic ZIKV infection in a pregnant woman is reported as a confirmed Zika case.

**Supplementary Figure 30:**
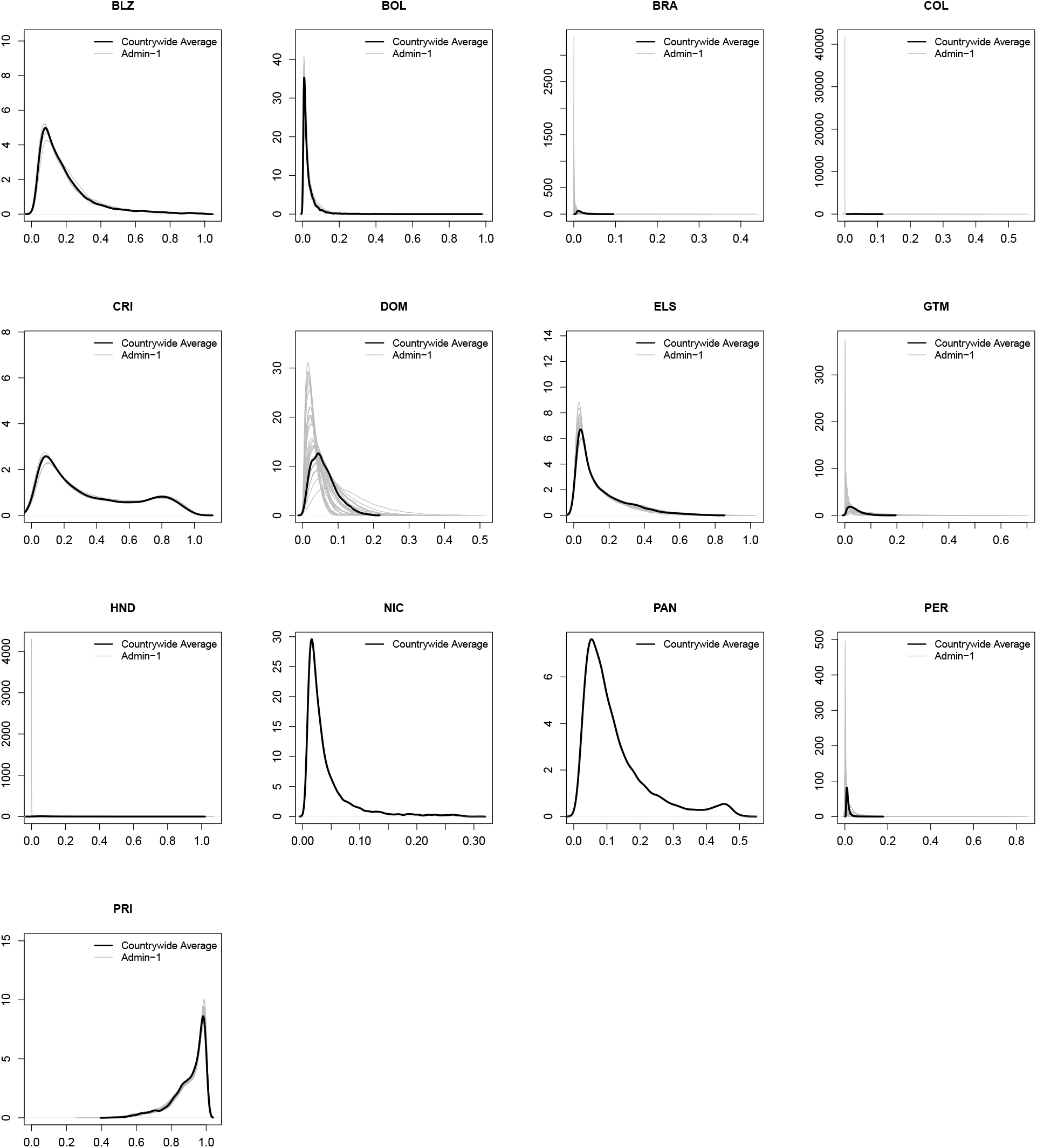
Posterior estimates from each country and territory of the probability that a symptomatic ZIKV infection is reported as a suspected Zika case. Black lines show the country-wide average reporting rate and grey lines show the estimated reporting rate in each administrative unit.

**Supplementary Figure 31:**
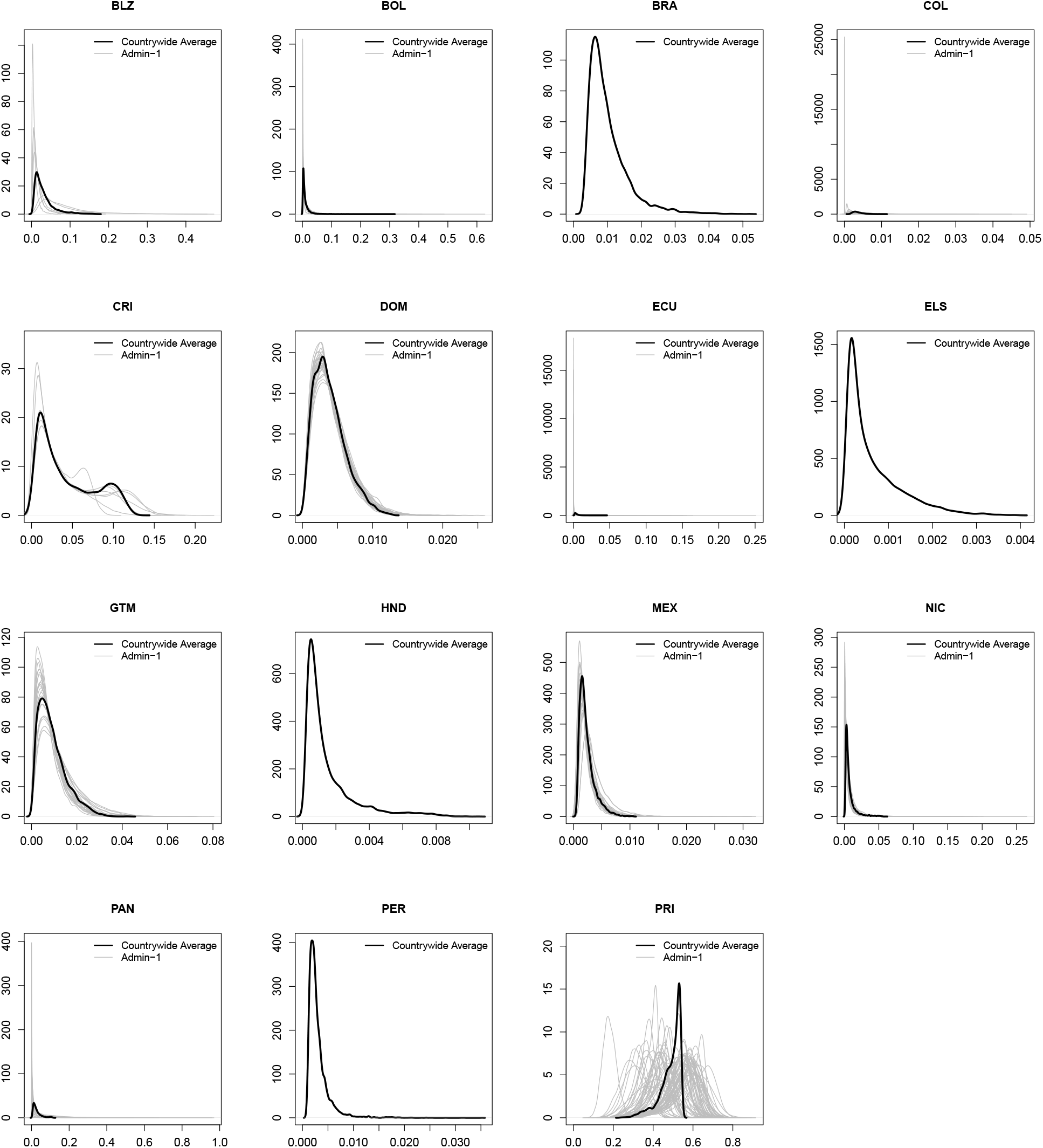
Posterior estimates from each country and territory of the probability that a symptomatic ZIKV infection is reported as a confirmed Zika case. Black lines show the country-wide average reporting rate and grey lines show the estimated reporting rate in each administrative unit.

**Supplementary Figure 32:**
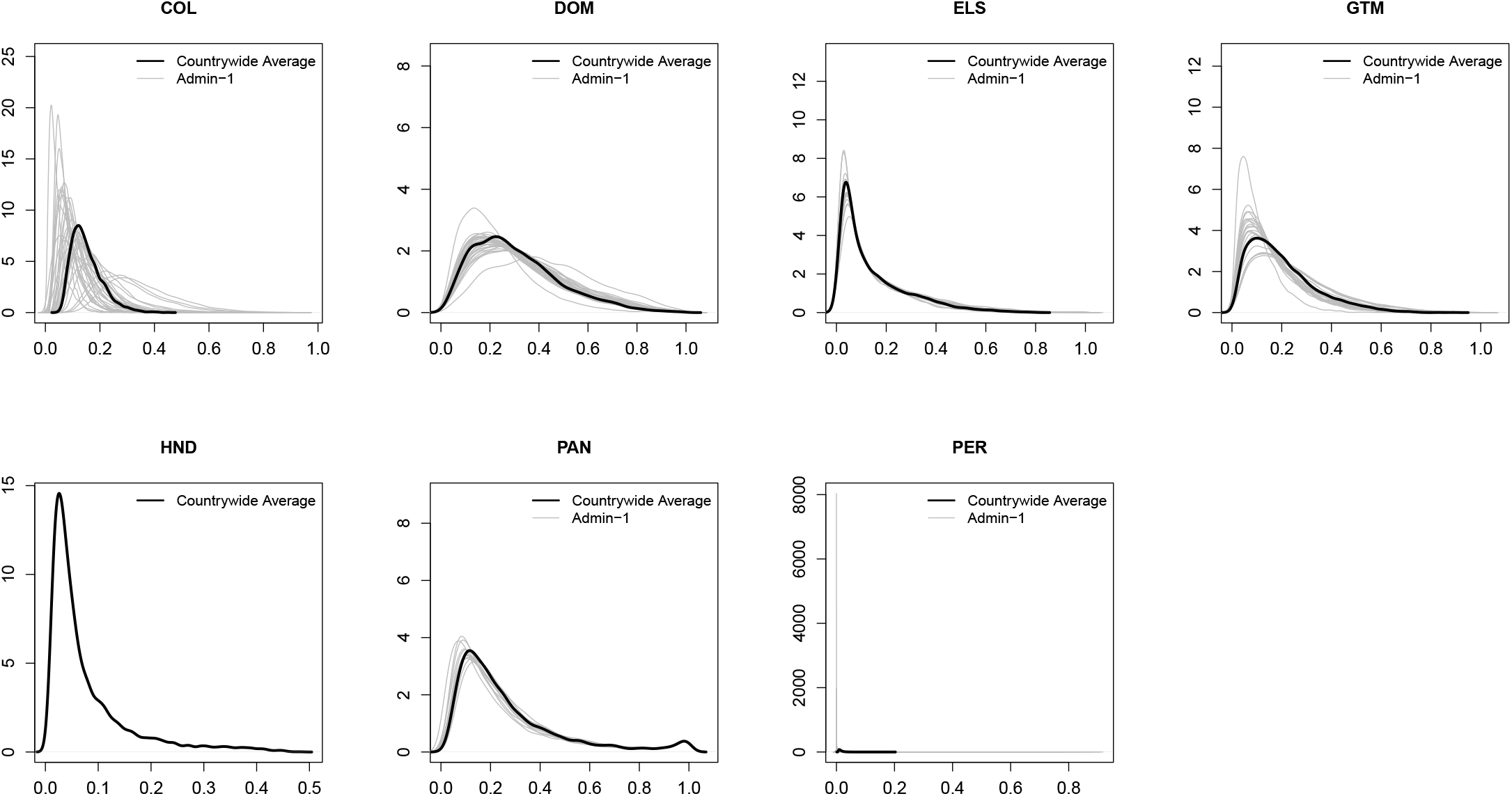
Posterior estimates from each country and territory of the probability that a symptomatic ZIKV infection in a pregnant woman is reported as a suspected Zika case. Black lines show the country-wide average reporting rate and grey lines show the estimated reporting rate in each administrative unit.

**Supplementary Figure 33:**
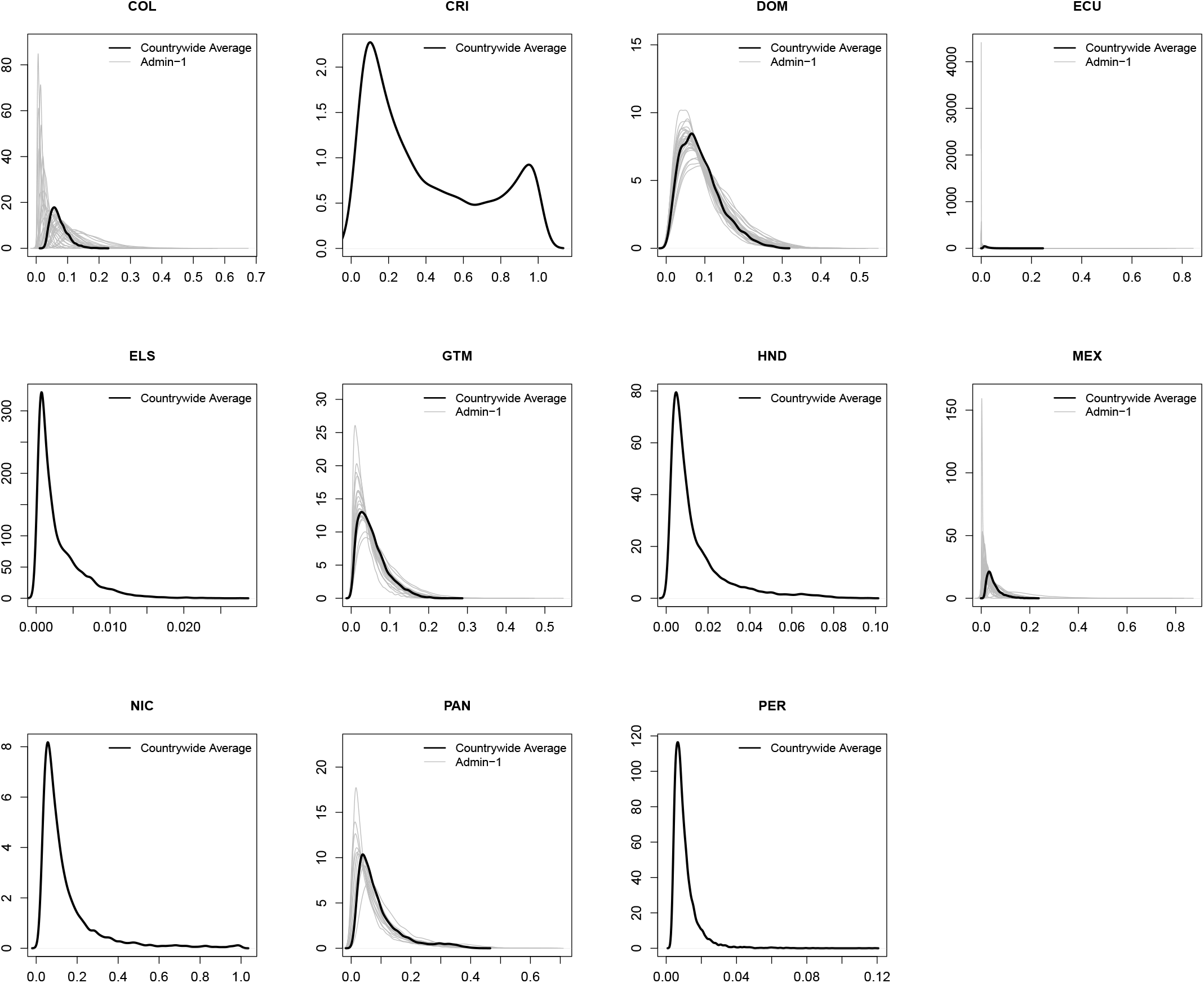
Posterior estimates from each country and territory of the probability that a symptomatic ZIKV infection in a pregnant woman is reported as a confirmed Zika case. Black lines show the country-wide average reporting rate and grey lines show the estimated reporting rate in each administrative unit.

**Supplementary Figure 34:**
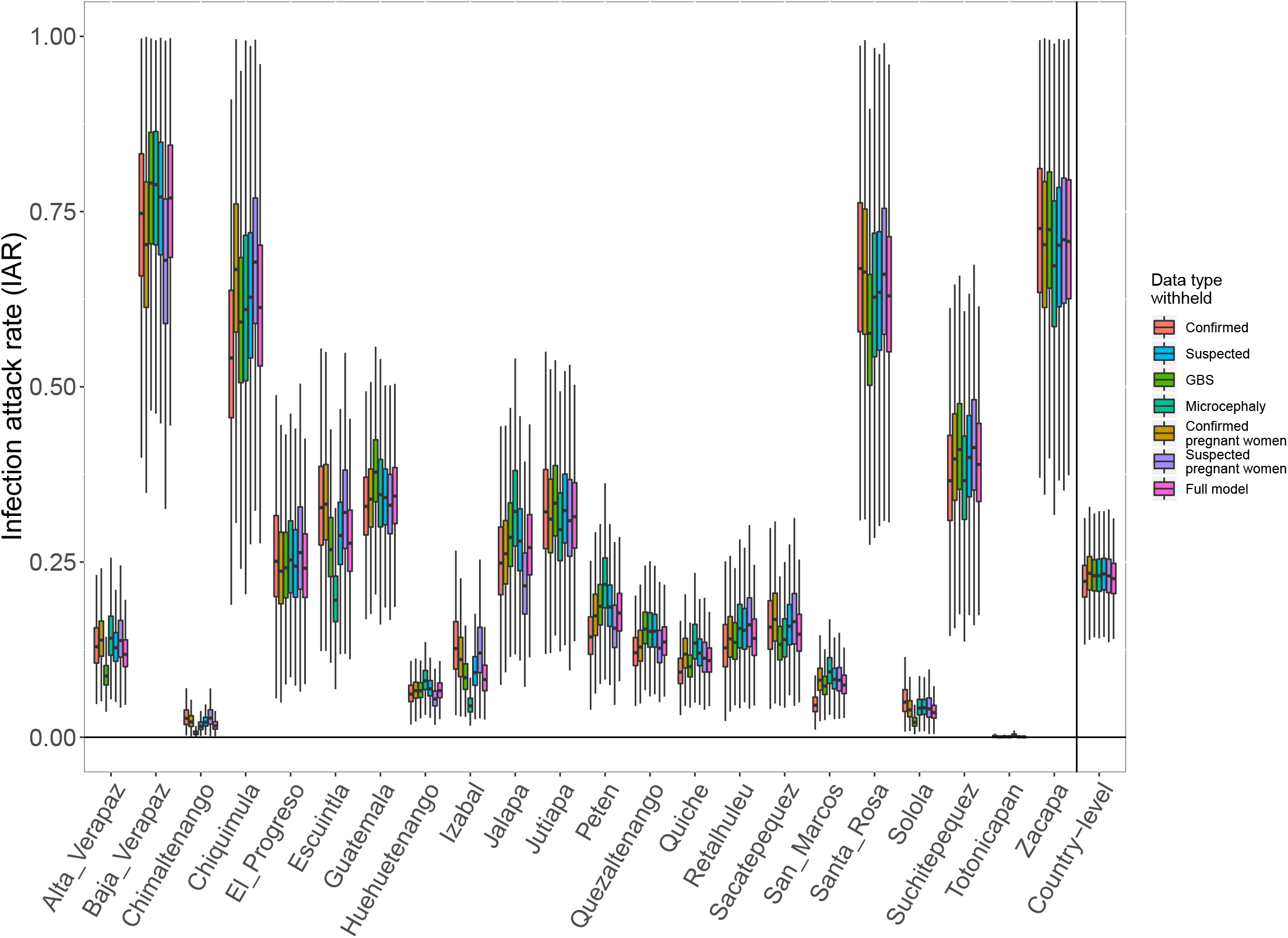
Posterior distributions of national and subnational ZIKV infection attack rates (IAR) for Guatemala when each of the different data types is withheld from the model fitting process.

**Supplementary Figure 35:**
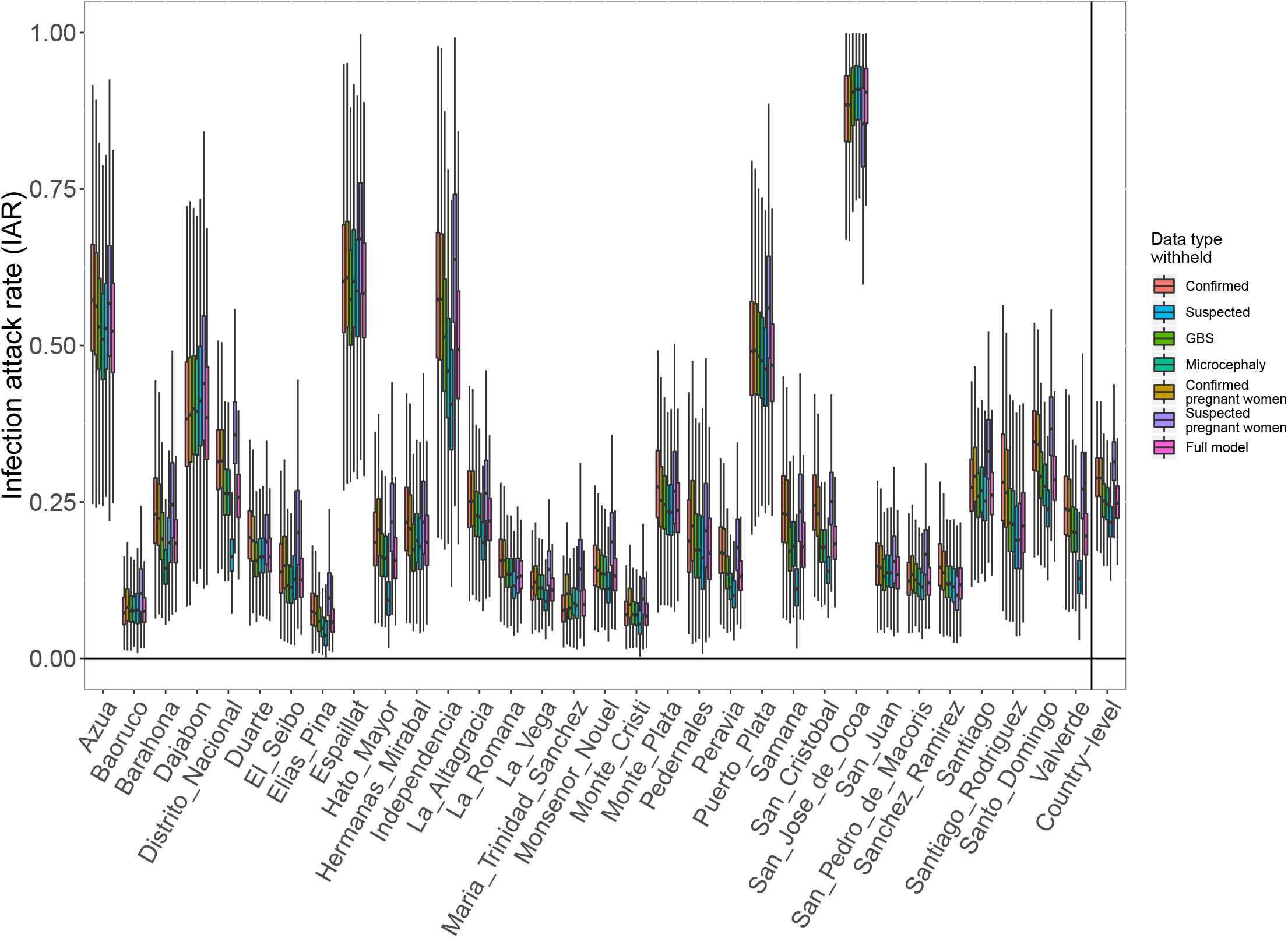
Posterior distributions of national and subnational ZIKV infection attack rates (IAR) for the Dominican Republic when each of the different data types is withheld from the model fitting process.

**Supplementary Figure 36:**
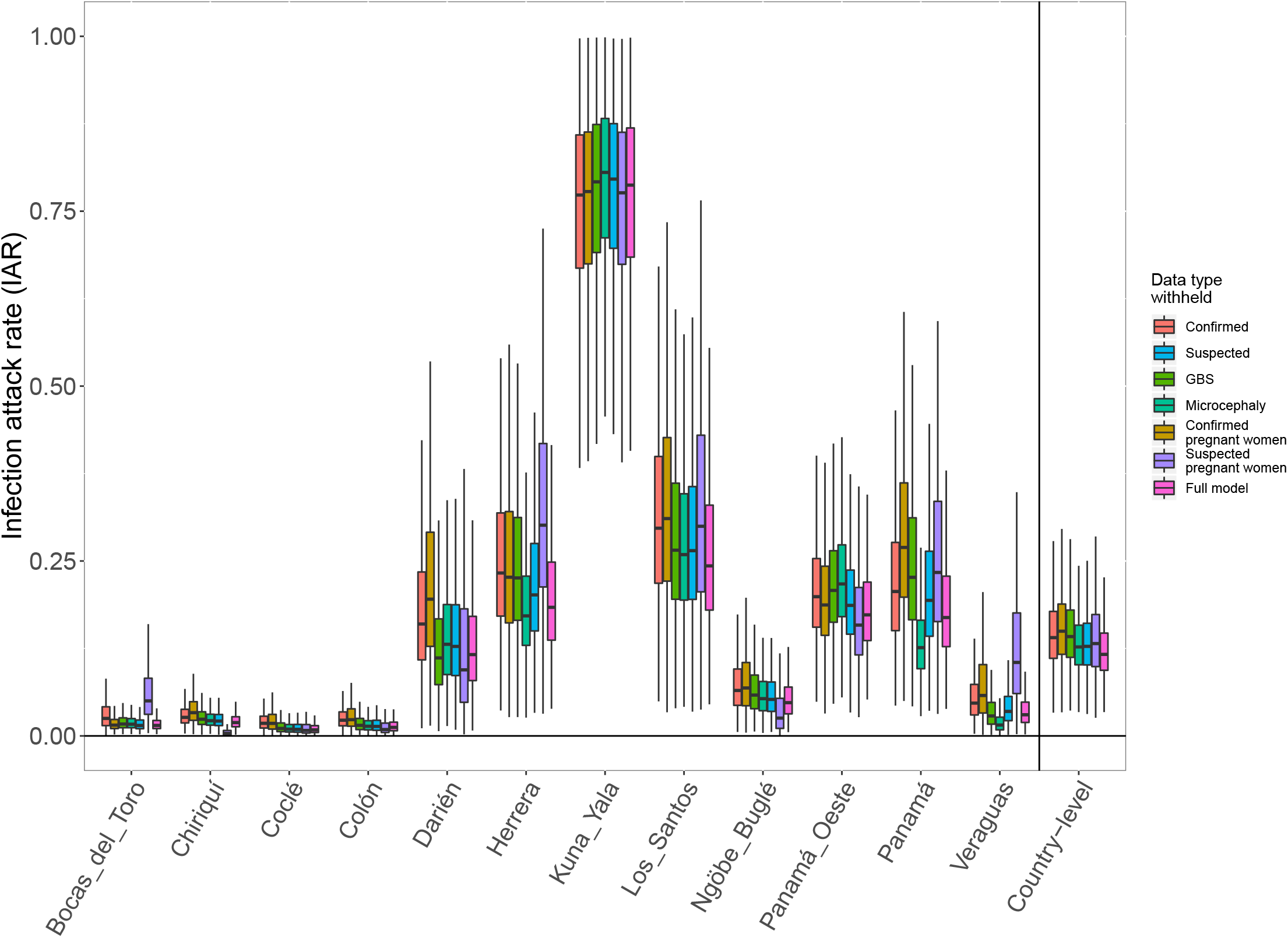
Posterior distributions of national and subnational ZIKV infection attack rates (IAR) for Panama when each of the different data types is withheld from the model fitting process.

**Supplementary Figure 37:**
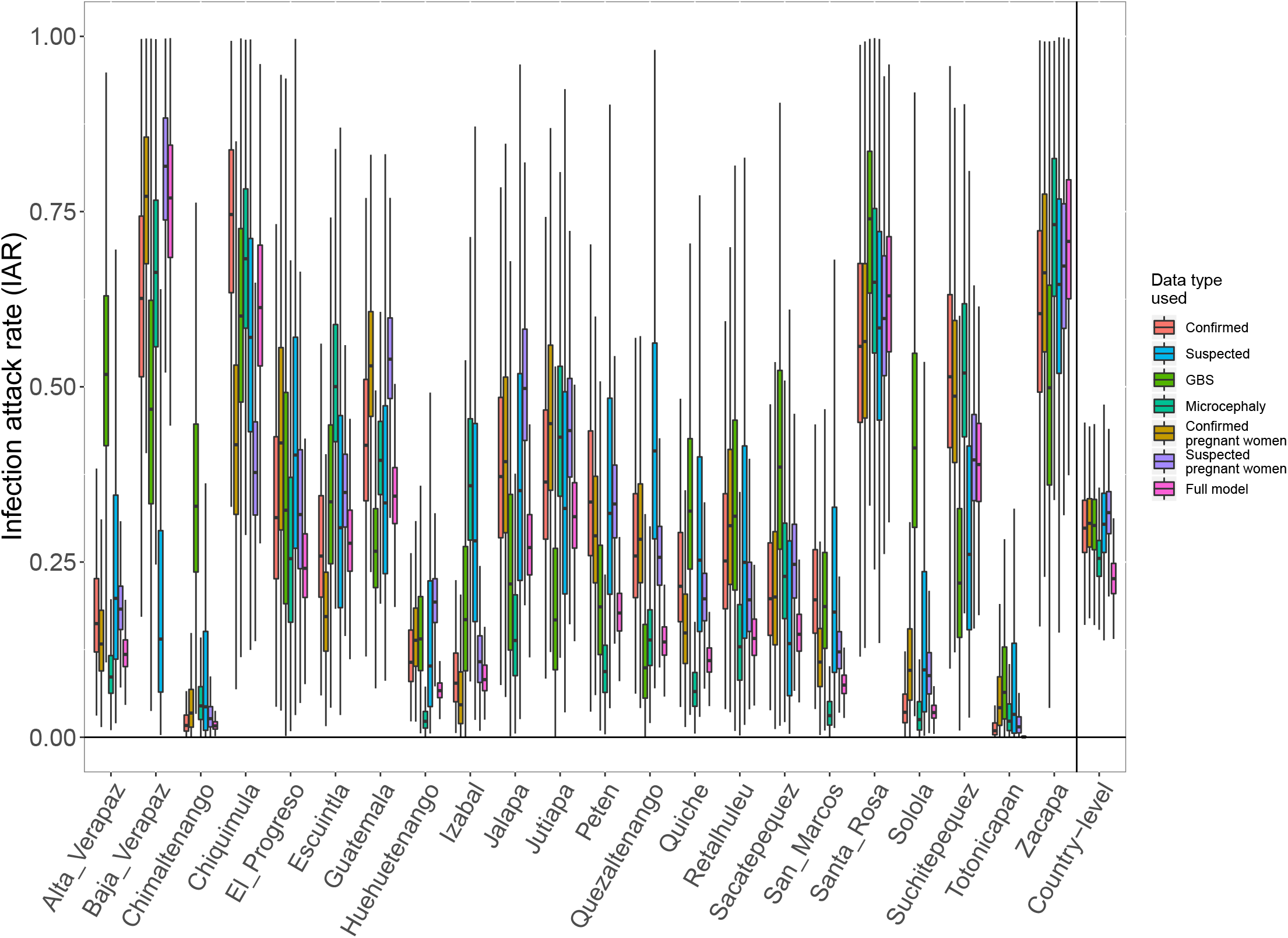
Posterior distributions of national and subnational ZIKV infection attack rates (IAR) for Guatemala from models fit to a single data type in comparison to the full model fit to all data types.

**Supplementary Figure 38:**
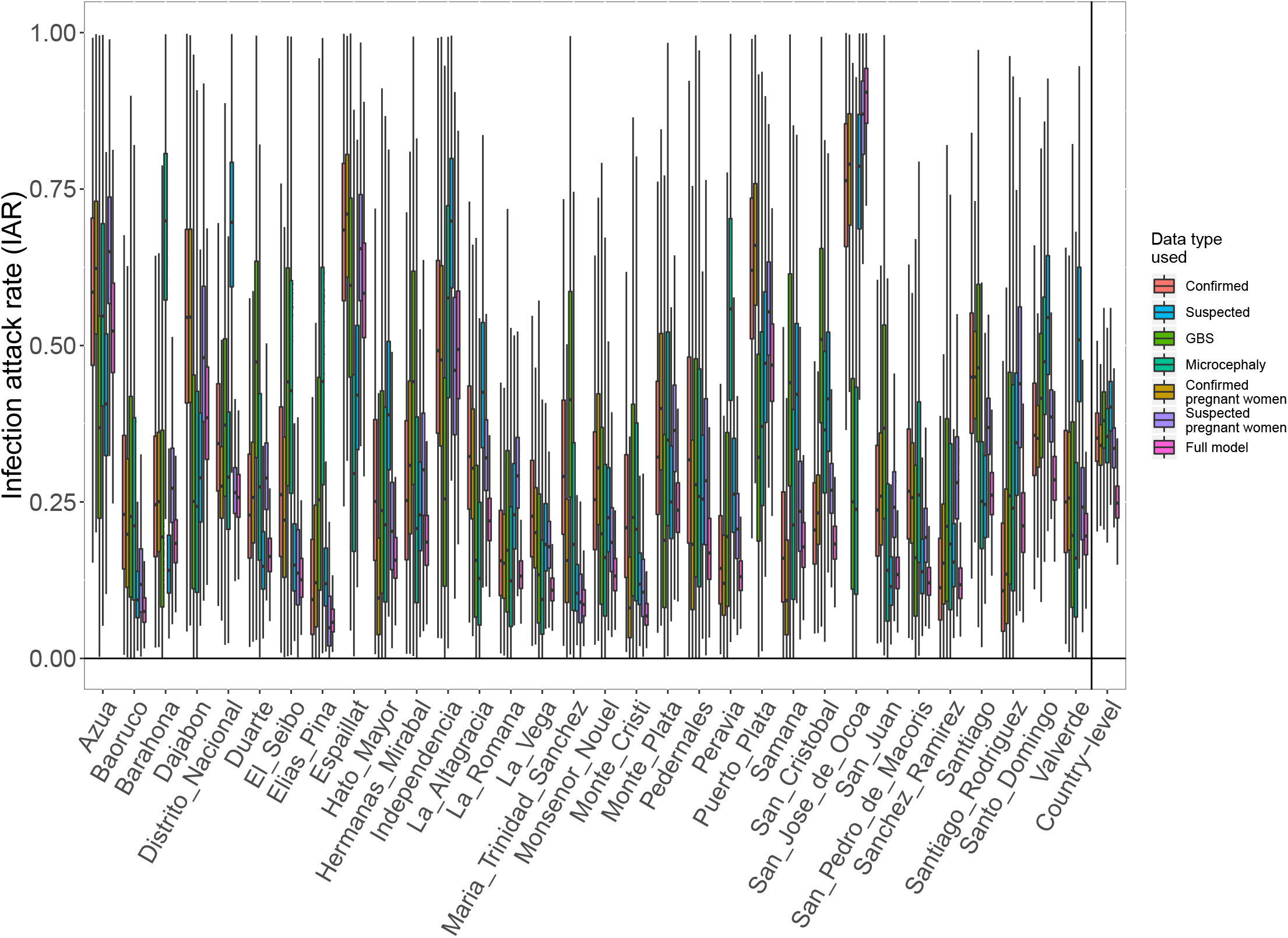
Posterior distributions of national and subnational ZIKV infection attack rates (IAR) for the Dominican Republic from models fit to a single data type in comparison to the full model fit to all data types.

**Supplementary Figure 39:**
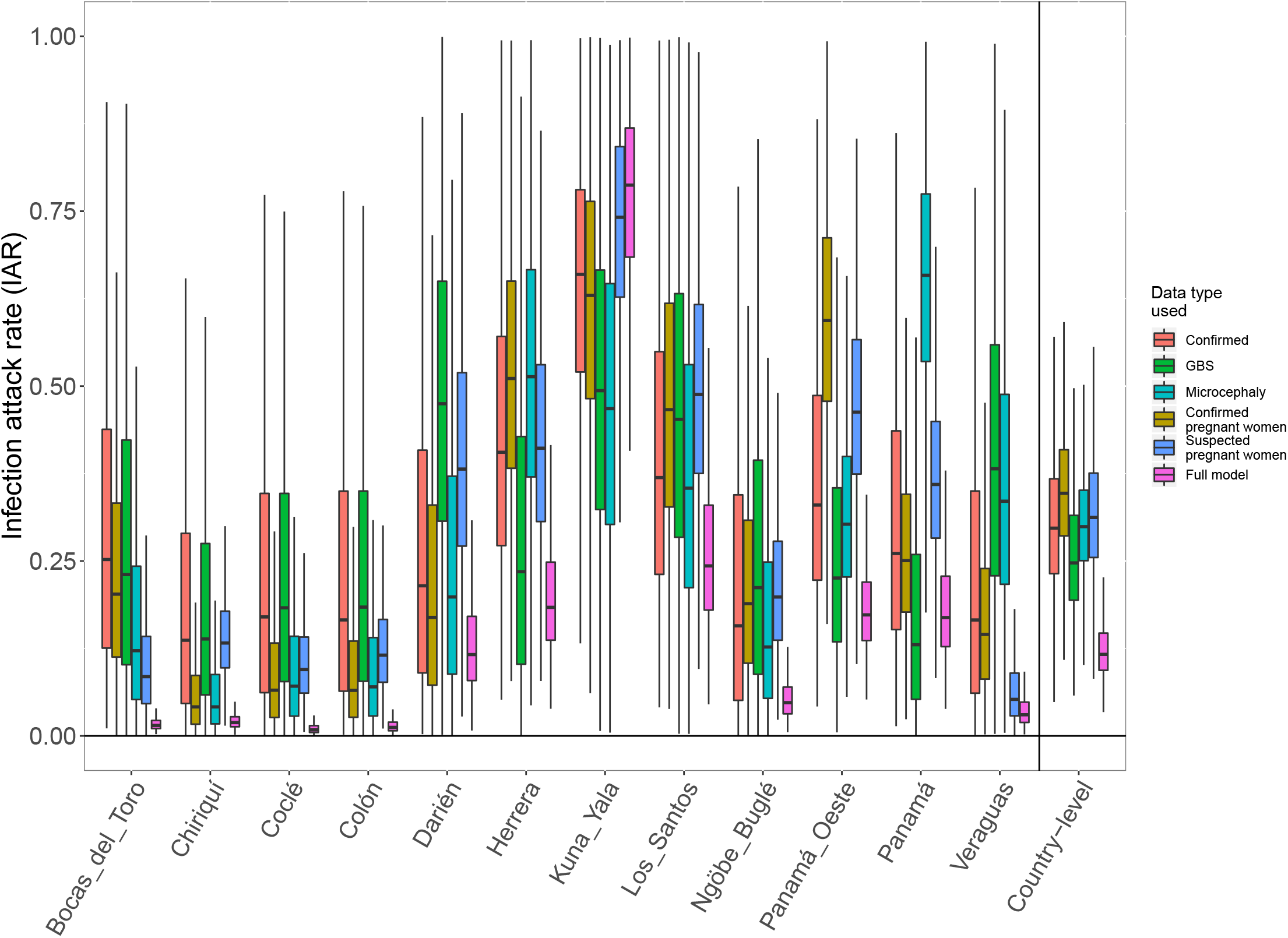
Posterior distributions of national and subnational ZIKV infection attack rates (IAR) for Panama from models fit to a single data type in comparison to the full model fit to all data types.

